# The Effect of Food and Non-alcoholic Beverage Marketing on Children’s Dietary Intake: A Systematic Review and Meta-Analysis

**DOI:** 10.64898/2026.08.06.26359515

**Authors:** Qiuyu Julia Chen, Yuchen Jia, Jaithri Ananthapavan, Brendan T. Smith, Hadis Mozaffari, Dahlia Parolin, Gavin W. K. Wong, Mahsa Jessri

## Abstract

**Importance:** Food and non-alcoholic beverage marketing drives children’s dietary intake, yet updated evidence quantifying effects by marketing medium and sociodemographic factors is needed to inform policy.

**Objective:** To quantify the effect of food marketing on dietary intake among children and adolescents (0-19 years) and examine variations by age, sex, socioeconomic position (SEP), weight status, marketing medium, and exposure duration.

**Data Sources:** Nineteen electronic databases were searched for articles published from April 2020 to February 2026, complemented by World Health Organization-commissioned reviews covering 1970 to March 2020.

**Study Selection:** Two reviewers independently selected peer-reviewed primary studies that assessed the association between food marketing and dietary intake, following PRISMA guidelines, with no language restrictions.

**Data Extraction and Synthesis:** Two reviewers independently extracted data and assessed the risk of bias. Random-effects meta-analyses were conducted. The certainty of evidence was assessed using GRADE.

**Main Outcomes and Measures:** Dietary intake (energy, quantity, or number of items consumed).

**Results:** A total of 55 studies (N = 6,877; range 2-18 years) were included. Food marketing was associated with higher dietary intake (mean difference [MD], 34.8 kcal; 95% CI, 20.2-49.4) compared with no or less marketing. Unhealthy marketing via television (20 studies; MD, 44.5 kcal; 95% CI, 11.2-77.8), digital media (11 studies; MD, 37.5 kcal; 95% CI, 20.1-54.9), and packaging (11 studies; MD, 20.5 kcal; 95% CI, 0.7-40.3) all increased intake; the difference across media was significant (p < .001). Higher intake was observed in males (3 studies; MD, 51.9 kcal; 95% CI, 45.4-58.3) but not in females (MD, -6.8 kcal; 95% CI, -60.3-46.6); difference was not significant (p = .082). Differences by weight status (p = .012) were seen (5 studies; normal weight: MD, 55.6 kcal; 95% CI, -51.3-162.5; overweight/obese: 146.9 kcal; 95% CI, 34.1-259.7). Effects varied by age (p = .003) and by digital media exposure duration (p = .044). One study examined ethnicity; none studied SEP.

**Conclusions and Relevance:** Food marketing is associated with increased dietary intake, with low certainty of evidence. Variations were observed across age, sex, weight status, and marketing medium. Further research is needed for adolescents and the role of SEP.

**KEY POINTS:** *Question:* What are the associations between the marketing of food and non-alcoholic beverages and children’s dietary intakes?

*Finding:* In this systematic review and meta-analysis of 55 experimental studies with 6,877 participants, food marketing was associated with higher dietary intake (34.8 kcal; 95% CI, 20.2-49.4 kcal). Some variation was observed across age, sex, weight status, and marketing medium, although evidence was limited for adolescents and by socio-economic status.

*Meaning:* These findings suggest that comprehensive, cross-platform marketing restrictions could help protect children’s health. Demographic factors are important moderators in the development of policy evaluation models.

## INTRODUCTION

Unhealthy diets are a key modifiable risk factor for childhood overweight and obesity worldwide.^1,2^ Childhood obesity often persists into adulthood and increases the risk of the earlier onset of noncommunicable diseases, such as cardiovascular diseases, type 2 diabetes mellitus, and some cancers, accounting for 75% of all deaths globally.^3–6^ Marketing of food and non-alcoholic beverages [hereafter referred to as “food”] that are high in saturated fats, free sugars, and sodium (HFSS) is a major environmental contributor to poor dietary patterns in children.^7^ The World Health Organization (WHO) and other international organizations have called for regulatory action to promote children’s dietary health by creating a healthier food environment and protecting children from unhealthy food marketing.^8,9^ Some countries have implemented policies restricting all food marketing to children,^10–12^ while others limit marketing of unhealthy foods based on nutrient profiling.^13,14^

Recent systematic reviews and meta-analyses show that food marketing to children (M2K) increases consumption of unhealthy products among children and adolescents and influences their dietary preferences.^15–20^ Several WHO-commissioned reviews, including publications from 1970 to March 2020, examined the effect of M2K on children’s dietary and health outcomes.^15,21,22^ M2K also results in a stronger preference for promoted products.^15^ These effects are observed across multiple media, including television (TV), print media, digital media, and packaging.^15,21,22^

While several reviews have examined the effect of M2K,^15–22^ none provide a comprehensive, up-to-date quantitative synthesis of dietary intake across all media for the 0–19 age group with sufficient granularity for policy outcome modeling and simulation studies. Meta-analyses of randomized trials show that acute exposure to unhealthy marketing increases energy intake by 30–60 kcal under experimental conditions, though evidence is limited to a single marketing channel or pre-digital expansion.^18,19^ Meta-analyses of screen advertising indicate that increases in intake are more pronounced among children with obesity compared to those with a normal weight.^18^ Additionally, although children from lower socioeconomic positions (SEP) and ethnic minority backgrounds face disproportionate exposure to unhealthy advertising,^23^ quantifiable differences in effect size across these groups are lacking. Without these estimates, the potential impacts of M2K regulations on health inequities cannot be assessed in policy cost-effectiveness modeling studies.^24^ Thus, our meta-analysis aims to fill this gap by quantifying the effect of food marketing on dietary intake among children and adolescents (ages 0-19 years) and examining variation by age, sex, ethnicity, SEP, weight status, marketing medium, and duration of exposure.

## METHOD

This systematic review and meta-analysis were conducted in accordance with the Preferred Reporting Items for Systematic Reviews and Meta-analyses (PRISMA) 2020 guidelines. The study protocol was prospectively registered with PROSPERO (CRD42025641870) in February 2025 and subsequently published in Systematic Reviews (November 2025; doi: 10.1186/s13643-025-02978-x). This article focuses specifically on the quantitative synthesis of experimental evidence as part of the protocol.

### Information Sources and Search Strategy

The literature search was conducted in two parts to identify evidence from 1970 to February 2026. For studies published after April 1, 2020, a bibliographic database search was developed and conducted in November 2024 and updated in February 2026 by the researcher (Q.C.) in consultation with a Health Science librarian specializing in human nutrition (K.M.). The database selection and search strategy were informed by two WHO-commissioned global reviews that assessed the M2K effect on dietary and health outcomes: Cairns et al.^22^ (covering evidence from 1970 to 2008) and Boyland et al.^15^ (covering evidence from 2009 to March 2020). The databases searched included MEDLINE, CINAHL, Web of Science, Embase, ERIC, The Cochrane Library (CDSR, CENTRAL), Business Source Ultimate, Communication & Mass Media Complete, Database of Promoting Health Effectiveness Reviews (DoPHER), EconLit, Emerald, Global Index Medicus, Healthevidence.org, IRIS (Institutional Repository for Information Sharing), JSTOR, KOREAMED, Index to Legal Periodicals & Books Full Text, The Campbell Library, and TRIP (Turning Research Into Practice). The full search strategy for the updated database search is available in eTable 1. Studies that met this study’s inclusion criteria from the two WHO-commissioned reviews were included to retrieve publications from 1970 to March 2020.^15,22^ To cross-reference, Scopus was used to conduct a backward citation search of the other 51 relevant reviews (eTable 2) that may contain targeted studies identified through the database search.

### Selection Criteria

All records identified through database and citation chain searches were imported into Covidence, which detected and removed duplicate results.^25^ Subsequently, each record was independently screened by two reviewers in a two-stage process. The agreement rate between reviewers and the detailed selection process was reported in the review protocol.^26^ Eligibility criteria are reported in Table 1.

**Table 1.** Inclusion and Exclusion Criteria.

| **Criterion type** | Description |
| --- | --- |
| **Inclusion criteria** |  |
| Language | No restriction |
| Population | Children and adolescents aged 0 to 19 years |
| Study design | Experimental studies in which participants were exposed to food and non-alcoholic beverage marketing and compared with a control condition exposed to non-food marketing or no marketing at all. |
| Exposure | - The intervention criteria encompassed any form of marketing that aligned with the WHO guideline for Policies to Protecting Children from the Harmful Effects of Food Marketing,<sup>1</sup> “any form of commercial communication, message or action that acts to advertise or otherwise promote a product or service, or its related brand, and is designed to increase, or has the effect of increasing, the recognition, appeal and/or consumption of products or services.”<br>- Marketing of both unhealthy and healthier products. The healthfulness of marketed food was determined using the studies’ definition when stated; otherwise, it was categorized by the researchers based on whether the marketed food was HFSS and/or UPF, or whether the food brand marketed was known for HFSS and/or UPF, such as fast-food chain restaurants, candies, or soft drinks. Examples of healthier options included fruits, vegetables, and milk. |
| Outcomes | - Food intake is quantified as energy (kilocalories [kcal] or kilojoules [kJ]), weight (grams or ounces), or the number of items consumed.<br>- Studies reporting overall outcomes and/or subgroup results—such as by age, sex, ethnicity, and socioeconomic position (SEP)—were included to capture heterogeneous effects across different population groups. |
| **Exclusion criteria** |  |
| Study design | Unpublished manuscripts, conference abstracts, and materials containing subjective or opinion-based information, such as editorials, commentaries, letters, or blogs, were excluded due to uncertainties regarding their peer review status. |
| Outcomes | - Outcomes such as children’s food choices, preferences, and purchasing behaviors were excluded, as this review focuses on direct measurement of food intake.<br>- Given the focus on quantitative assessment of intake amount, the frequency of consumption occasions was also excluded. |

<sup>1</sup>World Health Organization. Policies to protect children from the harmful impact of food marketing: WHO guideline [Internet]. 2023 [cited 2026 May 26]. Available from: https://www.who.int/publications/i/item/9789240075412

### Data Extraction

Two reviewers independently extracted data using Covidence, following a prospectively developed template published in the review protocol.^26^ To quantify the impact of food marketing on children’s dietary intake, the mean difference (MD) in intakes between intervention and control groups was the primary effect measure extracted for all continuous outcomes, including energy intake, food weight, and item counts. To facilitate the calculation of standard errors and effect sizes, inferential statistics, including t-values and 95% confidence intervals, were extracted for crossover studies, following Cochrane recommendations.^27^ Formulas used in the statistical conversions are available in eTable 4. When necessary, additional data were sought through direct contact with study authors or from related publications in which the authors had previously shared primary results.

### Risk of Bias Assessment and Certainty of Evidence

Two reviewers independently assessed the risk of bias in randomized controlled trials using the Cochrane Risk of Bias 2.0 (RoB 2) and nonrandomized controlled trials using the Risk Of Bias In Non-randomized Studies - of Interventions (ROBINS-I).^28,29^ Disagreements were resolved through team discussions to reach consensus. The Credibility of Effect Modification Analyses (ICEMAN) was used to assess the credibility of potential relevant effect modification.^30^ The Grading of Recommendations, Assessment, Development, and Evaluation (GRADE) framework was applied to determine the overall certainty of the evidence, with final consensus achieved through team discussion.^31^

### Statistical Analysis

The meta-analysis was performed in R (v.4.4.2) using the meta for package with a multivariate random-effects model and inverse-variance method for crossover studies. ^32^ The model accounts for dependence when studies include multiple treatment arms with overlapping participants. An assumed correlation of 0.5 was used for studies lacking t-statistics or CIs. Between-study variance was estimated using the restricted maximum likelihood (REML) estimator. Confidence intervals were calculated using the Wald-type method. Heterogeneity was assessed using Cochran’s Q test, I^2^ statistic, and tau-squared (tau^2^), with I^2^ > 50% indicating substantial heterogeneity. Subgroup differences were tested using Cochran’s Q. Where data were insufficient, a narrative synthesis summarized the evidence.

We assessed robustness by prespecified sensitivity analyses, removing statistical outliers (|z| > 1.96), and using leave-one-out, trim-and-fill, and Cook’s distance methods. Publication bias was evaluated with funnel plots and Egger’s regression. Two-sided tests with P < .05 defined significance. Data is available via Borealis.^33^

### Subgroup Analysis

Subgroup analyses were by age, sex, SEP, weight status, marketing medium, duration of marketing exposure, and food healthfulness. Age groups were <8, 9–12, and 13–19 years, considering the age cutoffs commonly used for dietary reference intake values for nutrients.^34^ Exposure duration was compared as ≤5 vs >5 minutes due to significant heterogeneity in marketing interventions, participant characteristics, and dietary outcome measures.^35^ The 5- minute cutoff followed prior precedent.^19^

## RESULTS

There were 17,488 articles identified from the database search, of which 4807 were duplicates. A total of 12,681 titles and abstracts and 236 potentially relevant full texts were screened for eligibility. Of those, we excluded 228 articles for which reasons for exclusion were recorded (eTable 3a). Overall, we included 39 studies from the two WHO-commissioned reviews,^36–72^ 7 from cross-referencing relevant reviews,^73–77^ and 9 from the database search.^78–85^

### Characteristics of Included Studies

Of 55 eligible studies, 51 were randomized controlled trials (RCTs),^36–40,42–64,67–85^ and 4 were nonrandomized trials.^41,64–66^ Detailed baseline characteristics of the included studies are presented in eTable 5. Among studies reporting mean differences, 47 RCTs included 6,269 total participants aged 2 to 18 years, and 3 nonrandomized trials included 608 total participants aged 4 to 10 years. The sex distribution was relatively balanced, with females comprising 42% to 69% across all studies, except in one study^54^ that included 100% boys. Weight status was reported in 41 studies, with overweight or obesity prevalence varying from 13% to 77.5%.^36–53,55–61,64,67,68,70–72,76–80,82,85^ The included studies were conducted across 17 countries: United Kingdom (n = 15),^40,42–45,58–60,70,74,77–80^ USA (n = 14),^41,46,52,53,61–64,64–66,69,75,85^ the Netherlands (n = 7),^38,39,47–51,82^ Belgium (n = 5),^36,73,76^ Australia (n = 2),^71,72^ Canada (n = 2),^37,54^ France (n = 2),^83^ Germany (n = 1),^84^ South Korea (n = 1),^81^ Spain (n = 1),^51^ India (n = 1),^56^ Italy (n = 1),^57^ Georgia (n = 1),^67^ Chile (n = 1),^68^ and Argentina, Brazil, Mexico (n = 1).^55^

Included studies measured various marketing media: TV (n=24),^37–41,44,46,75,53–61,63,80,67–69,84,85^ digital media (n=14),^42,43,45,47–51,62,72,74,78,79,82^ and food packaging (n=19).^73,81,36,52,55–57,64–66,70,76,77,83^ One study included TV and digital media in a single intervention arm.^71^ Overall, most studies measured the marketing of unhealthy foods,^36–41,43,46,48–61,64,67–73,76,78–81,83,85^ though 9 studies measured both unhealthy and healthy foods,^36,42,44,47,62,63,66,73,75^ and 8 studies only measured the marketing of healthy options.^45,64,65,74,77,82,84^ Reported exposure durations for TV and digital media marketing ranged from 0.75 to 12 minutes. As for comparison groups, the majority (n = 30) were to non-food marketing exposure,^37–40,42–51,53,58–63,69,71,75,78–80,82,84,85^ followed by no marketing exposure (n = 18),^52,54–57,64–68,72,74,77,81,83^ and less food marketing (n = 7).^36,41,70,73,76^ The majority of studies (n=34) reported dietary intakes as changes in energy (kcal/kJ),^37,41–44,46–53,55–57,59,60,63,64,66–69,71,72,75,78–80,83–85^ followed by reporting intakes in grams/ounces (n=20),^73,36,38–40,45,74,54,58,86,62,64,81,70,76,77,82^ and 1 study reported intake in pieces of the tested food.^65^

### Critical Appraisal

GRADE assessment (Table 2) rated the certainty of evidence for M2K effects on dietary intake as low, downgraded for risk of bias and inconsistency. Most RCTs (n=51) showed some risk of bias; reporting bias was a primary concern due to unclear predefined analysis plans. In crossover RCTs, the risk of biases related to period and carryover effects was also noted due to limited data on whether carryover effects had dissipated. Non-RCTs (n=4) had low risk of bias except for potential confounding. Detailed RoB 2 assessments are in eTables 6a-d; traffic-light plots in eFigures 1a-d. Heterogeneity was substantial (I² = 99.5%, p <0.001).

**Table 2.**
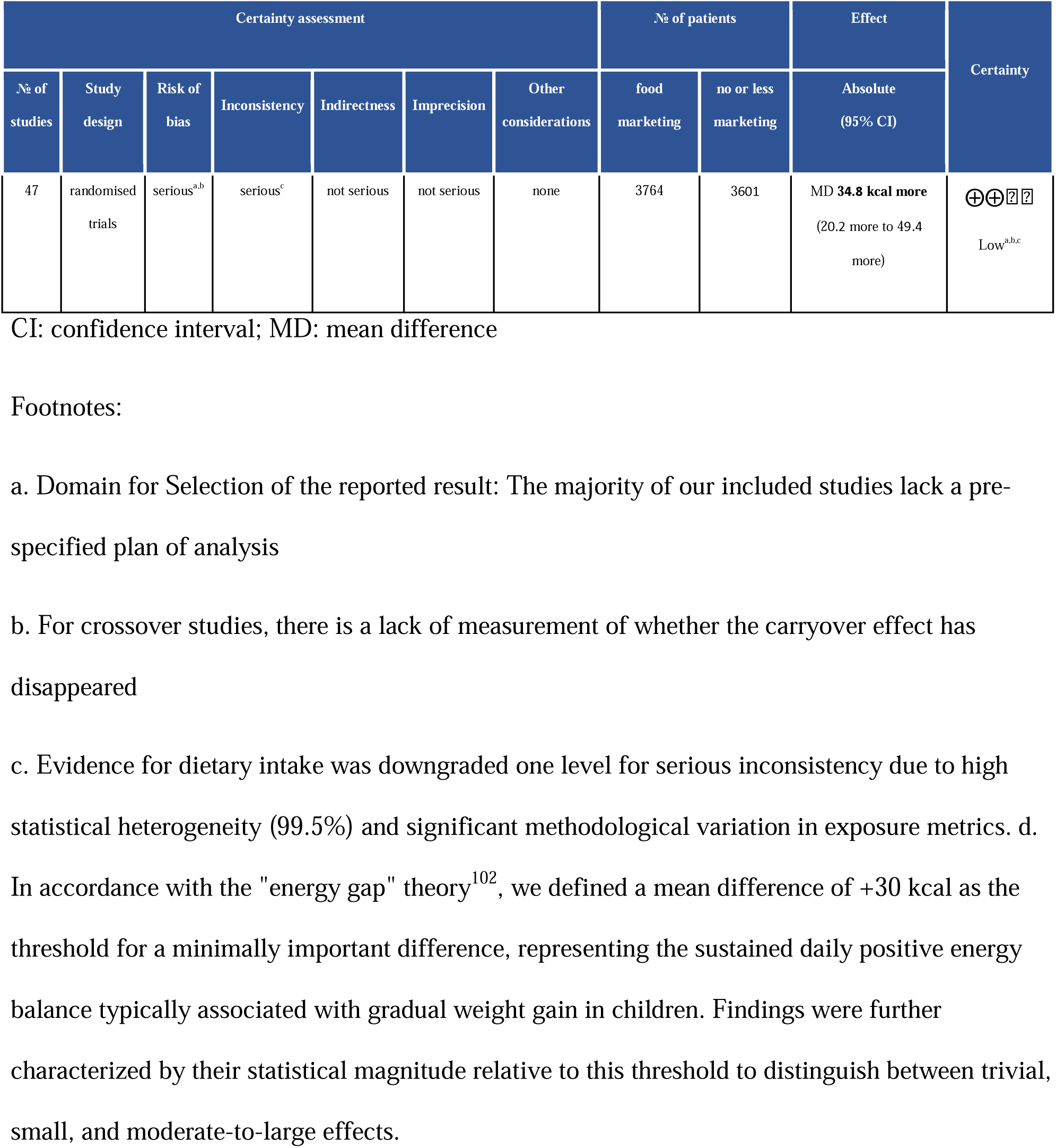
GRADE Summary of Findings.

| Certainty assessment |  |  |  |  |  |  | № of patients |  | Effect | Certainty |
| --- | --- | --- | --- | --- | --- | --- | --- | --- | --- | --- |
| № of studies | Study design | Risk of bias | Inconsistency | Indirectness | Imprecision | Other considerations | food marketing | no or less marketing | Absolute (95% CI) |  |
| 47 | randomised trials | serious<sup>a,b</sup> | serious<sup>c</sup> | not serious | not serious | none | 3764 | 3601 | MD **34.8 kcal more**<br>(20.2 more to 49.4 more) | ⊕⊕⊖⊖<br>Low<sup>a,b,c</sup> |

CI: confidence interval; MD: mean difference
a. Domain for Selection of the reported result: The majority of our included studies lack a pre-specified plan of analysis
b. For crossover studies, there is a lack of measurement of whether the carryover effect has disappeared
c. Evidence for dietary intake was downgraded one level for serious inconsistency due to high statistical heterogeneity (99.5%) and significant methodological variation in exposure metrics. d.
In accordance with the "energy gap" theory<sup>102</sup>, we defined a mean difference of +30 kcal as the threshold for a minimally important difference, representing the sustained daily positive energy balance typically associated with gradual weight gain in children. Findings were further characterized by their statistical magnitude relative to this threshold to distinguish between trivial, small, and moderate-to-large effects.

Subgroup analyses suggest effect modification by marketing medium, sex, age, and weight status, though the certainty of these findings varies. The strongest evidence was for the marketing medium (eTable 7a), with a significant interaction and a large number of trials. Sex (eTable 7b) and age (eTable 7c) showed moderate-to-high credibility. Findings on weight status (eTable 7d) and exposure duration (eTable 7e) were limited by small effects and few studies. These results are exploratory and should be interpreted with caution.

### Meta-Analysis of M2K Effects on Dietary Intakes

Figure 2 presents the overall impact of M2K on dietary intakes of marketing of both healthy and unhealthy foods in 47 RCTs (n=6,269). For the overall effect estimates of studies with separate intervention arms for both healthy and unhealthy food marketing, the unhealthy-arm estimates were used. Pooled data from a random-effects model indicate that food marketing exposure was associated with increased intake (MD, 34.8 kcal; 95% CI, 20.2-49.4), although with substantial heterogeneity. No non-RCTs found significant differences between control and intervention groups.

**Figure 1.**
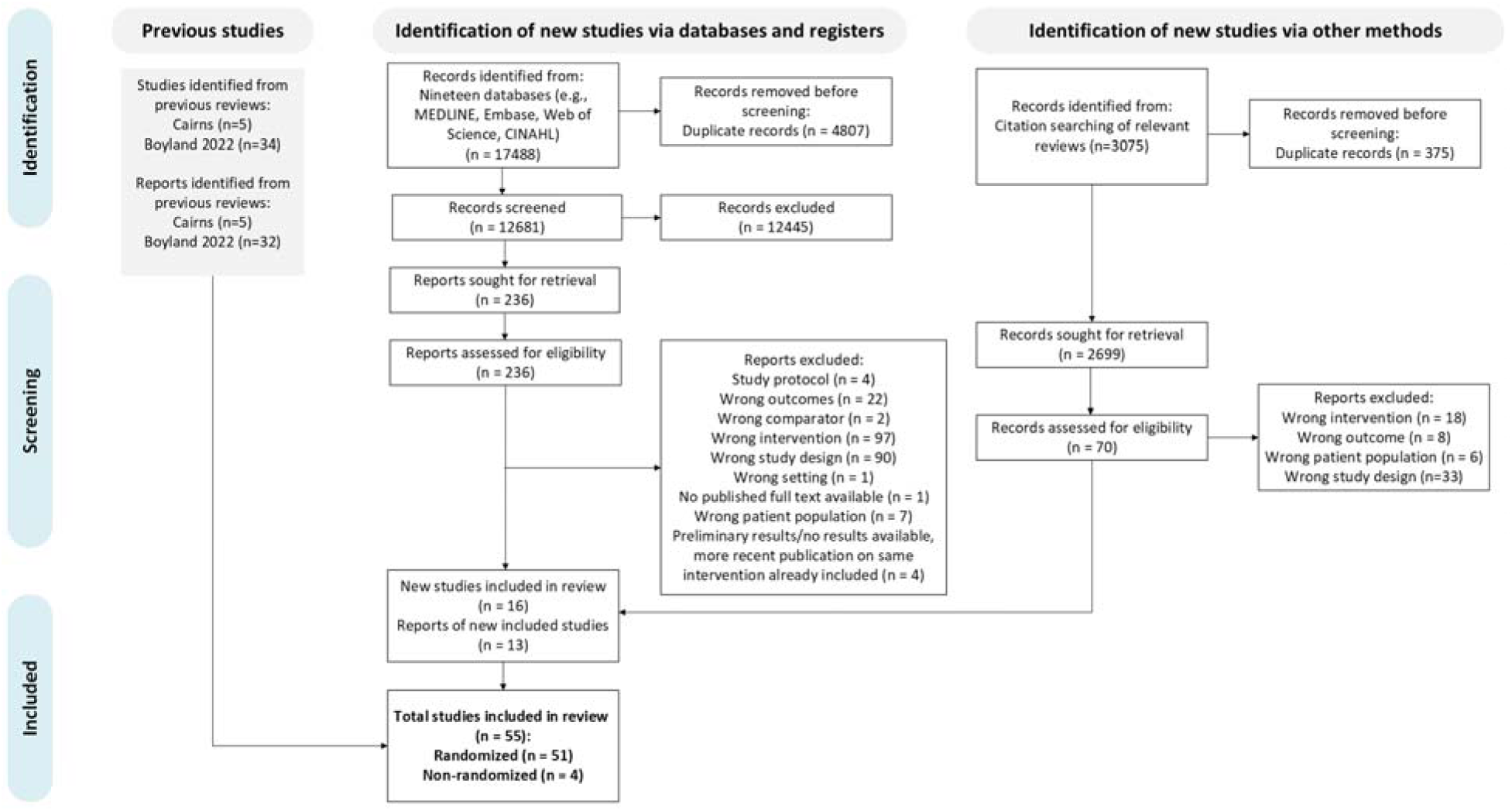
PRISMA Flow Diagram

**Figure 2.**
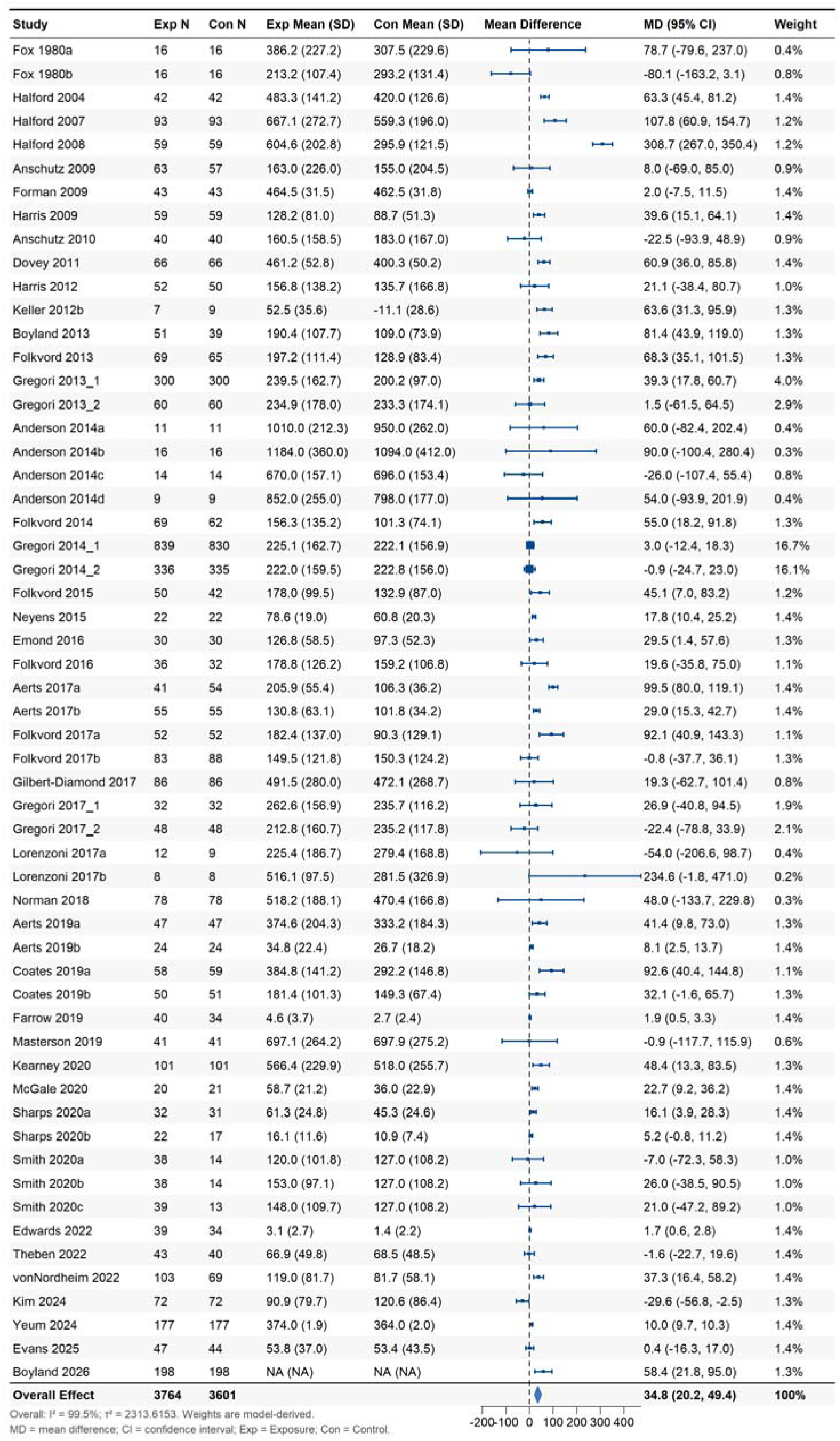
Effect of M2K and Dietary Intakes for Randomized Clinical Trials

### Subgroup Analysis

Figure 3 illustrates the impact of marketing unhealthy foods across media: TV, digital media, and packaging. Digital media (n= 11; MD, 37.5 kcal; 95% CI, 20.1-54.9; I^2^ = 49.0%), packaging (n=11; MD, 20.5 kcal; 95% CI, 0.7-40.3; I^2^ = 86.6%), and TV (n=20; MD, 44.5 kcal; 95% CI, 11.2-77.8; I^2^ = 66.2%) all significantly increased intake. Differences across marketing media were significant regardless of the healthfulness of the marketed food (overall effect, p < .001; marketing of healthy food, p = .017), as shown in eFigure 2-3.

**Figure 3.**
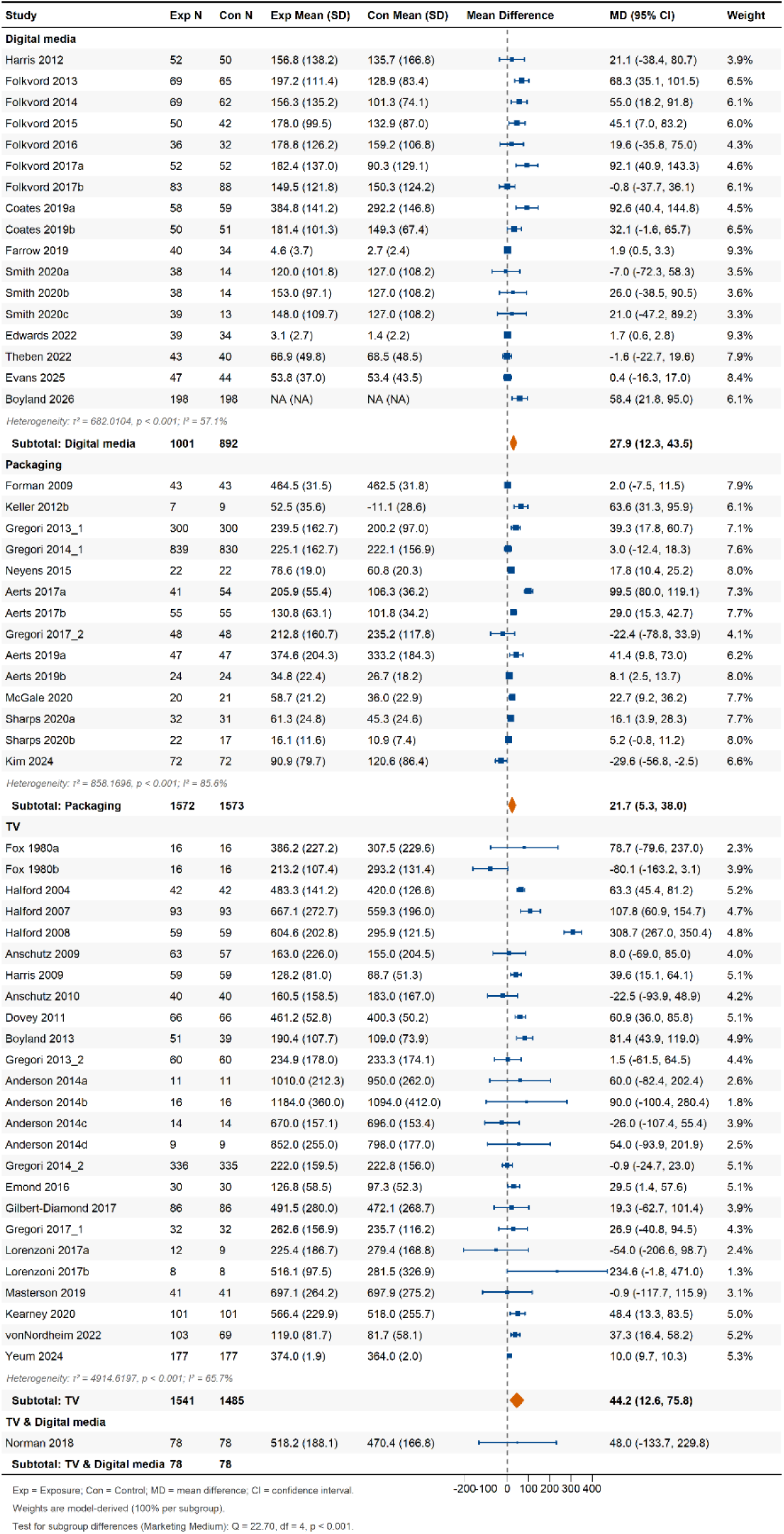
Effect of Unhealthy M2K and Dietary Intakes for Randomized Clinical Trials by Marketing Medium

As shown in eFigure 4, effects varied by age: children under 8 years had an increased energy intake (14 studies; MD, 31.4 kcal; 95% CI, 12.2 to 50.5), and the result for children aged 9-12 years (6 studies) is not significant (MD, 88 kcal; 95% CI, -2.3-178.3). Only one study included adolescents aged 13-18 years (MD, 0.4 kcal; 95% CI, -16.3 to 17.0). Sex differences (3 studies) were observed, with M2K associated with increased energy intake in males (MD, 51.9 kcal; 95%

CI, 45.4-58.3), but not in females (MD, -6.8 kcal; 95% CI, -60.3-46.6) (eFigure 5). People with overweight or obesity were observed to have higher intake (5 studies; MD, 146.9 kcal; 95% CI, 34.1-259.7) after marketing exposure, and there were non-significant impacts on people with normal weight (MD, 55.6 kcal; 95% CI, -51.3-162.5) (eFigure 6). Differences were significant for age (p = .003) and weight status (p = .012) and not for sex (p = .082).

For digital media, energy intake increased for exposures ≤ 5 minutes (9 studies; MD, 44.2 kcal; 95% CI, 27.0 to 61.3) but not for > 5 minutes (2 studies; MD, 1.9 kcal; 95% CI, -14.2 to 17.9). For TV, exposures ≤ 5 minutes (13 studies) showed a non-significant increase (MD, 41.3 kcal; 95% CI, -8.5 to 91.1), while exposures > 5 minutes (3 studies) showed an effect estimate of 10 kcal (95% CI, 9.7 to 10.3). The duration subgroup difference is significant for digital media (p = 0.044) but not TV (p = 0.703) (eFigure 7). Only one study examined the effects by ethnicity, and none reported differences in effect by SEPs.

### Publication Bias and Sensitivity Analysis

Egger’s test indicated significant funnel plot asymmetry (z = 2.055, P = 0.045), but the trim-and-fill analysis found no missing studies (eFigure 8). One study (Halford 2008)^87^ was identified as a statistical outlier (eFigure 9). Removing this outlier yielded a more conservative, but still significant, pooled estimate of 28.63 kcal (95% CI, 19.18-37.62; P < .001), with substantial heterogeneity remaining (I² = 98.5%). There was a marked reduction in τ² (between-study variance) from 2339.67 to 768.50. The removal of the outlier from the subgroup analysis of unhealthy TV marketing resulted in a decreased pooled estimate of 32.4 kcal (95% CI, 14.6-50.2) with a drop in heterogeneity from 66.2% to 24.6%. Sensitivity analysis confirmed the main finding: food marketing increases dietary intake.

## DISCUSSION

This systematic review and meta-analysis incorporated 21 additional studies and 3361 participants not included in the RCTs included in the most recent WHO-commissioned review,^15^ 9 from the database search, 7 from cross-referencing, and 5 from the 2009 WHO-commissioned review.^22^ It compares the effect as a mean difference in energy intake across marketing media for the first time. Our findings show that exposure to food marketing is associated with a small-to-moderate increase in acute dietary intake among children and adolescents. These findings are consistent with the WHO review showing food marketing increases intake in children and adolescents (standardized mean difference, 0.25; 95% CI, 0.15-0.35).^15^ This is the first quantitative comparison across media, indicating unhealthy food marketing via TV, digital media, and packaging increases dietary intake, with the largest effect for TV. These results inform evidence-based M2K policy, especially when resources are limited for comprehensive regulation across marketing media. However, caution is needed to minimize the risk of marketing migrating to other media when only one marketing medium is regulated.^88^

The certainty of evidence from randomized controlled trials was rated as low, mainly due to risk of bias in the selection of the reported result (lack of pre-specified analysis plans) and substantial between-study heterogeneity. In nutrition RCTs, the risk of bias is often rated as “some concerns” in the selective-reporting domain simply because pre-specified analysis plans are missing or vague.^89^ At the same time, empirical work suggests that, for most nutrition trials, the actual analytic choices rarely introduce substantial bias into effect estimates.^90^ Excluding this criterion, most included studies would be rated as low risk of bias, yielding moderate certainty of evidence from GRADE. Additionally, substantial heterogeneity and possible small-study effects were present in the overall effect estimate, indicating that the pooled estimate reflects variation across populations and contexts rather than a single true effect. The heterogeneity became moderate in stratified analyses by food healthfulness for both digital media and TV.

This study assesses how demographic and environmental factors influence children’s dietary outcomes related to commercial exposures. Pooled data suggest boys may be more susceptible to food marketing, leading to higher calorie intake, though based on only three studies. A recent review found mixed evidence, with boys experiencing greater exposure and preference changes, often due to targeted marketing.^19,91^ Further research is needed to explore biological and cultural differences. Children with overweight or obesity also show greater effects, possibly due to increased neurobiological susceptibility.^92^ Evidence on adolescents is limited, though some studies indicate that food marketing increases their consumption of unhealthy foods.^93–97^ Additionally, the impact of food marketing varies by SEP, with limited and mixed findings.^98–101^ More high-quality research, including randomized trials, is needed to understand the health equity implications and to inform effective, equitable policies.

### Limitations

This review is constrained by its focus on imbalanced geographic distributions, acute dietary outcomes measured in controlled experimental settings, and rapid marketing evaluations. Its generalizability is limited by the predominance of studies from high-income countries (91.1%, n = 51) and the scarcity of data from low-income countries. Most studies measured short-term dietary responses in controlled settings, limiting generalizability to real-world, long-term effects. Experimental designs allow precise measurement of intake but may not capture the complexity of children’s typical food environments. Future studies should explore emerging digital platforms, such as influencer marketing and sponsorships, to better understand their impact on dietary behaviors.

## CONCLUSION

This systematic review and meta-analysis of experimental studies demonstrated that exposure to food marketing was associated with increased dietary intake among children and adolescents, with variations based on demographic characteristics and marketing medium. Findings support comprehensive, cross-platform marketing restrictions to protect child health. Future research should focus on rigorously designed, preregistered trials that prioritize socioeconomic and geographic diversity and evaluate effects in adolescent populations.

## Supporting information

PRISMA checklist

## Data Availability

All data produced in the present study are available upon reasonable request to the authors

## OTHER INFORMATION

### Author Contributions

Ms Chen and Dr Jessri had full access to all the data in the study and took responsibility for the integrity of the data and accuracy of the data analysis.

Concept and design: Chen, Mozaffari, Wong, Jessri, Smith, Ananthapavan

Acquisition, analysis, or interpretation of data: All authors.

Drafting of the manuscript: Chen, Jia, Parolin, Wong, Smith, Ananthapavan

Critical revision of the manuscript for important intellectual content: Chen, Wong, Jessri, Smith, Ananthapavan

Statistical analysis: Chen, Wong

Obtained funding: Jessri

Administrative, technical, or material support: Chen, Wong, Jessri

Supervision: Wong, Jessri

