## Supplement for "The Effect of Food and Non-alcoholic Beverage Marketing on Children’s Dietary Intake: A Systematic Review and Meta-Analysis"

**Supplemental Online Content**

**eTable 1.** Search Strategy of the Updated Database Search

**eTable 2.** Relevant Reviews Screened (n=51)

**eTable 3a.** Studies Excluded at the Full Text Screening of the Database Search (n=228)

**eTable 3b.** Studies Excluded at the Full Text Screening of the Citation Search (n=65)

**eTable 4.** Formulas Used in Statistical Conversions

**eTable 5.** Characteristics of Included Studies

**eTable 6a.** Detailed Risk of Bias Assessment - Parallel Studies

**eTable 6b.** Detailed Risk of Bias Assessment - Crossover Studies

**eTable 6c.** Detailed Risk of Bias Assessment - Cluster Studies

**eTable 6d.** Detailed Risk of Bias Assessment – ROBINS-I

**eTable 7a.** Instrument to Assess the Credibility of Effect Modification Analyses (ICEMAN) in a Meta-analysis of Randomized Controlled Trials – Marketing Medium

**eTable 7b.** ICEMAN in a Meta-analysis of Randomized Controlled Trials – Sex

**eTable 7c.** ICEMAN in a Meta-analysis of Randomized Controlled Trials – Age Groups

**eTable 7d.** ICEMAN in a Meta-analysis of Randomized Controlled Trials – Weight Status

**eTable 7e.** ICEMAN in a Meta-analysis of Randomized Controlled Trials – Exposure Duration of Marketing

**eFigure 1a.** Risk of Bias Assessment – Parallel Studies

**eFigure 1b.** Risk of Bias Assessment – Crossover Studies

**eFigure 1c.** Risk of Bias Assessment – Cluster Studies

**eFigure 1d.** Risk of Bias Assessment – ROBINS-I

**eFigure 2.** Effect of M2K on Dietary Intakes by Marketing Medium in Randomized Clinical Trials (RCTs)

**eFigure 3.** Effect of Healthy M2K by Marketing Medium on Dietary Intakes in RCTs

**eFigure 4.** The Effect of Unhealthy M2K on Dietary Intakes by Age Groups in RCTs

**eFigure 5.** The Effect of Unhealthy M2K on Dietary Intakes by Sex in RCTs

**eFigure 6.** The Effect of Unhealthy M2K on Dietary Intakes by Weight Status in RCTs

**eFigure 7.** The Effect of Unhealthy M2K on Dietary Intakes by Exposure Duration in RCTs

**eFigure 8.** Trim-and-fill Analysis: Adjusted Funnel Plot

**eFigure 9.** Diagnostic Analysis: Funnel Plot, Cook’s Distance, and Leave-One-Out Analysis

**eMethod.** Converting Dietary Intakes to Energy Intake in Kilocalories (Kcal)
eTable 1. *Search Strategy of the Updated Database Search*

The search strategies used in the initial database search were published in the review protocol.^1^

Ovid MEDLINE(R) ALL <1946 to March 17, 2026>

Date limit: October 01, 2024 – February 28, 2026

Date searched: March 18, 2026

Results = 1545

1	Beverages/ or Carbonated Beverages/ or Diet/ or Energy Drinks/ or exp Food/ or exp Food Industry/ or "fruit and vegetable juices"/ or exp Milk/ or exp Milk Substitutes/ or sugar-sweetened beverages/ or exp Tea/ or Teas, Herbal/

2	(beverage* or diet* or drink* or food* or nutrition* or snack*).ti,kw,kf.

3	1 or 2

4	Advertising as Topic/ or Direct-to-Consumer Advertising/ or exp Marketing/

5	(adspend* or advert* or advergame* or commercial or commercials or market* or promot* or sponsor*).ti,ab,kw,kf.

6	4 or 5

7	((beverage* or diet* or drink* or fast-food* or food* or nutrition* or snack*) adj3 (adspend* or advert* or advergame* or commercial or commercials or market* or promot* or sponsor*)).ab.

8	(3 and 6) or 7

9	exp Adolescent/ or exp Child/ or exp Child, preschool/ or exp Infant/ or exp Schools/

10	(adolescen* or baby or babies or child* or infant* or pupil* or teen* or "young people" or "young person" or youth*).ti,ab,kw,kf.

11	9 or 10

12	exp Eating/ or Energy Intake/ or Feeding Behavior/ or Food Preferences/

13	(calori* or choice* or consum* or eating behavio* or energ* or favor* or favour* or intake* or liking or prefer* or snacking behavio*).ti,ab,kw,kf.

14	12 or 13

15	8 and 11 and 14

Embase <1974 to 2026 March 16>

Date limit: October 01, 2024 – February 28, 2026

Date searched: March 18, 2026

Results = 3489

1	Diet/ or exp Food/

2	(beverage* or diet* or drink* or food* or nutrition* or snack*).ti,kw,kf.

3	1 or 2

4	exp Advertising/ or Marketing/

5	(adspend* or advert* or advergame* or commercial or commercials or market* or promot* or sponsor*).ti,ab,kw,kf.

6	4 or 5

7	((beverage* or diet* or drink* or fast-food* or food* or nutrition* or snack*) adj3 (adspend* or advert* or advergame* or commercial or commercials or market* or promot* or sponsor*)).ab.

8	(3 and 6) or 7

9	exp Adolescent/ or exp Child/ or exp Schools/

10	(adolescen* or baby or babies or child* or infant* or pupil* or teen* or "young people" or "young person" or youth*).ti,ab,kw,kf.

11	9 or 10

12	exp Eating/ or exp Dietary Intake/ or Feeding Behavior/ or Food Preference/

13	(calori* or choice* or consum* or eating behavio* or energ* or favor* or favour* or intake* or liking or prefer* or snacking behavio*).ti,ab,kw,kf.

14	12 or 13

15	8 and 11 and 14

EBM Reviews - Cochrane Database of Systematic Reviews <2005 to March 11, 2026>

EBM Reviews - Cochrane Central Register of Controlled Trials <January 2026>

Date limit: 2024 – 2026

Date searched: March 18, 2026

Results = 119

1	((beverage* or diet* or drink* or food* or juice* or milk* or nutrition* or snack* or tea*) adj3 (adspend* or advert* or advergame* or commercial or commercials or market* or promot* or sponsor*)).ti,ab,kw,kf.

2	(adolescen* or baby or babies or child* or infant* or pupil* or teen* or "young people" or "young person" or youth*).ti,ab,kw,kf.

3	(calori* or choice* or consum* or eating behavio* or energ* or favor* or favour* or intake* or liking or prefer* or snacking behavio*).ti,ab,kw,kf.

4	1 and 2 and 3

Web of Science

Date limit: October 01, 2024 – February 28, 2026

Date searched: March 18, 2026

Results = 883

#1	TS=( (beverage* OR diet* OR drink* OR food* OR juice* OR milk* OR nutrition* OR snack* OR tea*) NEAR/3 (adspend* OR advert* OR advergame* OR commercial OR commercials OR market* OR promot* OR sponsor*) )

#2	TS=(adolescen* OR baby OR babies OR child* OR infant* OR pupil* OR teen* OR "young people" OR "young person" OR youth* OR schools)

#3	TS=(calori* OR choice* OR consum* OR "eating behavior" OR "feeding behavior" OR "food preferences" OR intake* OR liking OR prefer* OR "snacking behavior" OR "energy intake")

#4	#1 AND #2 AND #3

CINAHL Complete

Date limit: October 01, 2024 – February 28, 2026

Date searched: March 18, 2026

Results = 646

S1	(MH "Beverages") or (MH "Carbonated Beverages") or (MH "Energy Drinks") or (MH "Fruit Juices+") or (MH "Food+") or (MH "Kombucha") or (MH "Milk") or (MH "Milk Substitutes+") or (MH "Sweetened Beverages") or (MH "Tea+") or (MH "Food Industry+") or (MH "Diet") OR (TI(beverage* or diet* or drink* or food* or nutrition* or snack*))

S2	(MH "Marketing+") or ((TI(adspend* or advert* or advergame* or commercial or commercials or market* or promot* or sponsor*)) or (AB(adspend* or advert* or advergame* or commercial or commercials or market* or promot* or sponsor*))

S3	(AB((beverage* or diet* or drink* or fast-food* or food* or nutrition* or snack*) N3 (adspend* or advert* or advergame* or commercial or commercials or market* or promot* or sponsor*)))

S4	S1 AND S2

S5	S3 OR S4

S6	(MH "Adolescence+") or (MH "Child+") or (MH "Schools+") or ((TI(adolescen* or baby or babies or child* or infant* or pupil* or teen* or "young people" or "young person" or youth*)) or (AB(adolescen* or baby or babies or child* or infant* or pupil* or teen* or "young people" or "young person" or youth*)))

S7	(MH "Eating") or (MH "Eating Behavior") or (MH "Energy Intake") or (MH "Food Intake+") or (MH "Food Preferences") or (AB(calori* or choice* or consum* or eating behavio* or energ* or favor* or favour* or intake* or liking or prefer* or snacking behavio*)) or (TI(calori* or choice* or consum* or eating behavio* or energ* or favor* or favour* or intake* or liking or prefer* or snacking behavio*))

S8	S5 AND S6 AND S7

Business Source Ultimate

Date limit: October 01, 2024 – February 28, 2026

Date searched: March 18, 2026

Results = 148

S1	(DE "BEVERAGE industry") OR (DE "FOOD industry")

S2	(TI(beverage* or diet* or drink* or food* or nutrition* or snack*))

S3	S1 OR S2

S4	(DE "ADVERTISING & children") OR (DE "INTERNET advertising & children") OR (DE “ADVERTISING & minorities”) OR (DE "MARKETING")

S5	(TI(adspend* or advert* or advergame* or commercial or commercials or market* or promot* or sponsor*)) or (AB(adspend* or advert* or advergame* or commercial or commercials or market* or promot* or sponsor*)) or (KW(adspend* or advert* or advergame* or commercial or commercials or market* or promot* or sponsor*))

S6	S4 OR S5

S7	(AB((beverage* or diet* or drink* or fast-food* or food* or nutrition* or snack*) N3 (adspend* or advert* or advergame* or commercial or commercials or market* or promot* or sponsor*)))

S8	(S3 AND S6) OR S7

S9	(DE "CHILD consumers") OR (DE "STUDENTS as consumers") OR (DE "TEENAGE consumers") OR (DE "YOUNG consumers") OR (DE “MINORITY consumers”)

S10	(TI(adolescen* or baby or babies or child* or infant* or pupil* or teen* or "young people" or "young person" or youth*)) or (AB(adolescen* or baby or babies or child* or infant* or pupil* or teen* or "young people" or "young person" or youth*)) or (KW(adolescen* or baby or babies or child* or infant* or pupil* or teen* or "young people" or "young person" or youth*))

S11	S9 OR S10

S12	(DE "BEVERAGE consumption statistics") OR (DE "FOOD consumption statistics")

S13	(AB(calori* or choice* or consum* or eating behavio* or energ* or favor* or favour* or intake* or liking or prefer* or snacking behavio*)) OR (TI(calori* or choice* or consum* or eating behavio* or energ* or favor* or favour* or intake* or liking or prefer* or snacking behavio*)) OR (KW(calori* or choice* or consum* or eating behavio* or energ* or favor* or favour* or intake* or liking or prefer* or snacking behavio*))

S14	S12 OR S13

S15	S8 AND S11 AND S14

Communication & Mass Media Complete

Date limit: October 01, 2024 – February 28, 2026

Date searched: March 18, 2026

Results = 6

S1	(TI(beverage* or diet* or drink* or food* or juice* or milk* or nutrition* or snack* or tea*))

S2	(DE "ADVERTISING & children") OR (DE "INTERNET advertising & children") OR (DE “ADVERTISING & minorities”) OR (DE "MARKETING")

S3	(TI(adspend* or advert* or advergame* or commercial or commercials or market* or promot* or sponsor*)) or (AB(adspend* or advert* or advergame* or commercial or commercials or market* or promot* or sponsor*)) or (KW(adspend* or advert* or advergame* or commercial or commercials or market* or promot* or sponsor*))

S4	S1 AND (S2 OR S3)

S5	(DE "JUNK food advertising") OR (DE "FAST food restaurant advertising")

S6	(AB((beverage* or diet* or drink* or fast-food* or food* or nutrition* or snack*) N3 (adspend* or advert* or advergame* or commercial or commercials or market* or promot* or sponsor*)) )

S7	S4 OR S5 OR S6

S8	(TI(adolescen* or baby or babies or child* or infant* or pupil* or teen* or "young people" or "young person" or youth*)) or (AB(adolescen* or baby or babies or child* or infant* or pupil* or teen* or "young people" or "young person" or youth*)) or (KW(adolescen* or baby or babies or child* or infant* or pupil* or teen* or "young people" or "young person" or youth*))

S9	(AB(calori* or choice* or consum* or eating behavio* or energ* or favor* or favour* or intake* or liking or prefer* or snacking behavio*) OR (TI(calori* or choice* or consum* or eating behavio* or energ* or favor* or favour* or intake* or liking or prefer* or snacking behavio*)) OR (KW(calori* or choice* or consum* or eating behavio* or energ* or favor* or favour* or intake* or liking or prefer* or snacking behavio*))

S10	S7 AND S8 AND S9

ERIC

Date limit: October 01, 2024 – February 28, 2026

Date searched: March 18, 2026

Results = 8

S1	(DE “FOOD”)

S2	(TI(beverage* or diet* or drink* or food* or nutrition* or snack*))

S3	S1 OR S2

S4	(DE "ADVERTISING") OR (DE "MARKETING")

S5	(TI(adspend* or advert* or advergame* or commercial or commercials or market* or promot* or sponsor*)) or (AB(adspend* or advert* or advergame* or commercial or commercials or market* or promot* or sponsor*))

S6	S4 OR S5

S7	(AB((beverage* or diet* or drink* or fast-food* or food* or nutrition* or snack*) N3 (adspend* or advert* or advergame* or commercial or commercials or market* or promot* or sponsor*)) )

S8	(S3 AND S6) OR S7

S9	(DE "Young Children") OR (DE "Infants") OR (DE "Preschool Children") OR (DE "Toddlers") OR (DE "Children") OR (DE "Youth") OR (DE "Adolescents") OR (DE "Early Adolescents") OR (DE "Late Adolescents")

S10	(TI(adolescen* or baby or babies or child* or infant* or pupil* or teen* or "young people" or "young person" or youth*)) or (AB(adolescen* or baby or babies or child* or infant* or pupil* or teen* or "young people" or "young person" or youth*))

S11	S9 OR S10

S12	(DE "Eating Habits")

S13	(AB(calori* or choice* or consum* or eating behavio* or energ* or favor* or favour* or intake* or liking or prefer* or snacking behavio*)) OR (TI(calori* or choice* or consum* or eating behavio* or energ* or favor* or favour* or intake* or liking or prefer* or snacking behavio*))

S14	S12 OR S13

S15	S8 AND S11 AND S14

Index to Legal Periodicals and Books (H.W. Wilson)

Date limit: October 01, 2024 – February 28, 2026

Date searched: March 18, 2026

Results = 3

S1	(AB((beverage* OR diet* OR drink* OR food* OR juice* OR milk* OR nutrition* OR snack* OR tea*) N3 (adspend* or advert* or advergame* or commercial or commercials or market* or promot* or sponsor*))) OR (TI((beverage* OR diet* OR drink* OR food* OR juice* OR milk* OR nutrition* OR snack* OR tea*) N3 (adspend* or advert* or advergame* or commercial or commercials or market* or promot* or sponsor*)))

S2	(DE "Teenagers") OR (DE "Students")

S3	(TI(adolescen* or baby or babies or child* or infant* or pupil* or teen* or "young people" or "young person" or youth*)) or (AB(adolescen* or baby or babies or child* or infant* or pupil* or teen* or "young people" or "young person" or youth*))

S4	S2 OR S3

S5	(AB(calori* or choice* or consum* or eating behavio* or energ* or favor* or favour* or intake* or liking or prefer* or snacking behavio*)) OR (TI(calori* or choice* or consum* or eating behavio* or energ* or favor* or favour* or intake* or liking or prefer* or snacking behavio*))

S6	S1 AND S4 AND S5

EconLit

Date limit: October 01, 2024 – February 28, 2026

Date searched: March 18, 2026

Results = 8

((MAINSUBJECT.EXACT("Food; Beverages; Cosmetics; Tobacco; Wine and Spirits (L66)") OR title(beverage* OR diet* OR drink* OR food* OR nutrition* OR snack*) )

AND

(MAINSUBJECT.EXACT("Marketing and Advertising (M3)") OR MAINSUBJECT.EXACT("Advertising (M37)") OR MAINSUBJECT.EXACT("Marketing (M31)") OR MAINSUBJECT.EXACT("Marketing and Advertising: General (M30)") OR abstract(adspend* OR advert* OR advergame* OR commercial OR commercials OR market* OR promot* OR sponsor*) OR title(adspend* OR advert* OR advergame* OR commercial OR commercials OR market* OR promot* OR sponsor*))

OR abstract((beverage* OR diet* OR drink* OR fast-food* OR food* OR nutrition* OR snack*) NEAR/3 (adspend* OR advert* OR advergame* OR commercial OR commercials OR market* OR promot* OR sponsor*)))

AND

( title(adolescen* or baby or babies or child* or infant* or pupil* or teen* or "young people" or "young person" or youth*) or abstract(adolescen* or baby or babies or child* or infant* or pupil* or teen* or "young people" or "young person" or youth*) )

AND

( title(calori* or choice* or consum* or eating behavio* or energ* or favor* or favour* or intake* or liking or prefer* or snacking behavio*) OR abstract(calori* or choice* or consum* or eating behavio* or energ* or favor* or favour* or intake* or liking or prefer* or snacking behavio*) )

Database of Promoting Health Effectiveness Reviews (DoPHER)

Date limit: 2024 –2026

Date searched: March 18, 2026

Results = 26

1	Freetext (All but Author): ("beverage*" or "diet*" or "drink*" or "food*" or "juice*" or "milk*" or "nutrition*" or "snack*" or "tea*")

2	Freetext (All but Author): ("adspend*" or "advert*" or "advergame*" or commercial or commercials or "market*" or "promot*" or "sponsor*")

3	Freetext (All but Author): ("adolescen*" or baby or babies or "child*" or "infant*" or "pupil*" or "teen*" or "young people" or "young person" or "youth*" or schools)

4	Freetext (All but Author): ("calori*" or "choice*" or "consum*" or "eating behavio*" or "energ*" or "favor*" or "favour*" or "intake*" or liking or "prefer*" or "snacking behavio*")

5	1 AND 2 AND 3 AND 4

Global Index Medicus

Date limit: October 2024 – February 2026

Date searched: March 18, 2026

Results = 34

(MH: (G07.203.300* or J02.500*) OR TW: (beverage* or diet* or drink* or food* or nutrition* or snack*))

AND

(MH: (“Marketing” or J01.219.687.274*) OR TW: (adspend* or advert* or advergame* or commercial or commercials or market* or promot* or sponsor*))

AND

(MH: (“Adolescent” or M01.060.406* or M01.060.703*) OR TW: (adolescen* or baby or babies or child* or infant* or pupil* or teen* or "young people" or "young person" or youth*))

AND

(MH: (“Feeding Behavior” or “Energy Intake” or “Food Preferences”) OR TW: (calori* or choice* or consum* or eating behavio* or energ* or favor* or favour* or intake* or liking or prefer* or snacking behavio*))

Health Evidence

Date limit: 2024 – 2026

Date searched: March 18, 2026

Results = 69

(beverage* or diet* or drink* or food* or juice* or milk* or nutrition* or snack* or tea*)

AND

(adspend* or advert* or advergame* or commercial or commercials or market* or promot* or sponsor*)

AND

(adolescen* or baby or babies or child* or infant* or pupil* or teen* or young people or young person or youth* or schools )

AND

(calori* or choice* or consum* or eating behavio* or energ* or favor* or favour* or intake* or liking or prefer* or snacking behavio* )

JSTOR

Date limit: October 01, 2024 – February 28, 2026

Date searched: March 18, 2026

Results = 2

(food* or beverage* or drink* or snack*)

AND

(advert* or market* or promot*)

AND

(adolescen* or child* or infant*)

KoreaMed

Date limit: 2024 –2026

Date searched: March 18, 2026

Results = 27

((((food*[TIAB] OR beverage*[TIAB]) OR snack*[TIAB]) OR drink*[TIAB])

AND

((advert*[TIAB] OR market*[TIAB]) OR promot*[TIAB])

AND

((adolescen*[TIAB] OR child*[TIAB]) OR infant*[TIAB])

Campbell Systematic Reviews

Date limit: October 2024 – February 2026

Date searched: March 18, 2026

Results = 62

"beverage* OR diet* OR drink* OR food* OR juice* OR milk* OR nutrition* OR snack* OR tea*" anywhere

and

"adspend* OR advert* OR advergame* OR commercial OR commercials OR market* OR promot* OR sponsor*" anywhere

and

"adolescen* OR baby OR babies OR child* OR infant* OR pupil* OR teen* OR "young people" OR "young person" OR youth* OR schools" anywhere

and

"calori* OR choice* OR consum* OR "eating behavior" OR "feeding behavior" OR "food preferences" OR intake* OR liking OR prefer* OR "snacking behavior" OR "energy intake"" anywhere

### **eTable 2.** *Relevant Reviews Screened (n=51)*

| Author, Year | Title |
| --- | --- |
| Alghamdi Athir and Bitar 2023 | The positive impact of gamification in imparting nutritional knowledge and combating childhood obesity: A systematic review on the recent solutions |
| Almeida et al. 2024 | Effectiveness of nudge interventions to promote fruit and vegetables' selection, purchase, or consumption: A systematic review |
| Arrona-Cardoza et al. 2023 | The Effects of Food Advertisements on Food Intake and Neural Activity: A Systematic Review and Meta-Analysis of Recent Experimental Studies |
| Boyland 2023 | Is it ethical to advertise unhealthy foods to children? |
| Boyland et al. 2022 | Association of Food and Nonalcoholic Beverage Marketing With Children and Adolescents' Eating Behaviors and Health: A Systematic Review and Meta-analysis |
| Boyland et al. 2022 | Systematic review of the effect of policies to restrict the marketing of foods and non-alcoholic beverages to which children are exposed |
| Chow et al. 2020 | Can games change childrenâ€™s eating behaviour? A review of gamification and serious games |
| Chu et al. 2022 | The impact of food packaging on measured food intake: A systematic review of experimental, field and naturalistic studies |
| Chung et al. 2021 | Adolescent Peer Influence on Eating Behaviors via Social Media: Scoping Review |
| Chung et al. 2022 | Policies to restrict unhealthy food and beverage advertising in outdoor spaces and on publicly owned assets: A scoping review of the literature |
| Coleman et al. 2022 | A rapid review of the evidence for children's TV and online advertisement restrictions to fight obesity |
| Dallagiacoma et al. 2023 | The efficacy of digital media tools to promote a healthy diet in children: A systematic review of intervention studies |
| Delgado et al. 2022 | Unhealthy food advertising. A position paper by the AEP Committee on Nutrition and Breastfeeding |
| Deshpande et al. 2023 | The dark side of advertising: promoting unhealthy food consumption |
| Elliott and Truman 2020 | The Power of Packaging: A Scoping Review and Assessment of Child-Targeted Food Packaging |
| Ertz and Le Bouhart 2021 | The Other Pandemic: A Conceptual Framework and Future Research Directions of Junk Food Marketing to Children and Childhood Obesity |
| Esmaeilpour and Shabani Nashtaee 2024 | Food marketing communication targeting children: A content analysis of research literature (2000â€“2023) |
| Ezike and Da Silva 2023 | Technology-Based Interventions to Reduce Sugar-Sweetened Beverages among Adolescents: A Scoping Review |
| Finlay et al. 2022 | A scoping review of outdoor food marketing: exposure, power and impacts on eating behaviour and health |
| Fischer et al. 2021 | Protecting Our Youth: Support Policy to Combat Health Disparities Fueled by Targeted Food Advertising |
| Folkvord and Hermans 2020 | Food Marketing in an Obesogenic Environment: a Narrative Overview of the Potential of Healthy Food Promotion to Children and Adults |
| Folkvord et al. 2021 | Promoting Fruit and Vegetable Consumption for Childhood Obesity Prevention |
| Gregori et al. 2014 | Randomized Controlled Trials Evaluating Effect of Television Advertising on Food Intake in Children: Why Such a Sensitive Topic is Lacking Top-Level Evidence? |
| Hallez et al. 2020 | That's My Cue to Eat: A Systematic Review of the Persuasiveness of Front-of-Pack Cues on Food Packages for Children vs. Adults |
| Harris and Taillie 2024 | More than a Nuisance: Implications of Food Marketing for Public Health Efforts to Curb Childhood Obesity |
| Hebestreit and Sina 2024 | [Consequences of digital media on the health of children and adolescents with a focus on the consumption of unhealthy foods] |
| Ide et al. 2020 | Priority Actions to Advance Population Sodium Reduction |
| Jena et al. 2023 | Knowledge, practices and influencing factors defining unhealthy food behavior among adolescents in India: a scoping review |
| Kucharczuk et al. 2022 | Social media's influence on adolescents' food choices: A mixed studies systematic literature review |
| Lamas et al. 2023 | The Influence of Serious Games in the Promotion of Healthy Diet and Physical Activity Health: A Systematic Review |
| Lianbiaklal and Rehman 2023 | Revisiting 42 Years of literature on food marketing to children: A morphological analysis |
| Mc Carthy et al. 2022 | The influence of unhealthy food and beverage marketing through social media and advergaming on diet-related outcomes in children-A systematic review |
| Naderer 2020 | Advertising Unhealthy Food to Children: on the Importance of Regulations, Parenting Styles, and Media Literacy |
| Oke and Tan 2022 | Techniques for Advertising Healthy Food in School Settings to Increase Fruit and Vegetable Consumption |
| Omidvar et al. 2021 | Food Marketing to Children in Iran: Regulation that Needs Further Regulation |
| Packer et al. 2022 | The impact on dietary outcomes of licensed and brand equity characters in marketing unhealthy foods to children: A systematic review and meta-analysis |
| Packer et al. 2022 | The Impact on Dietary Outcomes of Celebrities and Influencers in Marketing Unhealthy Foods to Children: A Systematic Review and Meta-Analysis |
| Prowse and Carsley 2021 | Digital Interventions to Promote Healthy Eating in Children: Umbrella Review |
| Prybutok et al. 2024 | Social Media Influences on Dietary Awareness in Children |
| Qutteina et al. 2019 | Media food marketing and eating outcomes among pre-adolescents and adolescents: A systematic review and meta-analysis |
| Russell et al. 2019 | The effect of screen advertising on children's dietary intake: A systematic review and meta-analysis |
| Schneider et al. 2021 | Determinants of soft drink consumption among children and adolescents in developed countries - a systematic review |
| Sina et al. 2022 | Social Media and Children's and Adolescents' Diets: A Systematic Review of the Underlying Social and Physiological Mechanisms |
| Soto Núñez and Martín Salinas 2021 | Analysis of food advertising and its relationship with childhood |
| Suleiman-Martos et al. 2021 | Gamification for the Improvement of Diet, Nutritional Habits, and Body Composition in Children and Adolescents: A Systematic Review and Meta-Analysis |
| Tedstone et al. 2022 | Towards a regulation of food advertising? |
| Tsochantaridou et al. 2023 | Food Advertisement and Dietary Choices in Adolescents: An Overview of Recent Studies |
| Varela et al. 2024 | Bringing down barriers to children's healthy eating: a critical review of opportunities, within a complex food system |
| Villegas-Navas et al. 2020 | The Effects of Foods Embedded in Entertainment Media on Children's Food Choices and Food Intake: A Systematic Review and Meta-Analyses |
| Yoshida-Montezuma et al. 2020 | Does gamification improve fruit and vegetable intake in adolescents? a systematic review |
| Zhang et al. 2020 | Fruit and Vegetable Purchases and Consumption among WIC Participants after the 2009 WIC Food Package Revision: A Systematic Review |

### **eTable 3a.** *Studies Excluded at the Full Text Screening of the Database Search* *(n=228)*

| Author, Year | Title | Exclusion Reason |
| --- | --- | --- |
| Lima et al. 2022 | The influence of nutritional marketing on the preferences, attitudes and consumption of children from 6 to 10 years old | Preliminary results/no results available, more recent publication on same intervention already included |
| Venugopal et al. 2023 | High Fat, Salt and Sugar (HFSS) food consumption and television advertisements: an undesirable association | No published full text available |
| Norman et al. 2018 | Children's self-regulation of eating provides no defense against television and online food marketing | Preliminary results/no results available, more recent publication on same intervention already included |
| Kakwani. 2024 | Teens' Junk Food Consumption Spikes After Watching Gaming App Ads - New Study Reveals. | Preliminary results/no results available, more recent publication on same intervention already included |
| Yamada et al. 2023 | Traditional and modern fast food consumption patterns and associated factors among adolescents and young adults in Hanoi, Vietnam | Preliminary results/no results available, more recent publication on same intervention already included |
| Adams et al. 2019 | Design and rationale for evaluating salad bars and students' fruit and vegetable consumption: A cluster randomized factorial trial with objective assessments. | Study protocol |
| Bestle et al. 2020 | Reducing Young Schoolchildren's Intake of Sugar-Rich Food and Drinks: Study Protocol and Intervention Design for "Are You Too Sweet?" A Multicomponent 3.5-Month Cluster Randomised Family-Based Intervention Study. | Study protocol |
| Melo et al. 2020 | Tailored smartphone intervention to promote healthy eating among Brazilian adolescents: a randomised controlled trial protocol. | Study protocol |
| Hammond et al. 2022 | The Conceptual Framework for the International Food Policy Study: Evaluating the Population-Level Impact of Food Policy. | Study protocol |
| Bragg et al. 2021 | Latino and non-latino white adolescents' preferences for latino-targeted celeb and non-celeb ads | Wrong comparator |
| Ha et al. 2020 | Promoting Resilience to Food Commercials Decreases Susceptibility to Unhealthy Food Decision-Making | Wrong comparator |
| Xing et al. 2022 | [Correlation analysis between children and adolescents watching food TV advertising and fast food consumption]. | Wrong intervention |
| Llauradó et al. 2024 | A 16-month follow-up after a youth-led social marketing intervention to encourage healthy lifestyles in children (aged 9 at baseline and 11 at follow-up) from disadvantaged neighbourhoods: the European Youth Tackling Obesity-Kids project. | Wrong intervention |
| Sutherland et al. 2022 | A cluster randomised controlled trial of a secondary school intervention to reduce intake of sugar-sweetened beverages: Mid-intervention impact of switchURsip environmental strategies. | Wrong intervention |
| Khalil et al. 2020 | A Cross-Sectional Study of Electronic Media Influence on Eating Habits among School Going Adolescents | Wrong intervention |
| Uzsen et al. 2019 | A game-based nutrition education: Teaching healthy eating to primary school students | Wrong intervention |
| Austin et al. 2020 | A Media Literacy-Based Nutrition Program Fosters Parent-Child Food Marketing Discussions, Improves Home Food Environment, and Youth Consumption of Fruits and Vegetables. | Wrong intervention |
| Wengreen et al. 2021 | A randomized controlled trial evaluating the fit game‚Äôs efficacy in increasing fruit and vegetable consumption | Wrong intervention |
| Nasui et al. 2023 | Adolescents' Lifestyle Determinants in Relation to Their Nutritional Status during COVID-19 Pandemic Distance Learning in the North-Western Part of Romania—A Cross-Sectional Study. | Wrong intervention |
| Gesualdo et al. 2019 | Advertising Susceptibility and Youth Preference for and Consumption of Sugar-Sweetened Beverages: Findings from a National Survey. | Wrong intervention |
| Mekonen 2024 | Animal source food consumption and its determinants among children aged 6 to 23 months in sub-Saharan African countries: a multilevel analysis of demographic and health survey. | Wrong intervention |
| Chiong et al. 2022 | Association between perceptions and trust of food advertisements and consumption of Ultra-Processed Foods among US parents and their adolescents | Wrong intervention |
| Tani et al. 2021 | Association of Nursery School-Level Promotion of Vegetable Eating with Caregiver-Reported Vegetable Consumption Behaviours among Preschool Children: A Multilevel Analysis of Japanese Children. | Wrong intervention |
| Chaffee et al. 2021 | Beverage Advertisement Receptivity Associated With Sugary Drink Intake and Harm Perceptions Among California Adolescents. | Wrong intervention |
| Choi et al. 2022 | Caregivers' provision of sweetened fruit-flavoured drinks to young children: importance of perceived product attributes and differences by socio-demographic and behavioural characteristics | Wrong intervention |
| Tan et al. 2024 | Correlates of lifestyle patterns among children in Singapore aged 10 years: the growing up in Singapore towards healthy outcomes (GUSTO) study. | Wrong intervention |
| Kruger et al. 2022 | Decreased frequency of sugar sweetened beverages intake among young children probably linked to the implementation of the health promotion levy in South Africa | Wrong intervention |
| Lebacq et al. 2020 | Determinants of energy drink consumption in adolescents: identification of sex-specific patterns. | Wrong intervention |
| Daly et al. 2023 | Determining the food choice motivations of Irish teens and their association with dietary intakes, using the Food Choice Questionnaire. | Wrong intervention |
| Vasil et al. 2023 | Digital applications as a means for promotion of healthy behaviours among Albanian children. | Wrong intervention |
| Ríos-Reyna et al. 2022 | Efecto de una intervención nutricional en el consumo de alimentos en escolares de educación básica de Reynosa, Tamaulipas, México | Wrong intervention |
| Bestle et al. 2023 | Effectiveness of a Family-based Cluster Randomized Intervention Delivered through School Health Nurses on Reducing Intake of Discretionary Foods and Drinks in Schoolchildren | Wrong intervention |
| Alexandrou et al. 2023 | Effectiveness of a Smartphone App (MINISTOP 2.0) integrated in primary child health care to promote healthy diet and physical activity behaviors and prevent obesity in preschool-aged children: randomized controlled trial. | Wrong intervention |
| Larson et al. 2023 | Effectiveness of the Eggs Make Kids demand-creation campaign at improving household availability of eggs and egg consumption by young children in Nigeria: A quasi-experimental study. | Wrong intervention |
| McGill et al. 2024 | Effectiveness of the Go4Fun program: a comparison of face-to-face and digital delivery | Wrong intervention |
| Shatwan et al. 2023 | Effects of a Smartphone Application on Fruit and Vegetable Consumption Among Saudi Adolescents | Wrong intervention |
| Verjans-Janssen et al. 2020 | Effects of the KEIGAAF intervention on the BMI z-score and energy balance-related behaviors of primary school-aged children. | Wrong intervention |
| Ragelienƒó et al. 2022 | Efficacy of a smartphone application-based intervention for encouraging children's healthy eating in Denmark. | Wrong intervention |
| Gudelj Rakic et al. 2019 | Energy drinks use and relationship with health complaints among Serbian adolescents | Wrong intervention |
| De Droog et al. 2014 | Enhancing children's vegetable consumption using vegetable-promoting picture books. The impact of interactive shared reading and character-product congruence | Wrong intervention |
| Nosi et al. 2021 | Evaluating a social marketing campaign on healthy nutrition and lifestyle among primary-school children: A mixed-method research design. | Wrong intervention |
| Ibeanu et al. 2020 | Evidence-based strategy for prevention of hidden hunger among adolescents in a suburb of Nigeria. | Wrong intervention |
| Gangrade et al. 2023 | Examining the feasibility of a youth advocacy program promoting healthy snacking in New York City: a mixed-methods process evaluation. | Wrong intervention |
| Verma et al. 2023 | Exploring the influence of food labels and advertisements on eating habits of children: a cross-sectional study from Punjab, India. | Wrong intervention |
| Scully et al. 2020 | Factors associated with frequent consumption of fast food among Australian secondary school students. | Wrong intervention |
| Kamar et al. 2016 | Factors influencing adolescent whole grain intake: In-depth interviews with adolescents using SenseCam technology | Wrong intervention |
| Salleh 2019 | Fast food consumption among adolescent and its related factors: Findings from NHMS adolescent nutrition survey 2017 | Wrong intervention |
| Ling et al. 2024 | FirstStep2Health: A cluster randomised trial to promote healthy behaviours and prevent obesity amongst low-income preschoolers. | Wrong intervention |
| Tedstone et al. 2022 | Five years of national policies: progress towards tackling obesity in England. | Wrong intervention |
| Tarroni et al. 2021 | Food and lifestyle education in Tuscan schoolchildren: 2018-2019 follow-up of a long-term campaign...14th European Public Health Conference (Virtual), Public health futures in a changing world, November 10-12, 2021. | Wrong intervention |
| Kuster et al. 2019 | Food packaging cues as vehicles of healthy information: Visions of millennials (early adults and adolescents). | Wrong intervention |
| Ferguson et al. 2021 | Food-Focused Media Literacy for Remotely Acculturating Adolescents and Mothers: A Randomized Controlled Trial of the "JUS Media? Programme". | Wrong intervention |
| Lee et al. 2024 | Food-related media use and eating behavior in different food-related lifestyle groups of Korean adolescents in metropolitan areas | Wrong intervention |
| Jones et al. 2014 | Gamification of dietary decision-making in an elementary-school cafeteria | Wrong intervention |
| Larose et al. 2024 | Healthy Behaviours Promotion Program in Summer Day Camps | Wrong intervention |
| Federici et al. 2024 | Healthy Snack Project: Improving Healthy Choices through Multidisciplinary Food Education Actions. | Wrong intervention |
| Kaar et al. 2020 | High prevalence of obesity-related diet and activity behaviors by age 3 identifies need for early interventions | Wrong intervention |
| Conlon et al. 2019 | Home Environment Factors and Health Behaviors of Low-income, Overweight, and Obese Youth. | Wrong intervention |
| Lawman et al. 2020 | Hydrate Philly: An Intervention to Increase Water Access and Appeal in Recreation Centers. | Wrong intervention |
| Lioutas et al. 2015 | 'I saw Santa drinking soda!' Advertising and children's food preferences | Wrong intervention |
| Bradley et al. 2020 | Impact of a health marketing campaign on sugars intake by children aged 5-11 years and parental views on reducing children's consumption. | Wrong intervention |
| Hager et al. 2023 | Impact of Produce Prescriptions on Diet, Food Security, and Cardiometabolic Health Outcomes: A Multisite Evaluation of 9 Produce Prescription Programs in the United States. | Wrong intervention |
| Steenbock et al. 2019 | Impact of the intervention program "JolinchenKids - fit and healthy in daycare" on energy balance related-behaviors: results of a cluster controlled trial. | Wrong intervention |
| Hamdi et al. 2020 | Implementation of a Multi-Component School Lunch Environmental Change Intervention to Improve Child Fruit and Vegetable Intake: A Mixed-Methods Study. | Wrong intervention |
| Horne et al. 2004 | Increasing children's fruit and vegetable consumption: A peer-modelling and rewards-based intervention | Wrong intervention |
| Garcia-Blanco et al. 2022 | Individual and family predictors of ultra-processed food consumption in Spanish children: The SENDO project | Wrong intervention |
| García-Blanco et al. 2023 | Individual and family predictors of ultra-processed food consumption in Spanish children: The SENDO project | Wrong intervention |
| Jimeno Martinez et al. 2023 | Lifestyle changes after a school-based intervention in children. FLUYE Program | Wrong intervention |
| Cullen et al. 2016 | Meal-Specific Dietary Changes From Squires Quest! II: A Serious Video Game Intervention | Wrong intervention |
| Daas et al. 2024 | Mediators and moderators of the effects of a school-based intervention on adolescents' fruit and vegetable consumption: the HEIA study. | Wrong intervention |
| Daly et al. 2022 | Motivations for food choices in Irish teens from the National Teens' Food Survey II | Wrong intervention |
| Lin et al. 2022 | Multilevel Understanding of the Impact of Individual- and School-Level Determinants on Lipid Profiles in Adolescents: The Cross-Level Interaction of Food Environment and Body Mass Index. | Wrong intervention |
| Duplaga et al. 2021 | Nutritional Behaviors, Health Literacy, and Health Locus of Control of Secondary Schoolers in Southern Poland: A Cross-Sectional Study. | Wrong intervention |
| Wahrenburg et al. 2020 | P62 Impact Evaluation Results of Rethink Your Drink Nevada: A Campaign to Promote Healthful Beverage Choices Among SNAP Households...Society for Nutrition Education and Behavior, 53rd Annual Conference, Virtual Conference, July 20-24, 2020. | Wrong intervention |
| Norman et al. 2019 | Parental support in promoting children's health behaviours and preventing overweight and obesity - a long-term follow-up of the cluster-randomised healthy school start study II trial. | Wrong intervention |
| Ghadirian et al. 2022 | Participatory Video Intervention Increased Critical Nutrition Literacy of Ghanaian Adolescent Girls: A Cluster Randomized Control Trial | Wrong intervention |
| Agarwal et al. 2024 | Perception and Impact of Food and Beverage Marketing on Children's Eating Behaviors and Associated Health Issues | Wrong intervention |
| Gascoyne et al. 2023 | Potential impact of the adult-targeted LiveLighter "Sugary Drinks" campaign on adolescent consumption: Findings from a national cross-sectional school survey. | Wrong intervention |
| Franken et al. 2025 | Promoting water consumption among children through a social network intervention: a cluster randomized controlled trial on a Caribbean island | Wrong intervention |
| Villasana et al. 2020 | Promotion of Healthy Lifestyles to Teenagers with Mobile Devices: A Case Study in Portugal | Wrong intervention |
| Garden et al. 2020 | Relationship between primary school healthy eating and physical activity promoting environments and children's dietary intake, physical activity and weight status: a longitudinal study in the West Midlands, UK. | Wrong intervention |
| Gay et al. 2019 | Role of Organizational Support on Implementation of an Environmental Change Intervention to Improve Child Fruit and Vegetable Intake: a Randomized Cross-Over Design. | Wrong intervention |
| Korn et al. 2021 | Role of social ecological model level on young Pacific children's sugar-sweetened beverage and water intakes: Children's Healthy Living intervention. | Wrong intervention |
| Vonk et al. 2024 | School health promotion and fruit and vegetable consumption in secondary schools: a repeated cross-sectional multilevel study. | Wrong intervention |
| Cole et al. 2019 | Setting Kids Up for Success (SKUFS): Outcomes of an Innovation Project for Promoting Healthy Lifestyles in a Pediatric Patient-Centered Medical Home. | Wrong intervention |
| Liu et al. 2021 | Students' perceptions of school sugar-free, food and exercise environments enhance healthy eating and physical activity | Wrong intervention |
| Cunningham et al. 2024 | Text messages to improve young child diets: Results from a cluster-randomized controlled trial in Kanchanpur, Nepal | Wrong intervention |
| Folkvord et al. 2019 | The effect of a memory-game with images of vegetables on children's vegetable intake: An experimental study. | Wrong intervention |
| Lyu et al. 2022 | The Effect of a Multifaceted Intervention on Dietary Quality in Schoolchildren and the Mediating Effect of Dietary Quality between Intervention and Changes in Adiposity Indicators: A Cluster Randomized Controlled Trial. | Wrong intervention |
| Aygun et al. 2022 | The effect of a school-based fruit and vegetable promotion program on adolescents' fruit and vegetable consumption behavior in Turkey. | Wrong intervention |
| Folkvord et al. 2021 | The Effect of a Serious Health Game on Children‚Äôs Eating Behavior: Cluster-Randomized Controlled Trial | Wrong intervention |
| Hahnraths et al. 2021 | The Effects of the Healthy Primary School of the Future on Children's Fruit and Vegetable Preferences, Familiarity and Intake. | Wrong intervention |
| Fraga et al. 2020 | The habit of buying foods announced on television increases ultra-processed products intake among schoolchildren. | Wrong intervention |
| Hennessy et al. 2023 | The impact of a community social marketing campaign on children's meal orders and consumption: main outcomes from a group randomised controlled trial. | Wrong intervention |
| Moyeenudin et al. 2022 | The Impact of Social Media Applications among Children and Adolescents | Wrong intervention |
| Gowland-Ella et al. 2023 | The outcomes of Thirsty? Choose Water! Determining the effects of a behavioural and an environmental intervention on water and sugar sweetened beverage consumption in adolescents: A randomised controlled trial. | Wrong intervention |
| Mostafavi et al. 2021 | The promotion of healthy breakfast and snacks based on the social marketing model: a mixed-methods study. | Wrong intervention |
| Soares Guimaraes et al. 2023 | The relationship between parent's self-reported exposure to food marketing and child and parental purchasing and consumption outcomes in five countries: findings from the International Food Policy Study. | Wrong intervention |
| Mcgowan et al. 2020 | The start childhood obesity campaign on the island of Ireland: Supporting parents to change behaviours | Wrong intervention |
| Kajons et al. 2023 | Thirsty? Choose Water! A regional perspective to promoting water consumption in secondary school students. | Wrong intervention |
| Gowland-Ella et al. 2022 | Thirsty? Choose Water! Encouraging Secondary School Students to choose water over sugary drinks. A descriptive analysis of intervention components. | Wrong intervention |
| Lundquist et al. 2019 | Time spent looking at food during a delay of gratification task is positively associated with children's consumption at ad libitum laboratory meals. | Wrong intervention |
| Smith et al. 2020 | To play or not to play? The relationship between active video game play and electrophysiological indices of food‚Äêrelated inhibitory control in adolescents | Wrong intervention |
| Harris et al. 2022 | TV exposure, attitudes about targeted food ads and brands, and unhealthy consumption by adolescents: Modeling a hierarchical relationship. | Wrong intervention |
| Lgeret et al. 2022 | Use of Health-Promoting Food and Supplements in Swiss Children. | Wrong intervention |
| Gonzalez et al. 2023 | Using implementation research to improve programs targeting adolescents- experiences from Indonesia and Bangladesh | Wrong intervention |
| Braga-Pontes et al. 2021 | Veggies4myHeart—a digital game to promote vegetable consumption in preschool children...Coimbra Health School Annual Meeting, June 17-19, 2021. | Wrong intervention |
| Kamin et al. 2022 | Water Wins, Communication Matters: School-Based Intervention to Reduce Intake of Sugar-Sweetened Beverages and Increase Intake of Water. | Wrong intervention |
| Vanderlee et al. 2021 | A comparison of self-reported exposure to fast food and sugary drinks marketing among parents of children across five countries. | Wrong outcomes |
| Carroll et al. 2021 | Associations between advertisement-supported media exposure and dietary quality among preschool-age children. | Wrong outcomes |
| De Moraes et al. 2022 | Can Food and Beverage Advertising Questionnaire Predict Overweight and Obesity in Children and Adolescents from Low- and-Middle-Income Countries? | Wrong outcomes |
| Caldwell et al. 2020 | Does Exposure to the Campaign Increase Parental Intentions to Promote More Water and Less Sugar-Sweetened Beverage Consumption? | Wrong outcomes |
| Izadi et al. 2024 | Examining school nutrition policies and their effect on the promotion of low-nutrient foods in the context of sports advertising. | Wrong outcomes |
| Ares et al. 2023 | Exposure effects to unfamiliar food advertisements on YouTube: A randomized controlled trial among adolescents | Wrong outcomes |
| Trubswasser et al. 2022 | Factors Influencing Adolescents' Dietary Behaviors in the School and Home Environment in Addis Ababa, Ethiopia. | Wrong outcomes |
| McKerchar et al. 2020 | Food store environment examination - FoodSee: a new method to study the food store environment using wearable cameras. | Wrong outcomes |
| Hlongwane et al. 2023 | Food-related health challenges of children and the role of the Consumer Protection Act 68 of 2008 in regulating unhealthy food advertising. | Wrong outcomes |
| Boynton-Jarrett et al. 2003 | Impact of television viewing patterns on fruit and vegetable consumption among adolescents | Wrong outcomes |
| Teixeira et al. 2020 | Impacto da mídia no consumo de bebida gaseificada a base de cola | Wrong outcomes |
| Binde et al. 2023 | Influence of advertising on children's food choices | Wrong outcomes |
| McKerchar et al. 2020 | Kids in a Candy Store: An Objective Analysis of Children's Interactions with Food in Convenience Stores. | Wrong outcomes |
| Masterson et al. 2019 | Measurement of external food cue responsiveness in preschool-age children: Preliminary evidence for the use of the external food cue responsiveness scale. | Wrong outcomes |
| Gearhardt et al. 2020 | Neural response to fast food commercials in adolescents predicts intake. | Wrong outcomes |
| Pfender et al. 2023 | Perceptions of Sports and Energy Drinks: Factors Associated with Adolescent Beliefs. | Wrong outcomes |
| Basto-Abreu et al. 2024 | Predicted impact of banning nonessential, energy-dense food and beverages in schools in Mexico: A microsimulation study. | Wrong outcomes |
| Evans et al. 2023 | Recall of food marketing on videogame livestreaming platforms: Associations with adolescent diet-related behaviours and health. | Wrong outcomes |
| Kucuk et al. 2023 | Relationship between Problematic Internet Use and Eating Awareness in Adolescents: A Correlation Study. | Wrong outcomes |
| Domoff et al. 2021 | The association of adolescents' television viewing with Body Mass Index percentile, food addiction, and addictive phone use. | Wrong outcomes |
| Skala et al. 2023 | The impact of marketing activities on children's healthy food choices. | Wrong outcomes |
| Ferreira et al. 2019 | The short- and long-term impact of an incentive intervention on healthier eating: a quasi-experiment in primary- and secondary-school cafeterias in Brazil. | Wrong outcomes |
| Palcu et al. 2019 | Advertising models in the act of eating: How the depiction of different eating phases affects consumption desire and behavior. | Wrong patient population |
| Appleton 2023 | Appearance-based health promotion messages for increasing fruit and vegetable consumption: gender, age and adverse effects. | Wrong patient population |
| Havermans et al. 2024 | Claims highlighting health benefits may promote vegetable consumption: A cross-national comparison of consumer perspectives on the importance of vegetable taste, health, and nutrition | Wrong patient population |
| Caldwell et al. 2020 | Does Exposure to the Choose Water Campaign Increase Parental Intentions to Promote More Water and Less Sugar-Sweetened Beverage Consumption? | Wrong patient population |
| Nayyar et al. 2020 | Does online media self-regulate consumption behavior of INDIAN youth? | Wrong patient population |
| Kay et al. 2024 | Effectiveness of visual nudges for encouraging healthier beverage choices from vending machines. | Wrong patient population |
| Cosgrove et al. 2021 | Predictors of COVID-19-Related Perceived Improvements in Dietary Health: Results from a US Cross-Sectional Study. | Wrong patient population |
| Thomas et al. 2019 | Area deprivation, screen time and consumption of food and drink high in fat salt and sugar (HFSS) in young people: results from a cross-sectional study in the UK. | Wrong setting |
| Adams et al. 2025 | A cluster randomized factorial trial of school-lunch salad bars and marketing on elementary students' objectively measured fruit and vegetable consumption. | Wrong study design |
| Lwin et al. 2020 | A macro-level assessment of introducing children food advertising restrictions on children's unhealthy food cognitions and behaviors. | Wrong study design |
| Pettigrew et al. 2017 | A path analysis model of factors influencing children's requests for unhealthy foods | Wrong study design |
| Ganesamoorthy et al. 2024 | Assessment of screen time and its correlates among adolescents in selected rural areas of Puducherry. | Wrong study design |
| Scully et al. 2012 | Association between food marketing exposure and adolescents' food choices and eating behaviors | Wrong study design |
| Fernandez et al. 2019 | Association between food marketing exposure and consumption of confectioneries among pre-school children in Jakarta | Wrong study design |
| Akter et al. 2025 | Association of sociodemographic factors and television food advertisements with junk food consumption among a sample of adolescents aged 13 to 17 in Magura district, Bangladesh: a cross-sectional survey | Wrong study design |
| Buijzen et al. 2008 | Associations between children's television advertising exposure and their food consumption patterns: A household diary-survey study | Wrong study design |
| Ares et al. 2025 | Associations between exposure to digital food marketing and food consumption in adolescence: a cross-sectional study in an emerging country. | Wrong study design |
| Minaker et al. 2011 | Associations between the perceived presence of vending machines and food and beverage logos in schools and adolescents-diet and weight status | Wrong study design |
| Ellithorpe et al. 2023 | Athletes Drink Gatorade: DMA Advertising Expenditures, Ad Recall, and Athletic Identity Influence Energy and Sports Drink Consumption. | Wrong study design |
| Critchlow et al. 2020 | Awareness of marketing for high fat, salt or sugar foods, and the association with higher weekly consumption among adolescents: a rejoinder to the UK government's consultations on marketing regulation. | Wrong study design |
| Vondikakis et al. 2025 | Behavioral Change Through Serious Gaming Approaches for Childhood Obesity Interventions. | Wrong study design |
| Kostecka et al. 2024 | Beverage Consumption and Factors Influencing the Choice of Beverages among Polish Children Aged 11-13 Years in 2018-2023. | Wrong study design |
| Venegas Hargous et al. 2025 | Changes in Children's Adherence to Sustainable Healthy Diets During the Implementation of Chile's Food Labelling and Advertising Law: A Longitudinal Study (2016-2019). | Wrong study design |
| Fretes et al. 2023 | Changes in children's and adolescents' dietary intake after the implementation of Chile's law of food labeling, advertising and sales in schools: a longitudinal study. | Wrong study design |
| Putnam et al. 2018 | Character Apps for Children's Snacks: Effects of Character Awareness on Snack Selection and Consumption Patterns | Wrong study design |
| Rocha et al. 2021 | Characteristics of the School Food Environment Affect the Consumption of Sugar-Sweetened Beverages Among Adolescents | Wrong study design |
| Tarabashkina et al. 2017 | Children and energy-dense foods – parents, peers, acceptability or advertising? | Wrong study design |
| Hallisky et al. 2025 | Children's satiety responsiveness moderates the association between food reinforcement and eating in the absence of hunger. | Wrong study design |
| Demers-Potvin et al. 2024 | Children's self-reported exposure to sugary beverage advertisements and association with intake across six countries before and during the COVID-19 pandemic: a repeat cross-sectional study | Wrong study design |
| Dalton et al. 2017 | Child-targeted fast-food television advertising exposure is linked with fast-food intake among pre-school children | Wrong study design |
| Kruger et al. 2025 | Decreased frequency of sugar-sweetened beverages intake among young children following the implementation of the health promotion levy in South Africa. | Wrong study design |
| Fallick et al. 2025 | Decreasing Young Children's Sugary Drink Intake through Pediatricians and Social Marketing | Wrong study design |
| Mouse et al. 2026 | Determinants of animal source foods (ASFs) consumption among Somali children aged 6-23 months; evidence from the 2020 Somalia demographic and health survey; a multilevel mixed effect model analysis | Wrong study design |
| Bryl et al. 2025 | Determinants of Diet Quality in Young Football Players from Poznan, Poland. | Wrong study design |
| Park 2024 | Determinants of Sugar-sweetened Beverage Consumption Among Adolescents: A Path Analysis | Wrong study design |
| Díaz-Ramírez et al. 2013 | Effect of the exposure to TV food advertisements on the consumption of foods by mothers and children | Wrong study design |
| Larson et al. 2022 | Effectiveness of the Eggs Make Kids demand-creation campaign at improving household availability of eggs and egg consumption by young children in Nigeria: A quasi-experimental study | Wrong study design |
| Lee et al. 2014 | Effects of exposure to television advertising for energy-dense/nutrient-poor food on children's food intake and obesity in South Korea | Wrong study design |
| Sharps et al. 2016 | Encouraging children to eat more fruit and vegetables: Health vs. descriptive social norm-based messages | Wrong study design |
| Smits et al. 2012 | Endorsing children's appetite for healthy foods: Celebrity versus non-celebrity spokes-characters | Wrong study design |
| Galimov et al. 2019 | Energy drink consumption among German adolescents: Prevalence, correlates, and predictors of initiation. | Wrong study design |
| Hunchangsith et al. 2022 | Estimating the impact of school-based education and restriction on television advertising to prevent childhood obesity in Thailand | Wrong study design |
| Kang et al. 2026 | Evaluation of young children's dietary behaviors by parental growth concern levels in Gyeonggi area: a descriptive study | Wrong study design |
| Vecchio et al. 2019 | Even a very intense exposure to TV advertising promoting fruit consumption is not enough to make children eat more fruit: Results from an experimental study in Italy | Wrong study design |
| Jensen et al. 2021 | Examining Chile's unique food marketing policy: TV advertising and dietary intake in preschool children, a pre- and post- policy study. | Wrong study design |
| Yang et al. 2022 | Excessive Gaming and Online Energy-Drink Marketing Exposure Associated with Energy-Drink Consumption among Adolescents. | Wrong study design |
| Giese et al. 2015 | Exploring the association between television advertising of healthy and unhealthy foods, self-control, and food intake in three European countries | Wrong study design |
| Emond et al. 2019 | Exposure to Child-Directed TV Advertising and Preschoolers' Intake of Advertised Cereals. | Wrong study design |
| Andreyeva et al. 2011 | Exposure to food advertising on television: Associations with children's fast food and soft drink consumption and obesity | Wrong study design |
| Hermans et al. 2018 | Feed the Alien! the Effects of a Nutrition Instruction Game on Children's Nutritional Knowledge and Food Intake | Wrong study design |
| Delfino et al. 2020 | Food advertisements on television and eating habits in adolescents: a school-based study. | Wrong study design |
| NR 2017 | Food and beverage television advertising exposure and youth consumption, body mass index and adiposity outcomes. | Wrong study design |
| Gascoyne et al. 2021 | Food and drink marketing on social media and dietary intake in Australian adolescents: Findings from a cross-sectional survey. | Wrong study design |
| Berhane et al. 2025 | Food environment around schools and adolescent consumption of unhealthy foods in Addis Ababa, Ethiopia. | Wrong study design |
| Qutteina et al. 2022 | Food for teens: how social media is associated with adolescent eating outcomes. | Wrong study design |
| Yan et al. 2022 | Impact of Obesogenic Environments on Sugar-Sweetened Beverage Consumption among Preschoolers: Findings from a Cross-Sectional Survey in Beijing. | Wrong study design |
| Alanazi et al. 2022 | Impact of social media as a risk factor of increased fast-food consumption and increased bad health habits in children and adolescents in Saudi Arabia | Wrong study design |
| Boyland et al. 2021 | Indirect Associations Between Commercial Television Exposure and Child Body Mass Index. | Wrong study design |
| Andrade et al. 2026 | Individual and environmental factors affect the consumption of ultra-processed foods among Brazilian adolescents: results from the National School Health Survey. | Wrong study design |
| Oliveira et al. 2025 | Influence of Family Meals on Nutritional Status, Food Consumption, and Behaviors in Brazilian Children and Caregivers...Society for Nutrition Education and Behavior (SNEB) Conference, July 8-11, 2025, Indianapolis, Indiana | Wrong study design |
| Xian et al. 2021 | Influence of the request and purchase of television advertised foods on dietary intake and obesity among children in China. | Wrong study design |
| Smith et al. 2024 | Influences on the dietary patterns and eating behaviours of 18-36-month-old toddlers in Ireland | Wrong study design |
| Gascoyne et al. 2024 | Is food and drink advertising across various settings associated with dietary behaviours and intake among Australian adolescents? Findings from a national cross-sectional survey | Wrong study design |
| Baldwin et al. 2018 | Like and share: Associations between social media engagement and dietary choices in children | Wrong study design |
| Maximova et al. 2025 | Mitigating child health inequalities through equity, diversity, inclusion, and accessibility school practices in Canada. | Wrong study design |
| Bolton 1983 | Modeling the impact of television food advertising on childrens diets | Wrong study design |
| Ensaff 2018 | Nudging adolescents towards plant-based food choices | Wrong study design |
| Pandey et al. 2025 | Nudging strategies to promote plant-based and sustainable food consumption in canteens. | Wrong study design |
| Florack et al. 2018 | Playing with food: The effects of food pre-exposure on consumption in young children | Wrong study design |
| Ibrahim et al. 2025 | Prevalence of fast food consumption and associated factors among secondary school adolescents in Jigjiga Town Somali Region Eastern Ethiopia. | Wrong study design |
| Lemay et al. 2025 | Promoting Dairy Consumption Among Families: Development and User Experience Study of a Web-Based Nutrition Intervention. | Wrong study design |
| Masserot et al. 2010 | Publicité et obésité enfantine. L'impact des annonces publicitaires télévisées sur les choix alimentaires des enfants | Wrong study design |
| Minaker et al. 2025 | Restaurant marketing to kids in Canada: associations with restaurant consumption and appealing restaurant advertisement features in a nationally representative sample of Canadian young people aged 9-17 years. | Wrong study design |
| Hennessy et al. 2015 | Sugar-Sweetened Beverage Consumption by Adult Caregivers and Their Children: The Role of Drink Features and Advertising Exposure | Wrong study design |
| Harris et al. 2018 | Teaching children about good health? Halo effects in child-directed advertisements for unhealthy food | Wrong study design |
| Kelly et al. 2016 | Television advertising, not viewing, is associated with negative dietary patterns in children | Wrong study design |
| Klepp et al. 2007 | Television viewing and exposure to food-related commercials among European school children, associations with fruit and vegetable intake: A cross sectional study | Wrong study design |
| Kelly et al. 2023 | Testing a conceptual Hierarchy of Effects model of food marketing exposure and associations with children and adolescents' diet-related outcomes. | Wrong study design |
| Vergeer et al. 2025 | The association between exposure to food marketing and dietary intake among youth in six countries. | Wrong study design |
| Liu et al. 2025 | The effect of Internet use on adolescent nutritional outcomes: evidence from China. | Wrong study design |
| Buijzen 2009 | The effectiveness of parental communication in modifying the relation between food advertising and children's consumption behaviour | Wrong study design |
| Chang et al. 2022 | The Effects of a Computer Game (Healthy Rat King) on Preschool Children's Nutritional Knowledge and Junk Food Intake Behavior: Nonrandomized Controlled Trial | Wrong study design |
| Dixon et al. 2007 | The effects of television advertisements for junk food versus nutritious food on children's food attitudes and preferences | Wrong study design |
| Öztürkler Çakir et al. 2023 | The Effects of Time of Watching Television and Food Advertisements on Nutritional Status of Preschool Children | Wrong study design |
| Wang 2025 | The Health Symbols in Western Films and the Dietary Behavior of Chinese Adolescents: The Necessity and Strategies of Health Education Intervention | Wrong study design |
| Bagnato et al. 2023 | The impact of fast food marketing on brand preferences and fast food intake of youth aged 10-17 across six countries. | Wrong study design |
| Silva et al. 2020 | The influence of television on the food habits of schoolchildren and its association with dental caries. | Wrong study design |
| Paquet et al. 2017 | The moderating role of food cue sensitivity in the behavioral response of children to their neighborhood food environment: A cross-sectional study | Wrong study design |
| Fleary et al. 2025 | The Relationship Between Media Food Marketing Influence and Unhealthy Food Intake in Parent–Adolescent Dyads: An Actor–Partner Interdependence Model. | Wrong study design |
| Esmi et al. 2010 | The relationship between watching TV commercials and consumption pattern of Tehran's children and adolescents | Wrong study design |
| Vergeer et al. 2024 | The relationship between youth's exposure to unhealthy digital food marketing and their dietary intake in Canada | Wrong study design |
| Acton et al. 2024 | Trends in food and nutrition behaviours, knowledge and attitudes among youth in six countries: findings from the 2019-2021 International Food Policy Study Youth Surveys. | Wrong study design |
| Jensen et al. 2021 | TV advertising and dietary intake in adolescents: a pre- and post- study of Chile's Food Marketing Policy. | Wrong study design |
| Bacardí-Gascón et al. 2013 | Tv food advertisements' effect on food consumption and adiposity among women and children in Mexico | Wrong study design |
| Rosi et al. 2025 | Unhealthy Ultra-Processed Food Consumption in Children and Adolescents Living in the Mediterranean Area: The DELICIOUS Project. | Wrong study design |
| Chavez-Ugalde et al. 2025 | Using group model building to frame the commercial determinants of dietary behaviour in adolescence - findings from online system mapping workshops with adolescents, policymakers and public health practitioners in the Southwest of England. | Wrong study design |
| Wiecha et al. 2006 | When children eat what they watch: impact of television viewing on dietary intake in youth | Wrong study design |
| Olafsdottir et al. 2014 | Young children's screen habits are associated with consumption of sweetened beverages independently of parental norms | Wrong study design |

### **eTable 3b.** *Studies Excluded at the Full Text Screening of the Citation Search (n=65)*

| Author, Year | Title | Exclusion Reason |
| --- | --- | --- |
| Albert 2017 | #consumingitall: Understanding The Complex Relationship Between Media Consumption And Eating Behaviors | Wrong intervention |
| Angka et al. 2020 | How packaging colours and claims influence children's vegetable attitude and intake – An exploratory cross-cultural comparison between Indonesia and Denmark | Wrong intervention |
| Busse et al. 2017 | Assessing the longitudinal relationship between Peruvian children's TV exposure and unhealthy food consumption | Wrong intervention |
| Franken et al. 2018 | Promoting water consumption on a caribbean island: An intervention using children's social networks at schools | Wrong intervention |
| Haire et al. 2014 | Weight status moderates the relationship between package size and food Intake | Wrong intervention |
| Harris et al. 2022 | TV exposure, attitudes about targeted food ads and brands, and unhealthy consumption by adolescents: Modeling a hierarchical relationship | Wrong intervention |
| Jiang et al. 2016 | "Happy goat says": The effect of a food selection inhibitory control training game of children's response inhibition on eating behavior | Wrong intervention |
| Lwin et al. 2017 | Media exposure and parental mediation on fast-food consumption among children in metropolitan and suburban Indonesian | Wrong intervention |
| Smit et al. 2020 | The Impact of Social Media Influencers on Children's Dietary Behaviors | Wrong intervention |
| Smit et al. 2021 | Promoting water consumption among children: A three-arm cluster randomised controlled trial testing a social network intervention | Wrong intervention |
| Smit et al. 2016 | A social network-based intervention stimulating peer influence on children's self-reported water consumption: A randomized control trial | Wrong intervention |
| Taber et al. 2011 | State policies targeting junk food in schools: Racial/ethnic differences in the effect of policy change on soda consumption | Wrong intervention |
| Thai et al. 2017 | Perceptions of Food Advertising and Association With Consumption of Energy-Dense Nutrient-Poor Foods Among Adolescents in the United States: Results From a National Survey | Wrong intervention |
| Thomas et al. 2019 | Area deprivation, screen time and consumption of food and drink high in fat salt and sugar (HFSS) in young people: Results from a cross-sectional study in the UK | Wrong intervention |
| Van Kleef et al. 2014 | Nudging children towards whole wheat bread: A field experiment on the influence of fun bread roll shape on breakfast consumption | Wrong intervention |
| Vepsäläinen et al. 2022 | A Mobile App to Increase Fruit and Vegetable Acceptance among Finnish and Polish Preschoolers: Randomized Trial | Wrong intervention |
| Wengreen et al. 2013 | Incentivizing Children's Fruit and Vegetable Consumption: Results of a United States Pilot Study of the Food Dudes Program | Wrong intervention |
| Zask et al. 2012 | Tooty Fruity Vegie: An obesity prevention intervention evaluation in Australian preschools | Wrong intervention |
| Beales Iii et al. 2013 | Does advertising on television cause childhood obesity? A longitudinal analysis | Wrong outcomes |
| Belot et al. 2016 | Incentives and children's dietary choices: A field experiment in primary schools | Wrong outcomes |
| Hanks et al. 2016 | Marketing vegetables in elementary school cafeterias to increase uptake | Wrong outcomes |
| Matthes et al. 2015 | Children's consumption behavior in response to food product placements in movies | Wrong outcomes |
| Matthews 2008 | Children and obesity: A pan-European project examining the role of food marketing | Wrong outcomes |
| Ponce-Blandón et al. 2020 | Effects of advertising on food consumption preferences in children | Wrong outcomes |
| Velazquez et al. 2014 | Attention to food and beverage advertisements as measured by eye-tracking technology and the food preferences and choices of youth | Wrong outcomes |
| Galst 1980 | Television Food Commercials and Pro-Nutritional Public Service Announcements as Determinants of Young Children's Snack Choices | Wrong outcomes |
| Alblas et al. 2020 | Food at first sight: Visual attention to palatable food cues on TV and subsequent unhealthy food intake in unsuccessful restrained eaters | Wrong patient population |
| Egbert et al. 2020 | Binge eating, but not dietary restraint, moderates the association between unhealthy food marketing exposure and sugary food consumption | Wrong patient population |
| Kidd et al. 2018 | Junk food advertising moderates the indirect effect of reward sensitivity and food consumption via the urge to eat | Wrong patient population |
| Raghoebar et al. 2019 | Served portion sizes affect later food intake through social consumption norms | Wrong patient population |
| Sharma et al. 2016 | A study on prevalence of fast food intake among urban and semi urban adolescent students of Guwahati. International journal of home | Wrong patient population |
| Pollack et al. 2021 | Twitch user perceptions, attitudes and behaviours in relation to food and beverage marketing on Twitch compared with YouTube | Wrong patient population |
| Andreyeva et al. 2011 | Exposure to food advertising on television: Associations with children's fast food and soft drink consumption and obesity | Wrong study design |
| Bacardí-Gascón et al. 2013 | Tv food advertisements' effect on food consumption and adiposity among women and children in Mexico | Wrong study design |
| Baldwin et al. 2018 | Like and share: Associations between social media engagement and dietary choices in children | Wrong study design |
| Bolton et al. 1983 | Modeling the impact of television food advertising on childrens diets | Wrong study design |
| Buijzen et al. 2009 | The effectiveness of parental communication in modifying the relation between food advertising and children's consumption behaviour | Wrong study design |
| Buijzen et al. 2008 | Associations between children's television advertising exposure and their food consumption patterns: A household diary-survey study | Wrong study design |
| Cassidy et al. 2023 | The impact of racially-targeted food marketing and attentional biases on consumption in Black adolescent females with and without obesity: Pilot data from the Black Adolescent & Entertainment (BAE) study | Wrong study design |
| Cullen et al. 2016 | Meal-Specific Dietary Changes From Squires Quest! II: A Serious Video Game Intervention | Wrong study design |
| DeDroog et al. 2014 | Enhancing children's vegetable consumption using vegetable-promoting picture books. The impact of interactive shared reading and character-product congruence | Wrong study design |
| Díaz-Ramírez et al. 2013 | Effect of the exposure to TV food advertisements on the consumption of foods by mothers and children | Wrong study design |
| Dixon et al. 2007 | The effects of television advertisements for junk food versus nutritious food on children's food attitudes and preferences | Wrong study design |
| Edwards et al. 2022 | Exposure to models' positive facial expressions whilst eating a raw vegetable increases children's acceptance and consumption of the modelled vegetable | Wrong study design |
| Esmi et al. 2010 | The relationship between watching TV commercials and consumption pattern of Tehran's children and adolescents | Wrong study design |
| Florack et al. 2018 | Playing with food: The effects of food pre-exposure on consumption in young children | Wrong study design |
| Folkvord et al. 2020 | The effect of the promotion of vegetables by a social influencer on adolescents' subsequent vegetable intake: A pilot study | Wrong study design |
| Folkvord et al. 2019 | The effect of a memory-game with images of vegetables on children's vegetable intake: An experimental study | Wrong study design |
| Giese et al. 2015 | Exploring the association between television advertising of healthy and unhealthy foods, self-control, and food intake in three European countries | Wrong study design |
| Gketsios et al. 2022 | The Association of Junk Food Consumption with Preadolescents' Environmental Influences: A School-Based Epidemiological Study in Greece | Wrong study design |
| Hennessy et al. 2015 | Sugar-Sweetened Beverage Consumption by Adult Caregivers and Their Children: The Role of Drink Features and Advertising Exposure | Wrong study design |
| Hermans et al. 2018 | Feed the Alien! the Effects of a Nutrition Instruction Game on Children's Nutritional Knowledge and Food Intake | Wrong study design |
| Horne et al. 2004 | Increasing children's fruit and vegetable consumption: A peer-modelling and rewards-based intervention | Wrong study design |
| Jones et al. 2014 | Gamification of dietary decision-making in an elementary-school cafeteria | Wrong study design |
| Klepp et al. 2007 | Television viewing and exposure to food-related commercials among European school children, associations with fruit and vegetable intake: A cross sectional study | Wrong study design |
| Lee et al. 2014 | Effects of exposure to television advertising for energy-dense/nutrient-poor food on children's food intake and obesity in South Korea | Wrong study design |
| Lioutas et al. 2015 | 'I saw Santa drinking soda!' Advertising and children's food preferences | Wrong study design |
| Lwin et al. 2020 | A macro-level assessment of introducing children food advertising restrictions on children's unhealthy food cognitions and behaviors | Wrong study design |
| Norman et al. 2018 | Children's self-regulation of eating provides no defense against television and online food marketing | Wrong study design |
| Paquet et al. 2017 | The moderating role of food cue sensitivity in the behavioral response of children to their neighborhood food environment: A cross-sectional study | Wrong study design |
| Pettigrew et al. 2017 | A path analysis model of factors influencing children's requests for unhealthy foods | Wrong study design |
| Putnam et al. 2018 | Character Apps for Children's Snacks: Effects of Character Awareness on Snack Selection and Consumption Patterns | Wrong study design |
| Sharps et al. 2016 | Encouraging children to eat more fruit and vegetables: Health vs. descriptive social norm-based messages | Wrong study design |
| Tarabashkina et al. 2017 | Children and energy-dense foods: parents, peers, acceptability, or advertising? | Wrong study design |
| Wengreen et al. 2021 | A randomized controlled trial evaluating the fit game’s efficacy in increasing fruit and vegetable consumption | Wrong study design |

### **eTable 4.** *Formulas Used in Statistical Conversions*

| Converting the standard error (SE) to the standard deviation (SD) | SE = $\frac{SD}{\sqrt{n}}$ |
| --- | --- |
| Estimating mean/SD from median/interquartile range (IQR)^1^ | $\bar{x}= \frac{q_{1}+m+q_{3}}{3}$  SD$=\frac{q_{3}-q_{1}}{2\phi^{-1}(\frac{0.75n-0.125}{n+0.25})}$ |
| Estimating mean/SD from max/min^1^ | SD$=\frac{max-min}{2\phi^{-1}(\frac{n-0.375}{n+0.25})}$ |
| Converting t-statistics to the standard error of the treatment effect^2^ | $SE_{md}= \frac{MD}{t}$ |
| Converting f-statistics to t-statistics | T = $\sqrt{f}$ |
| Converting 95% upper limit and lower limit to the standard error of the treatment effect^2^ | $SE_{md}= \frac{Upper Limit-Lower Limit}{3.92}$ |
| Converting SD and the imputed correlation to the standard error of the treatment effect^2^ | $SE_{md}=\sqrt{\frac{{SD}_{1}^{2}}{n}+\frac{{SD}_{2}^{2}}{n}-\frac{2\rho SD_{1}SD_{2}}{n}}$ |

^1^Xiang W, Wenqian W, Jiming L, Tiejun T. Estimating the sample mean and standard deviation from the sample size, median, range and/or interquartile range. BMC medical research methodology. 2014 Dec 19;14. doi:[10.1186/1471-2288-14-135](https://doi.org/10.1186/1471-2288-14-135) PubMed PMID: 25524443.

^2^Higgins JP, Eldridge S, Li T. Chapter 23: Including variants on randomized trials | Cochrane Handbook for Systematic Reviews of Interventions version 6.5 (updated August 2024). In. [cited 2026 Apr 15]. Available from: <https://www.cochrane.org/authors/handbooks-and-manuals/handbook/current/chapter-23#section-23-2>

### **eTable 5.** *Characteristics of Included Studies*

| **Source, y, and country** | **Design** | **No. of participants (% female)** | **Participant Characteristics** | **Intervention** | **Comparison** | **Outcomes** |
| --- | --- | --- | --- | --- | --- | --- |
| Aerts et al, 2017a Belgium | RCT, between-subjects (School) | 95 (48.4) | Age range: 6-7 years  Age mean (SD): NR  Weight status: NR  SEP: NR  Ethnicity: NR | N = 41 Packaging - the larger (60g) of salted or sugared popcorn with a plain package.  Duration: NA | N = 54 Packaging - The regular (30g) of salted or sugared popcorn with a plain package. | During intervention intake of Jimmy’s Popcorn Sweet™ and Jimmy’s Popcorn Salted™. (Grams converted to kcal). Duration: 1h. |
| Aerts et al, 2017b Belgium | RCT, within-subjects (School) | 55 (47.3) | Age range: 3-6 years  Age mean (SD): 4.67 (0.86)  Weight status: NR  SEP: NR  Ethnicity: NR | N = 55 Packaging - **I1**: larger packages of cookies; **I2**: Larger packages of carrots. Duration: NA | N = 55 Packaging - The regular packages of cookies or carrots. | During intervention, ad libitum intake of baby carrots or ladyfinger cookies. (Grams converted to kcal). Duration: 10 mins. |
| Aerts et al, 2019a Belgium | RCT, within-subjects (School) | 47 (61.7) | Age range: 6-8 years  Age mean (SD): 6.6 (0.61)  Weight status: 15% underweight, 70% normal weight, 15% overweight  SEP: NR  Ethnicity: NR | N = 47 Packaging - **I1**: The large-sized portion size of 40 chocolate nuts; **I2**: The large-sized portion size of 40 grapes. Duration: NA | N = 47 Packaging - The regular portion size of 20 chocolate nuts or 20 grapes. | During intervention, ad libitum intake of dark red grapes and chocolate nuts. (Grams converted to kcal). Duration: 10 mins. |
| Aerts et al, 2019b Belgium | RCT, within-subjects (School) | 24 (54.2) | Age range: 5-7 years  Age mean (SD): 5.62 (0.92)  Weight status: 13% underweight, 74% normal weight, 13% overweight  SEP: NR  Ethnicity: NR | N = 24 Packaging - Large-sized spread manipulation depicted a slice of bread with 73 grams of spread. Duration: NA | N = 24 Packaging - Regular-sized spread manipulation depicted a slice of bread with 15 grams of spread. | During the intervention, slices of bread with cheese and chocolate spreads were served ad libitum from jars. (Grams converted to kcal). Duration: 30 mins. |
| Anderson et al, 2014 Canada | RCT, within-subjects (Lab) | 50 (46) | Age range: 9-14 years  Age mean (SD): NR  Weight status: 50% normal weight, 50% overweight/obese  SEP: NR  Ethnicity: NR | N = 50 TV - 30-min TV episode containing 15 food ads shown over four 2-min commercial breaks: 8 fast-food restaurants, 4 candies, 2 breakfast cereals, and 1 orange drink.  Duration: 8 mins | N = 50 TV - 30-min TV episode containing 15 ads that did not have food cues shown over four 2-min commercial breaks. | During the intervention, ad libitum pizza intake. Duration: 30 mins. |
| Anschutz et al, 2009 The Netherlands | RCT, between-subjects (School) | 120 (53.3) | Age range: 8-12 years  Age mean (SD): 9.8 (1.2)  Weight status: 5.9% underweight, 79% normal weight, 10.9% overweight, 4.2% obese  SEP: NR  Ethnicity: NR | N = 63 TV - 3 food commercials (from advertised Verkade cookies, McDonald's, Haribo candy, Dr. Oetker muffins, Kentucky Fried Chicken, and a Dr. Oetker dessert) mixed with 2 neutral commercials (eg, promotion of toys or video games). Duration: NR | N = 57 TV - Solely 5 neutral commercials. | During intervention, ad libitum intake of chocolate-coated peanuts. (Grams converted to kcal). Duration: 20 mins. |
| Anschutz et al, 2010 The Netherlands | RCT, between-subjects (School) | 80 (51.2) | Age range: 8-12 years  Age mean (SD): NR  Weight status: 0.8% underweight, 80.2% normal weight, 17.8% overweight, 0.8% obese  SEP: NR  Ethnicity: NR | N = 40 TV - 20-min movie clip with two 2.5-min commercial breaks each containing four commercials promoting energy-dense foods and one neutral commercial. Duration: 5 mins | N = 40 TV - 20-min movie clip with two 2.5-min commercial breaks, each containing five neutral commercials. | During the intervention, ad libitum intake of chocolate-coated peanuts. (Grams converted to kcal). Duration: 20 mins. |
| Boyland et al, 2013 United Kingdom | RCT, between-subjects (School) | 90 (50.3) | Age range: 8-11 years  Age mean (SD): 10 (0.9)  Weight status: BMI Mean (SD): 19.1 (0.3)  SEP: NR  Ethnicity: NR | N = 51 TV - 20-min cartoon containing a commercial for branded potato chips (Walker’s Ready Salted Crisps) featuring an endorsement by a former England international soccer player. Duration: 0.75 min | N = 39 TV - 20-min cartoon containing one toy commercial. | Post-intervention ad libitum intake of marketed-brand and supermarket-brand potato chips. (Grams converted to kcal). Duration: NR. |
| Boyland et al, 2026 United Kingdom | RCT, within-subjects (School) | 240 (53.3) | Age range: 7-15 years  Age mean (SD): 11 (2)  Weight status: BMI (z-score): 0.56 (1.13)  SEP: NR  Ethnicity: NR | N = 198 digital media - Ten 30-second ads for unhealthy foods.  Duration: 5 mins | N = 198 digital media - Ten 30-second ads for non-food items. | Post-intervention ad-libitum intake of six different unbranded snack and lunch foods, different from the products and brands advertised. Duration: 15 mins. |
| Brown et al, 2017 USA | NRS, between-subjects (NR) | 114 (NR) | Age range: 9-11 years  Age mean (SD): NR  Weight status: 71% normal weight, 12% overweight, 8% obese SEP: % each household income level: 2.7% <$25,000, 10.7% $25–$49,999, 8.0% $50–74,999, 17.0% $75–99,999, 61.6% >$100,000  Ethnicity: 69.3% White, 5.3% Black, 9.6% Hispanic, 15.8% Other | N = 54 TV - 92-min movie ("Alvin and the Chipmunks”) categorized as having a "high dose" frequency of unhealthy and branded food messages. Duration: 2 mins | N = 60 TV - 84-min movie ("Stuart Little”) categorized as having a "low dose" frequency of unhealthy and branded food messages. | Post-intervention ad libitum intake of participants' choice of one of two similar snacks in five categories. Four categories (Cheese snacks, fruit, pretzels, and chocolate) each contained a snack shown in the 'high-dose' movie and an alternative with similar taste and caloric content. One category (Snack cakes) contained two snacks not featured in either movie. Duration: 60 mins. |
| Coates et al, 2019a United Kingdom | RCT, between-subjects (School) | 117 (59.7) | Age range: 9-11 years  Age mean (SD): 10.5 (0.7)  Weight status: 71% normal weight, 18.2% overweight, 10.8% obesity  Ethnicity: **I1**: 85% White, British; **I2**: 83% White, British; **Control**: 88% White, British | N = 58 digital media - **I1**: Profiles consisted of the Instagram banner and 6 images (3 test and 3 filler) of the influencer holding an unhealthy product (e.g., chocolate cookies); **I2**: the influencer holding a healthy product (e.g., banana). Duration: 2 mins | N = 59 Digital media - Profiles consisted of the Instagram banner and 6 images (3 test and 3 filler) of the influencer holding a branded non-food item (control [e.g., sneakers]). | Post-intervention ad-libitum intake of 4 snacks from: (unhealthy: jelly candy and chocolate buttons; and healthy snacks (carrot baton and seedless white grapes) (Only used intake of unhealthy snacks). Duration: 10 mins. |
| Coates et al, 2019b United Kingdom | RCT, between-subjects (School) | 101 (53) | Age range: 9-11 years  Age mean (SD): 10.32 (0.6)  Weight status: 60% normal weight, 32% overweight, 8% obesity  SEP: NR  Ethnicity: NR | N = 50 digital media - A 5-min YouTube video featuring influencer marketing of a branded unhealthy snack (McVitie's chocolate digestives). Duration: 1 min | N = 51 digital media - A 5-min YouTube video featuring influencer marketing of a branded non‐food item (Apple iPhone 8). | Post-intervention ad-libitum intake of two plates of cookies that contained 100 g of McVitie's chocolate digestive cookies, but one was labeled “McVitie's” and the other was falsely labeled “Tesco's”. Duration: 5 mins. |
| Dovey et al, 2011 United Kingdom | RCT, within-subjects (School) | 66 (48.5) | Age range: 5-7 years  Age mean (SD): 6.0 (0.67)  Weight status: 74% normal weight, 17% overweight, 9% obese  SEP: NR  Ethnicity: NR | N = 66 TV - **I1**: 14-min cartoon with 2 mins of unhealthy food ads (McDonald’s, Cadbury’s Creme Egg, Burger King, Mr. Kipling Cakes, and Uncle Ben’s Express Rice). **I2**: 14-min cartoon with 2 min of healthy food adverts (The Co-op Fruit Range, Fruit and Vegetables (NHS 5-a-day), and Innocent Smoothies). Duration: 2 mins | N = 66 TV - 14-min cartoon with 2 mins of toy ads. | Post-intervention ad libitum intake of a low-fat savory snack (Snack-a-Jacks, cheese flavor), low-fat sweet snack (jelly candy), high-fat sweet snack (chocolate), high-fat savory snack (crisps), fruit (green seedless grapes), and vegetable (carrot sticks). Duration: 15 mins. |
| Edwards et al, 2022 United Kingdom | RCT, between-subjects (Home) | 111 (42.3) | Age range: 4-6 years  Age mean (SD): 5.5 (NR)  Weight status: BMI (z-score): 0.2 (-3.99 - 3.7)  SEP: Parental highest educational level achieved: 1.8% GCSE (or equivalent), 12.6% A level (or equivalent), 40.5% undergraduate degree, 44.1% postgraduate qualification, and 0.9% other.  Ethnicity: Parental ethnicity: 93.7% white, 2.7% indian, 3.6% mixed ethnicities. | N = 39 digital media - Video clips of a model facing forward, eating one piece of raw broccoli and displaying a positive facial expression.  Duration: 62 seconds | N = 34 digital media - Video clips of a model putting pens away into a pencil case whilst expressing a neutral facial expression. | Post-intervention ad libitum intake of raw broccoli. Duration: no limit. |
| Emond et al, 2016 USA | RCT, between-subjects (Lab) | 60 (45) | Age range: 2-5 years  Age mean (SD): 4.1 (0.9)  Weight status: 80% normal weight, 20% overweight/obesity  SEP: NR  Ethnicity: 80% non-Hispanic white, 6.7% Hispanic, 13.3% Other | N = 30 TV - 14-min TV segment containing nine 15- or 30-second ads for Bugles corn chips. Duration: 3 mins | N = 30 TV - 14-min TV segment containing six 30-second ads for a national department store. | During intervention, ad libitum intake of marketed (Bugles corn snacks) and non-marketed (Nabisco Teddy Grahams) snacks. Duration: 14 mins. |
| Evans et al, 2025 United Kingdom | RCT, between-subjects (Lab) | 91 (69.2) | Age range: 13-18 years  Age mean (SD): 17.83 (1.48)  Weight status: 64.8% normal weight, 18.7% overweight, 7.7% obesity, 8.8% missing  Ethnicity: 81.3% White, 8.8% Asian, 5.5% Black, 2.2% Arab, 2.2% Mixed | N = 47 digital media - A video game live stream featuring marketing of an HFSS snack (Doritos Lightly Salted crisps). Duration: 7 mins | N = 44 digital media - A video game live stream featuring marketing of a non-food item (Adidas trainers). | Post-intervention ad libitum intake of two bowls of tortilla crisps: one labeled 'Doritos' and the other falsely labeled 'Tesco's. Duration: 5 mins. |
| Farrow et al, 2019 United Kingdom | RCT, between-subjects (School) | 74 (50) | Age range: 3-6 years  Age mean (SD): 4.83 (1.06)  Weight status: NR  SEP: NR  Ethnicity: NR | N = 40 digital media - Maths advergame containing images of real vegetables (Vegetable Maths Masters app). Duration: 10 mins | N = 34 digital media - Maths games not containing images of food (Turtle Maths app). | Post-intervention ad libitum intake of up to 8 pieces each of marketed (corn and carrot) and non-marketed (yellow pepper and cherry tomato) vegetables. (Grams converted to kcal). Duration: no limit. |
| Folkvord et al, 2013 The Netherlands | RCT, between-subjects (School) | 134 (48.5) | Age range: 8-10 years  Age mean (SD): NR  Weight status: 1.9% underweight, 80% normal weight, 15.6% overweight, 2.6% obese  SEP: NR  Ethnicity: NR | N = 69 digital media - **I1**: Advergame promoting a popular candy brand; **I2**: Advergame promoting a popular fruit brand. Duration: 5 mins | N = 65 digital media - Advergame promoting a popular Dutch toy brand. | Post-intervention ad libitum intake of marketed and non-marketed energy-dense snacks (jelly candy and milk-chocolate) and fruit (bananas and apples). Duration: 5 mins. |
| Folkvord et al, 2014 The Netherlands | RCT, between-subjects (School) | 131 (49.8) | Age range: 7-10 years  Age mean (SD): NR  Weight status: 3.8% underweight, 71.3% normal weight, 18.4% overweight, 6.5% obese  SEP: NR  Ethnicity: NR | N = 69 digital media - Advergame promoting a popular candy brand. Duration: 5 mins | N = 62 digital media - Advergame promoting a popular Dutch toy brand. | Post-intervention ad libitum intake of marketed (jelly candy) and non-marketed (milk chocolate) energy-dense snacks. Duration: 5 mins. |
| Folkvord et al, 2015 The Netherlands | RCT, between-subjects (School) | 92 (54.4) | Age range: 7-10 years  Age mean (SD): 8.42 (NR)  Weight status: 6.3% underweight, 75% normal weight, 16.7% overweight, 2.1% obese  SEP: NR  Ethnicity: NR | N = 50 digital media - Advergame promoting a popular candy brand. Duration: 5 mins | N = 42 digital media - Advergame promoting a popular Dutch toy brand. | Post-intervention ad libitum intake of marketed (jelly candy) and non-marketed (milk chocolate) energy-dense snacks. Duration: 5 mins. |
| Folkvord et al, 2016 The Netherlands | RCT, between-subjects (School) | 68 (47) | Age range: 7-10 years  Age mean (SD): 8.9 (1)  Weight status: 3.8% underweight, 73.5% normal weight, 18.9% overweight, 3.8% obese  SEP: NR  Ethnicity: NR | N = 36 digital media - Advergame promoting a popular candy brand. Duration: 5 mins | N = 32 digital media - Advergame promoting a popular Dutch toy brand. | Post-intervention ad libitum intake of marketed (jelly candy) and non-marketed (milk chocolate) energy-dense snacks. Duration: 5 mins. |
| Folkvord et al, 2017a The Netherlands | RCT, between-subjects (School) | 104 (0.5) | Age range: 6-11 years  Age mean (SD): 9 (1.18)  Weight status: 7.1 % underweight, 74.3% normal weight, 13.3% overweight, 5.2% obese  SEP: NR  Ethnicity: NR | N = 52 digital media - Advergame promoting a popular candy brand. Duration: 5 mins | N = 52 digital media - Advergame promoting a popular Dutch toy brand. | During-intervention ad libitum intake of marketed (jelly candy) and non-marketed (milk chocolate) energy-dense snacks. Duration: 5 mins. |
| Folkvord et al, 2017b Spain | RCT, between-subjects (School) | 171 (0.5) | Age range: 6-12 years  Age mean (SD): 8.9 (1.68)  Weight status: 18.5% underweight, 65.5% normal weight, 11.1% overweight, 3.7% obese  SEP: NR  Ethnicity: NR | N = 83 digital media - Advergame promoting a popular candy brand. Duration: 5 mins | N = 88 digital media - Advergame promoting a popular Dutch toy brand. | During-intervention ad libitum intake of marketed (jelly candy) and non-marketed (milk chocolate) energy-dense snacks. Duration: 5 mins. |
| Forman et al, 2009 USA | RCT, within-subjects (Lab) | 43 (60.5) | Age range: 4-6 years  Age mean (SD): 5.9 (0.9)  Weight status: 53% non-overweight, 47% at risk for overweight  SEP: 45% low-income families, $20,000 per year  Ethnicity: 42% African American, 19% Hispanic, 19% Caucasian, 19% other (typically mixed ethnic origin) | N = 43 Packaging - Meals with visible brands on all foods (‘‘branded’’). Duration: NA | N = 43 Packaging - Meals with foods packaged in plain, unrecognizable plastic bags or containers (‘‘unbranded’’). | During intervention, ad libitum intake of Kraft Lunchables Pizzas, Del Monte mixed fruit cups, Nabisco Oreo Cookies, Rold Gold pretzels, Yoplait Trix Raspberry Rainbow/ Strawberry Banana Bash flavored yogurt, Lay's original potato chips, Kool-Aid cherry-flavored Jammers, and Nesquik chocolate milk. Duration: 30 mins. |
| Fox et al, 1980 USA | RCT, between-subjects (Lab) | 96 (50) | Age range: 4-10 years  Age mean: NR  Weight status: NR  SEP: NR  Ethnicity: NR | N = 32 TV - **I1**: a 7-min 50-second morning programming with two ads on Pepsi and two ads on Froot Loops. **I2**: 7-min 50-second morning programming with two ads for carrots and two for milk. Duration: 5 mins | N = 32 TV - Non-food: a 7-min 50-second morning programming with ads for Nerf footballs and basketballs, Silk Silver, and Bonkers. | Post-intervention ad-libitum intake of a tray of foods and beverages: Hershey bars, Fritos, Chips Ahoy cookies, Froot loops, Pepsi, cherry Kool-Aid, cheese, carrots, grapes, apples, milk, and orange juice. Duration: 8 mins. |
| Gilbert-Diamond et al, 2017 USA | RCT, between-subjects (Lab) | 172 (51.2) | Age range: 9-10 years  Age mean: 9.9 (0.6)  Weight status: 76.7% normal weight, 9.3% overweight, 14% obesity  SEP: 2.9% $25 000, 22.1% $25 000–$64 999, 47.1% $65 000–$144 999, 17.4% $145 000–$224 999, 10.5% $225 000  Ethnicity: 86% White, 14% non-white | N = 86 TV - 34-min TV show that contained 7.7 mins of ads for various unhealthy branded foods and 3.1 mins of neutral ads. Duration: 7.7 mins | N = 86 TV - 34-min TV show that included 7.7 mins of toy ads and 3.1 mins of neutral ads. | During intervention ad libitum intake of marketed (Gummy candy) and un-marketed (Cookies, chocolate, and cheese puffs) unhealthy snacks. Duration: 34 mins. |
| Gorn and Goldberg, 1980 Canada | RCT, between-subjects (Boy Scout Organization) | 77 (0) | Age range: 8-10 years  Age mean: NR  Weight status: NR  SEP: NR  Ethnicity: NR | N = 37 TV - One group was exposed to the same ice cream commercial five times, and the other viewed five different commercials for the same brand of ice cream. Duration: 3 mins | N = 40 TV - A control group viewed the "Flintstone" program with no commercial inserts. | Post-intervention ad libitum intake of chocolate or vanilla ice cream in excessive amounts. (No data) Duration: 15 mins. |
| Gregori et al, 2013a Argentina, Brazil, Mexico | RCT, between-subjects (School) | 600 (50) | Age range: 3-10 years  Age mean: NR  Weight status: BMI Median (Q1/3): 16.37 (14.91/18.35)  SEP: "The schools were of a middle socio-economic level."  Ethnicity: 20% Argentina, 60% Brazil, 20% Mexico; | N = 300 Packaging - chocolate was offered with a toy. Duration: NA | N = 300 Packaging - Chocolate was offered without a toy. | During intervention, ad libitum intake of a chocolate-based product mimicking an Easter egg. Duration: 22 mins. |
| Gregori et al, 2013b Argentina, Brazil, Mexico | RCT, between-subjects (School) | 120 (50) |  | N = 60 TV - Approximately 22-min cartoon with three 30-second food ads. Duration: 1.5 mins | N = 60 TV - Approximately 22-min cartoon with no ads. | During intervention, ad libitum intake of up to 12 chocolate eggs (Marketed food). Duration: 22 mins. |
| Gregori et al, 2014a India | RCT, between-subjects (School) | 1669 (50) | Age range: 3-10 years,  Age median (IQR): 6.5 (5; 8)  Weight status: 18% underweight, 59% normal weight, 12% overweight, 11% obese  SEP: Mothers' Education: 3% No one, 5% Elementary school, 10% Middle school, 27% High school, 55% Advanced degree; Fathers' Education: 2% No one, 5% Elementary school, 12% Middle school, 22% High school, 59% Advanced degree; 25% Low SEP | N = 839 Packaging - chocolate was offered with a gadget (toy). Duration: NA | N = 830 Packaging - Chocolate was offered without a toy. | During intervention, ad libitum intake of a chocolate-based product mimicking an Easter egg. Duration: 22 mins. |
| Gregori et al, 2014b India | RCT, between-subjects (School) | 671 (50) |  | N = 336 TV - Approximately 22-min cartoon with three 30-second food ads. Duration: 1.5 mins | N = 335 TV - Approximately 22-min cartoon with no ads. | During intervention, ad libitum intake of up to 12 chocolate eggs (Marketed food). Duration: 22 mins. |
| Gregori et al, 2017a Italy | RCT, between-subjects (Lab) | 64 (50) | Age range: 6-11 years  Age mean (SD): NR  Weight status: BMI Median(Q1/3): 16.88 (15.28/18.75)  SEP: 15% mother unemployed; 100% father unemployed; 2/3/3 number of rooms in the house; 53% have ≥2 cars/vans owned, 2% have no cars/vans owned; 4% have 1 car owned; 41% have 1 van owned  Ethnicity: NR | N = 32 TV - 16-min TV episode containing seven ad segments for various sweet snacks. Duration: 3 mins | N = 32 TV - 16-min TV episode containing no ads. | During intervention, ad libitum intake of marketed sweet snacks. Duration: 16 mins. |
| Gregori et al, 2017b Italy | RCT, between-subjects (Lab) | 96 (50) |  | N = 48 Packaging - 11 snacks with branded packaging were placed on a tray. Duration: NA | N = 48 Packaging - 11 snacks with unbranded packages were placed on a tray. | During the intervention, ad libitum intake of snacks they preferred (cocoa biscuits; soft pastries with apricot jam, milk cream, and chocolate chips; soft sponge with cocoa cream topping and milk cream fillings; mini plum yogurt cakes). Duration: 16 mins. |
| Halford et al, 2004 United Kingdom | RCT, within-subjects (Lab) | 42 (57.1) | Age range: 9-11 years  Age mean (SD): 10.4 (NR)  Weight status: 67% normal weight, 21% overweight, 12% obese  SEP: NR  Ethnicity: NR | N = 42 TV - a collection of food-related ads. Duration: NR | N = 42 TV - A collection of non-food-related ads. | Post-intervention ad libitum intake of Ryvita wholegrain crackers (low-fat savory), Haribo jelly sweets (low-fat sweet), chocolate (high-fat sweet), and butter puffs (high-fat savory). (Grams converted to kcal). Duration: no limit. |
| Halford et al, 2007 United Kingdom | RCT, within-subjects (Lab) | 93 (58.1) | Age range: 5-7 years  Age mean (SD): 6.25 (NR)  Weight status: 70% normal weight, 14% overweight, 16% obese  SEP: NR  Ethnicity: NR | N = 93 TV - a collection of food-related ads. Duration: NR | N = 93 TV - A collection of non-food-related ads. | Post-intervention ad libitum intake of low-fat savory (snack-a-jacks), low-fat sweet (Haribo jelly sweets), high-fat sweet (chocolate buttons), high-fat savory (Walkers’ Ready Salted potato crisps), and fruit (green seedless grapes). Duration: no limit. |
| Halford et al, 2008 United Kingdom | RCT, within-subjects (Lab) | 59 (45.8) | Age range: 9-11 years  Age mean (SD): 10.16 (NR)  Weight status: 56% normal weight, 25% overweight, 19% obese  SEP: NR  Ethnicity: NR | N = 59 TV - a collection of 10 food-related ads: fish fingers (fish sticks), fast-food burger chain, baked beans, chocolate-flavored rice breakfast cereal, fast-food burger chain, oat-based breakfast cereal, white chocolate bar, fruit-flavored sweets, high-fat savory potato snack. Duration: 5 mins | N = 59 TV - A collection of 10 toy ads. | Post-intervention ad libitum intake of low-fat savory (snack-a-jacks, cheese flavor), low-fat sweet (Haribo jelly sweets), high-fat sweet (Cadbury's chocolate buttons), high-fat savory (Walker’s ready salted potato crisps), and low-energy fruit (green seedless grapes).  Duration: no limit. |
| Harris et al, 2009 USA | RCT, between-subjects (School/camp) | 118 (46.8) | Age range: 7-11 years  Age mean: 8.8 (NR)  Weight status: 3% underweight, 62% normal weight, 21% at risk of overweight, 14% overweight  Ethnicity: 95% white, non-Hispanic | N = 59 TV - 14-min cartoon including four 30-second food commercials promoting snack and breakfast foods of poor nutritional quality using a fun and happiness message. Duration: 2 mins | N = 59 TV - 14-min cartoon containing four 30-second non-food commercials (games and entertainment products). | During intervention, ad libitum intake of cheddar cheese and Goldfish crackers. (Grams converted to kcal).  Duration: 14 mins. |
| Harris et al, 2012 USA | RCT, between-subjects (Lab) | 102 (47.4) | Age range: 7-12 years  Age mean: 9.4 (NR)  Weight status: NR  SEP: NR  Ethnicity: NR | **I1**: N = 52 digital media - 2 advergames promoting sweet snack foods (Pop-Tarts and Oreo cookies); **I2**: N = 47 digital media - 2 advergames promoting fruit and vegetables (Dole Foods). Duration: 12 mins | N = 50 digital media - Online games with non-food subjects (Jewel Quest and Tumblebugs). | Post-intervention ad-libitum intake of six snack foods, ranging from very healthy (carrots and grapes), to somewhat unhealthy (fruit snacks and Goldfish crackers), to very unhealthy (potato chips and chocolate chip cookies). (Grams converted to kcal). Duration: 20 mins. |
| Jeffrey et al, 1982 USA | RCT, between-subjects (Lab) | 47 (NR) | Age range: 4-5 years  Age mean: NR  Weight status: NR  SEP: NR  Ethnicity: NR | N = NR TV – **I1**: Low Nutrition: 12-min children's program with 3 different 30-second ads for Pepsi, Fritos, and Hershey Chocolate that were shown twice in a cartoon segment. **I2**: 12-minute children's programming with 3 different 30-second ads for grapes, milk, and cheese that were shown twice in a cartoon segment.  Duration: 1.5 mins | N = NR TV - Non-food control: 12-min children's program with 3 different 30-second ads for 3 toys that were shown twice in a cartoon segment. | Post-intervention ad libitum intake of 6 pro-nutrition foods and 6 low-nutrition foods. (No data). Duration: 8 mins. |
| Kearney et al, 2020 United Kingdom | RCT, within-subjects (School) | 101 (60.4) | Age range: 8-10 years  Age mean: 9·86 (0·54)  Weight status: 77% normal weight, 22% overweight/obesity  SEP: Quintile 1 (least deprived) = 2 (2·0%); Quintile 2= 3 (3·0%); Quintile 3= 6 (5·9%); Quintile 4= 53 (52·5%); Quintile 5 (most deprived) = 37 (36·6%)  Ethnicity: NR | N = 101 TV - 21-min cartoon episode containing four ~30-second food ads promoting high-sugar food and beverage items (snacks, confectionery, and sugar-sweetened beverages).  Duration: 2 mins | N = 101 TV - 21-min cartoon episode containing four ~30-second toy ads. | Post-intervention ad libitum intake of unbranded unhealthy (Chocolate buttons, jelly sweets, orange juice) and healthy (Grapes, carrot sticks, water) snack foods and beverages (Assume not advertised). Duration: 15 mins. |
| Keller et al, 2012a USA | NRS, between-subjects (Lab) | 41 (49) | Age range: 7-9 years  Age mean (SD): 8.4 (0.5)  Weight status: 54% non-overweight, 46% at risk for overweight  SEP: NR  Ethnicity: 33% African-American, 32% Hispanic-American | N = 41 Packaging - Multi-item test-meal that was “branded” with the logo of a popular fast-food restaurant. Duration: NA | N = 41 Packaging - Multi-item test-meal that was “unbranded” without fast food logos. | During the intervention, ad libitum intake of test meals included turkey and cheese, ham and cheese, and peanut butter and jelly sandwiches; pretzels, graham crackers, apple slices, carrot sticks, pudding, plain and chocolate milks. Duration: 30 mins. |
| Keller et al, 2012b USA | RCT, between-subjects (Lab) | 16 (NR) | Age range: 4-5 years  Age mean: NR  Weight status: NR  SEP: NR  Ethnicity: NR | N = 7 Packaging - Children received fruits and vegetables in cartoon-decorated containers with collectible reward stickers in weeks 3-6, while baseline and follow-up weeks used plain containers. Duration: NA | N = 9 Packaging - The control group received F&V in plain plastic containers throughout the study. | During intervention, ad libitum intake of eight-ounce plastic containers filled with the following F&V: beets, broccoli, carrots, red peppers, pineapple, and blueberries. (Grams converted to kcal). Duration: NA. |
| Kim et al, 2024 South Korea | RCT, between-subjects (Kindergarten) | 144 (54.9) | Age range: 3-6 years  Age mean: 5.1 (0.7)  Weight status: NR  SEP: NR  Ethnicity: NR | N = 72 Packaging - The experimenter introduced the cookie to the children as “Kiki”. Duration: NA | N = 72 Packaging - The experimenter introduced the cookie to the children as "Cookie". | Post-intervention ad libitum intake of cookies. (Grams converted to kcal). Duration: NA. |
| Kotler et al, 2012 USA | NRS, between-subjects (School) | 343 (51) | Age range: 2-6 years  Age mean: 4.08 (0.99)  Weight status: NR  SEP: NR  Ethnicity: 42% White, 35% African American, 11% Latino, 5% Asian American, 7% multiracial | N = NR Packaging - Target Food with a familiar character (Elmo from Sesame Street) placed in front of the first food in the pair. Duration: NA | N = NR Packaging - The foods with no character cut-out placed in front of the first food in the pair. | During intervention, ad libitum intake of zucchini vs. celery, grapes vs. banana, and chocolate vs. broccoli. (No data) Duration: NA. |
| Leonard et al, 2019a USA | NRS, between-subjects (Lab) | 58 (45) | Age range: 4-7 years  Age mean: 5.8 (NR)  Weight status: NR  SEP: NR  Ethnicity: NR | N = 28 Packaging - Children were seated and viewed a package of cookies with a Scooby Doo image. Duration: NA | N = 30 Packaging - Children were seated and viewed a package of cookies without Scooby Doo. | During intervention, ad libitum intake of a bowl of cookies. Duration: 3 mins. |
| Leonard et al, 2019b USA | NRS, between-subjects (Lab) | 52 (45) |  | N = 27 Packaging - Children were seated and viewed a package of dried apricots with a Scooby Doo image. Duration: NA | N = 25 Packaging - Children were seated and viewed a package of dried apricots without Scooby Doo. | During intervention, ad libitum intake of a bowl of dried apricots. Duration: 3 mins. |
| Lorenzoni et al, 2017a Georgia | RCT, between-subjects (School) | 21 (51) | Age range: 3-11 years,  Age median (IQR): 6 (5;9)  Weight status: 54% normal weight, 28% overweight, 18% obese  SEP: NR  Ethnicity: NR | N = 12 TV - Approximately 22-min cartoon with three 30-second food ads.  Duration: 1.5 min | N = 9 TV - Approximately 22-min cartoon with no ads. | During intervention, ad libitum intake of up to 12 chocolate eggs (Marketed food). Duration: 22 mins. |
| Lorenzoni et al, 2017b Chile | RCT, between-subjects (School) | 16 (50) | Age range: 6-12 years  Age mean (SD): 9.5 (NR)  Weight status: 2.5% underweight, 20% normal weight, 40% overweight, 37.5% obese  SEP: NR  Ethnicity: 12.8% Mapuche, 12.8% Mestizo, 74.3% white | N = 8 TV - 22-min cartoon with three food ads. Duration: NR | N = 8 TV - 22-min cartoon with no ads. | During the intervention, ad libitum intake of 10 different sweet snacks was provided in their branded packaging. Duration: 22 mins. |
| Masterson et al, 2019 USA | RCT, within-subjects (Lab) | 41 (46.3) | Age range: 7-9 years  Age mean: 7.90 (0.70)  Weight status: 61% normal weight, 24.5% overweight, 14.5% obesity  SEP: Primarily college-educated parents with relatively high household incomes ($76,000 – $100,000/year)  Ethnicity: 82.9% Caucasian, 9.8% Black, 4.9% Asian, 2.4% Hispanic | N = 41 TV - 12-min cartoon containing ten food commercials presented in two 90-second commercial breaks. Commercials were selected using Nielsen data (Nielsen Ad Intel, 2015) for the most-advertised brands seen by children. Duration: 3 mins | N = 41 TV - 12-min cartoon with ten toy commercials presented in two 90-second commercial breaks. | Post-intervention ad libitum intake of three high-energy dense foods (Pizza, chocolate chip cookies, French Fries) and three low-energy dense foods (Grilled chicken, broccoli, red grapes). Duration: 30 mins. |
| McGale et al, 2020 United Kingdom | RCT, between-subjects (School) | 41 (46.3) | Age range: 7-11 years  Age mean: 9.0 (1.5)  Weight status: 82.9% normal weight, 17.1% overweight/obese  SEP: Parental education level: 0% post-graduate, 7.3% degree, 36.6% A levels, 14.6% GCSE, 19.5% other, 22% undisclosed  Ethnicity: 68.3% British White, 9.7% British Other; 2.4% Mixed Other; 17.1% undisclosed | N = 20 Packaging - The cereal box with bowl contained a larger portion visual cue depicted on the front (three times the recommended serving: 90g). Duration: NA | N = 21 Packaging - The cereal box with bowl contained a small portion visual cue depicted on the front (same as the written gram serving on pack: 30g). | Post-intervention ad libitum intake of cereal (Kellogg's Corn Flakes). (Grams converted to kcal). Duration: no limit. |
| Neyens et al, 2015 Belgium | RCT, within-subjects (School) | 22 (54.5) | Age range: 4-5 years  Age mean: 4.36 (0.49)  Weight status: 18% overweight  SEP: NR  Ethnicity: NR | N = 22 Packaging - cereal packages with a large portion-size image. Duration: NA | N = 22 Packaging - Cereal packages with a small portion-size image. | Post-intervention ad libitum intake of Crownfield's Frosted Flakes™ and Crownfield's regular Corn Flakes™. (Grams converted to kcal). Duration: 30 mins. |
| Norman et al, 2018 Australia | RCT, within-subjects (Camp) | 78 (50) | Age range: 7-12 years  Age mean: 9.3 (1.6)  Weight status: 3.3% underweight, 80.5% normal weight, 12.3% overweight, 3.9% obese  SEP: Median household income: $2000–2499 per week (substantially higher than the NSW median household income of $800–999 per week)  Ethnicity: NR | N = 78 TV & advergame - 10-min cartoon containing 10 approx. 30-second unhealthy food ads, followed by 5 mins playing an iPad advergame featuring an unhealthy food brand. Advertised foods were classified as high in fat, salt, and/or sugar in accordance with Food Standards Australia New Zealand's nutrient profiling criteria.  Duration: 5 mins | N = 78 TV & advergame - 10-min cartoon containing 10 approx. 30-second non-food ads, followed by 5 mins playing an iPad advergame featuring a non-food brand. | Ad libitum intake of snacks: high-fat savory (crinkle-cut crisps, plain potato crisps, or chicken-flavored crackers); low-fat savory (pretzels, plain crackers, or rice crackers); high-fat sweet (milk chocolate, chocolate-covered biscuits, or sugar-coated chocolate confectionery); low-fat sweet (assorted jelly lollies); fruit (green and black grapes or peeled mandarin segments); and vegetable (carrot sticks). Duration: 15 mins. |
| Sharps et al, 2020a United Kingdom | RCT, between-subjects (School) | 63 (60.3) | Age range: 6-11 years  Age mean (SD): 8.9 (1.41)  Weight status: 73% normal weight  NR | N = 32 Packaging - Laminated photographic image of green grapes placed on the given plastic white plate. Duration: NA | N = 31 Packaging - No image presented on the given plastic white plate. | During the intervention, ad libitum intake of a bowl of grapes. (Grams converted to kcal). Duration: 7 mins. |
| Sharps et al, 2020b United Kingdom | RCT, between-subjects (Family Science Event) | 39 (52.5) | Age range: 5-13 years  Age mean (SD): 8.57 (2.13)  Weight status: 85% normal weight  SEP: NR  Ethnicity: NR | N = 22 Packaging - A plate contained a laminated photographic image of a large portion of carrots. Duration: NA | N = 17 Packaging - No image presented on the plate. | Post-intervention ad libitum intake of a bowl of carrots. (Grams converted to kcal). Duration: 7 mins. |
| Smith et al, 2020a Australia | RCT, between-subjects (Lab) | 52 (45) | Age range: 7-12 years  Age mean (SD): 8.7 (1.5)  Weight status: 9% underweight, 62% normal weight, 15% overweight, 13% obesity  SEP: NR  Ethnicity: NR | N = 38 digital media - Web-based game in which the player is instructed to collect as many coins as possible before time runs out, with a rectangular banner ad for the test brand directly beneath the game onscreen. Duration: 4 mins | N= 41 digital media - Game with no advertising techniques (no instances of the test brand). Participants instructed to collect as many coins as they could before the time finished. | Post-intervention ad libitum intake of one snack from a selection of four items (green grapes and three types of Gummy Lollies, including the marketed brand). Duration: 10 mins. |
| Smith et al, 2020b Australia | RCT, between-subjects (Lab) | 52 (45) |  | N = 38 digital media - Web-based game in which the player is instructed to collect as many coins and brand logos as possible before time runs out, with a rectangular banner ad for the test brand directly beneath the game onscreen. Duration: 4 mins |  |  |
| Smith et al, 2020c Australia | RCT, between-subjects (Lab) | 52 (45) |  | N = 39 digital media - Participants collected coins before time ran out, with a 30-second rewarded video ad pausing the game at a lock and unlocking a new level with higher-value coins. Duration: 0.5 min |  |  |
| Theben et al, 2022 Netherlands | RCT, between-subjects (School) | 83 (60) | Age range: 7-13 years  Age mean (SD): 10.4 (1.79)  Weight status: 2.4% underweight, 84.6% normal weight, 12.2% overweigh, 0.8% obese  SEP: NR  Ethnicity: NR | N = 43 digital media - Fruit memory adver­game (a popular fruit brand's pictures and logos, and 8 different fruit, fruit drinks, or cups with fruit from this popular brand).  Duration: 5 mins | N = 40 digital media - Non-food advergame (a popular Dutch toy brand's pictures and logos, and 8 individual toys from this popular toy brand). | Post-intervention ad libitum intake of 2 pre-weighted bowls containing bananas and apple pieces. (Grams converted to kcal). Duration: 5 mins. |
| Verfay and Werle, 2025a France | RCT, between-subjects (Community centre) | 59 (52.5) | Age range: 6-11 years  Age mean (SD): 7.3 (1.51)  Weight status: NR  SEP: NR  Ethnicity: NR | Packaging - Exposure to a water bottle with a well-known character and a juice bottle with no character. Duration: NA | Packaging - Exposure to a regular water bottle without a character and a regular juice bottle without a character. | Post-intervention consumption of chosen beverage. (No data). Duration: NR. |
| Verfay and Werle, 2025b France | RCT, between-subjects (Community centre) | 121 (52) | Age range: 5-7 years  Age mean (SD): 6.1 (0.9)  Weight status: NR  SEP: NR  Ethnicity: NR | Packaging - Exposure to a water bottle with either a well-known character or an unknown character and a juice bottle with no character. Duration: NA | Packaging - Exposure to a regular water bottle without a character and a regular juice bottle without a character. | Post-intervention consumption of chosen beverage. (No data). Duration: NR. |
| vonNordheim et al, 2022 Germany | RCT, between-subjects (Nursery room) | 172 (51.7) | Age range: 3-7 years  Age mean (SD): 4.72 (0.99)  Weight status: NR  SEP: NR  Ethnicity: NR | N = 103 TV - 7-min TV episode (Shaun the Sheep) containing the same 1-min healthy food ad shown three times.  Duration: 3 mins | N = 69 TV - 7-min TV episode (Shaun the Sheep) containing the same 1-min toy ad shown three times. | Post-intervention ad libitum intake of seven healthy foods in bite-size portions (Apples, orange segments, carrot sticks, cucumber wheels, pepper slices, cherry tomatoes, and half slices of whole-meal bread). Duration: 30 mins. |
| Yeum et al, 2024 USA | RCT, within-subjects (Lab) | 177 (42.9) | Age range: 9-12 years  Age mean: 10.9 (1.18)  Weight status: 1.1% underweight, 70.1% normal weight, 14.1% overweight, 14.7% obese  SEP: 1.1% < $25 000, 11.9% $25 000 – $65 000, 48.6% $65 000 – $145 000, 34.3% $145 000 – $225 000, 9.6% >$225 000, 4.5% Prefer not to answer; Ethnicity: 94.4% non-Hispanic, 3.4% Hispanic, 2.3% missing | N = 177 TV - 35-min TV show (MythBusters) containing various branded food ads.  Duration: 7.3 mins | N = 177 TV - 35-min TV show (MythBusters) containing toy/electronic ads. | During intervention, ad libitum intake of one sweet snack (Gummy candy), one savory snack (Goldfish Crackers), and one low-energy-density snack (Red grapes). Duration: 35 mins. |

Footnotes:

- I1: Intervention 1; I2: Intervention 2

### **eTable 6a.** *Detailed Risk of Bias Assessment - Parallel Studies*

| **Author, Year** | Anschutz 2009 | Anschutz 2010 | Boyland 2013 | Coates 2019a | Coates 2019b | Edwards 2022 | Emond 2016 | Evans 2025 | Farrow 2019 | Folkvord 2013 | Folkvord 2014 | Folkvord 2015 | Folkvord 2016 | Folkvord 2017 | Fox 1980 | Gorn and Goldberg 1980 | Gilbert-Diamond 2017 |
| --- | --- | --- | --- | --- | --- | --- | --- | --- | --- | --- | --- | --- | --- | --- | --- | --- | --- |
| Domain 1: Randomization Process | | | | | | | | | | | | | | | | | |
| 1.1 | Y | Y | NI | Y | PY | Y | Y | Y | PN | Y | Y | Y | Y | PY | Y | PY | Y |
| 1.2 | Y | Y | Y | Y | Y | Y | Y | Y | Y | Y | Y | Y | Y | PY | Y | Y | Y |
| 1.3 | N | N | N | N | N | N | N | N | N | N | N | N | N | PN | N | PN | N |
| ROB | L | L | L | L | L | L | L | L | SC | L | L | L | L | L | L | L | L |
| Domain 2: Deviations from intended interventions | | | | | | | | | | | | | | | | | |
| 2.1 | N | NI | PY | Y | N | NI | NI | N | Y | N | N | N | N | N | N | N | N |
| 2.2 | Y | Y | PY | Y | Y | Y | PY | Y | Y | Y | Y | Y | Y | Y | Y | Y | Y |
| 2.3 | N | N | N | N | N | N | N | N | N | N | N | N | N | N | N | N | N |
| 2.4 | NA | NA | NA | NA | NA | NA | NA | NA | NA | NA | NA | NA | NA | NA | NA | NA | NA |
| 2.5 | NA | NA | NA | NA | NA | NA | NA | NA | NA | NA | NA | NA | NA | NA | NA | NA | NA |
| 2.6 | Y | Y | Y | Y | Y | Y | Y | Y | Y | Y | Y | Y | Y | Y | Y | Y | Y |
| 2.7 | NA | NA | NA | NA | NA | NA | NA | NA | NA | NA | NA | NA | NA | NA | NA | NA | NA |
| ROB | L | L | L | L | L | L | L | L | L | L | L | L | L | L | L | L | L |
| Domain 3: Missing outcome data | | | | | | | | | | | | | | | | | |
| 3.1 | Y | Y | Y | Y | Y | Y | Y | Y | Y | Y | Y | Y | Y | PY | Y | PY | Y |
| 3.2 | NA | NA | NA | NA | NA | NA | NA | NA | NA | NA | NA | NA | NA | NA | NA | NA | NA |
| 3.3 | NA | NA | NA | NA | NA | NA | NA | NA | NA | NA | NA | NA | NA | NA | NA | NA | NA |
| 3.4 | NA | NA | NA | NA | NA | NA | NA | NA | NA | NA | NA | NA | NA | NA | NA | NA | NA |
| ROB | L | L | L | L | L | L | L | L | L | L | L | L | L | L | L | L | L |
| Domain 4: Measuremeng of the outcome | | | | | | | | | | | | | | | | | |
| 4.1 | N | N | N | N | N | N | N | N | N | N | N | N | N | N | N | N | N |
| 4.2 | N | N | N | N | N | N | N | N | N | N | N | N | N | N | N | N | N |
| 4.3 | Y | Y | Y | Y | Y | Y | Y | Y | Y | Y | PY | Y | Y | PY | Y | PY | Y |
| 4.4 | N | N | N | N | N | PN | N | N | N | N | N | N | N | PN | N | PN | N |
| 4.5 | NA | NA | NA | NA | NA | NA | NA | NA | NA | NA | NA | NA | NA | NA | NA | NA | NA |
| ROB | L | L | L | L | L | L | L | L | L | L | L | L | L | L | L | L | L |
| Domain 5: Selection of the reported result | | | | | | | | | | | | | | | | | |
| 5.1 | NI | NI | N | NI | NI | NI | NI | Y | NI | Y | Y | NI | NI | NI | NI | NI | NI |
| 5.2 | N | N | N | N | N | N | N | N | N | N | N | N | N | N | N | N | N |
| 5.3 | N | N | N | N | N | N | N | N | N | N | N | N | N | N | N | N | N |
| ROB | SC | SC | SC | SC | SC | SC | SC | L | SC | L | L | SC | SC | SC | SC | SC | SC |
| Overall ROB | SC | SC | SC | SC | SC | SC | SC | L | SC | L | L | SC | SC | SC | SC | SC | SC |

| Gregori 2013 | Gregori 2014 | Harris 2009 | Harris 2012 | Harris 2018 | Hermans 2018 | Jeffrey 1982 | Keller 2012b | Kim 2024 | Lorenzoni 2017a | Lorenzoni 2017b | McGale 2020 | Sharps 2020a | Sharps 2020b | Smith 2020 | Theben 2022 | Verfay and Werle 2025a | Verfay and Werle 2025b |
| --- | --- | --- | --- | --- | --- | --- | --- | --- | --- | --- | --- | --- | --- | --- | --- | --- | --- |
| Domain 1: Randomization Process | | | | | | | | | | | | | | | | | |
| Y | Y | Y | Y | PY | Y | Y | Y | Y | Y | Y | Y | Y | Y | Y | Y | Y | Y |
| Y | Y | Y | Y | Y | Y | Y | Y | Y | Y | Y | Y | Y | Y | Y | Y | Y | Y |
| N | N | N | N | N | N | N | N | N | N | PN | N | N | N | N | N | N | N |
| L | L | L | L | L | L | L | L | L | L | L | L | L | L | L | L | L | L |
| Domain 2: Deviations from intended interventions | | | | | | | | | | | | | | | | | |
| NI | NI | N | NI | Y | N | NI | NI | NI | NI | NI | N | N | N | N | N | Y | Y |
| Y | PY | Y | Y | Y | Y | Y | Y | Y | Y | Y | Y | Y | Y | Y | Y | Y | Y |
| N | PN | N | N | N | N | N | N | N | N | N | N | N | N | N | N | N | N |
| NA | NA | NA | NA | NA | NA | NA | NA | NA | NA | NA | NA | NA | NA | NA | NA | NA | NA |
| NA | NA | NA | NA | NA | NA | NA | NA | NA | NA | NA | NA | NA | NA | NA | NA | NA | NA |
| Y | Y | Y | Y | Y | Y | Y | Y | Y | Y | Y | Y | Y | Y | Y | Y | Y | Y |
| NA | NA | NA | NA | NA | NA | NA | NA | NA | NA | NA | NA | NA | NA | NA | NA | NA | NA |
| L | L | L | L | L | L | L | L | L | L | L | L | L | L | L | L | L | L |
| Domain 3: Missing outcome data | | | | | | | | | | | | | | | | | |
| Y | Y | Y | Y | Y | Y | PY | N | Y | Y | Y | Y | Y | Y | Y | Y | Y | Y |
| NA | NA | NA | NA | NA | NA | NA | N | NA | NA | NA | NA | NA | NA | NA | NA | NA | NA |
| NA | NA | NA | NA | NA | NA | NA | PN | NA | NA | NA | NA | NA | NA | NA | NA | NA | NA |
| NA | NA | NA | NA | NA | NA | NA | NA | NA | NA | NA | NA | NA | NA | NA | NA | NA | NA |
| L | L | L | L | L | L | L | L | L | L | L | L | L | L | L | L | L | L |
| Domain 4: Measuremeng of the outcome | | | | | | | | | | | | | | | | | |
| N | N | N | N | N | N | N | N | N | N | N | N | N | N | N | N | N | N |
| N | N | N | N | N | N | N | N | N | N | N | N | N | N | N | N | N | N |
| PY | Y | Y | Y | Y | PY | Y | Y | Y | Y | Y | Y | Y | Y | Y | Y | Y | Y |
| N | N | N | N | N | N | N | N | N | N | N | N | N | N | N | N | N | N |
| NA | NA | NA | NA | NA | NA | NA | NA | NA | NA | NA | NA | NA | NA | NA | NA | NA | NA |
| L | L | L | L | L | L | L | L | L | L | L | L | L | L | L | L | L | L |
| Domain 5: Selection of the reported result | | | | | | | | | | | | | | | | | |
| NI | NI | NI | NI | NI | NI | NI | NI | NI | NI | NI | NI | NI | NI | Y | NI | NI | NI |
| N | N | N | N | N | N | N | N | N | N | N | N | N | N | N | N | N | N |
| Y | Y | N | N | N | N | N | N | N | Y | Y | N | N | N | N | N | N | N |
| H | H | SC | SC | SC | SC | SC | SC | SC | H | H | SC | SC | SC | L | SC | SC | SC |
| H | H | SC | SC | SC | SC | SC | SC | SC | H | H | SC | SC | SC | L | SC | SC | SC |

### **eTable 6b.** *Detailed Risk of Bias Assessment - Crossover Studies*

| **Author, Year** | Aerts 2017 | Aerts 2019a | Aerts 2019b | Anderson 2014 | Boyland 2026 | Dovey 2011 | Forman 2009 | Gregori 2017 | Halford 2004 | Halford 2007 | Halford 2008 | Kearney 2020 | Masterson 2019 | Neyens 2015 | Norman 2018 | Yeum 2024 |
| --- | --- | --- | --- | --- | --- | --- | --- | --- | --- | --- | --- | --- | --- | --- | --- | --- |
| Domain 1: Randomization Process | | | | | | | | | | | | | | | | |
| 1.1 | Y | NI | NI | Y | Y | NI | Y | Y | NI | NI | NI | Y | Y | Y | Y | Y |
| 1.2 | Y | Y | Y | Y | Y | Y | Y | Y | Y | Y | Y | Y | Y | Y | Y | Y |
| 1.3 | N | N | N | N | N | NI | N | N | NI | N | N | N | NI | N | N | N |
| ROB | L | L | L | L | L | L | L | L | L | L | L | L | L | L | L | L |
| Domain S. Risk of bias arising from period and carryover effects | | | | | | | | | | | | | | | | |
| S.1 | NI | NI | NI | NI | Y | NI | NI | Y | Y | NI | Y | Y | Y | NI | Y | Y |
| S.2 | N | N | N | N | NA | N | N | NA | NA | N | NA | NA | NA | N | NA | NA |
| S.3 | NI | NI | NI | NI | Y | NI | NI | NI | NI | NI | NI | Y | NI | NI | NI | NI |
| ROB | SC | SC | SC | SC | L | SC | SC | SC | SC | SC | SC | L | SC | SC | SC | SC |
| Domain 2: Deviations from intended interventions | | | | | | | | | | | | | | | | |
| 2.1 | N | N | N | NI | N | NI | N | NI | N | NI | N | NI | Y | N | N | NI |
| 2.2 | Y | Y | Y | Y | Y | Y | Y | Y | Y | Y | Y | Y | Y | Y | Y | Y |
| 2.3 | N | N | N | N | N | N | N | N | N | N | N | N | N | N | N | N |
| 2.4 | NA | NA | NA | NA | NA | NA | NA | NA | NA | NA | NA | NA | NA | NA | NA | NA |
| 2.5 | NA | NA | NA | NA | NA | NA | NA | NA | NA | NA | NA | NA | NA | NA | NA | NA |
| 2.6 | Y | Y | Y | Y | Y | Y | Y | Y | Y | Y | Y | Y | Y | Y | Y | Y |
| 2.7 | NA | NA | NA | NA | NA | NA | NA | NA | NA | NA | NA | NA | NA | NA | NA | NA |
| ROB | L | L | L | L | L | L | L | L | L | L | L | L | L | L | L | L |
| Domain 3: Missing outcome data | | | | | | | | | | | | | | | | |
| 3.1 | Y | Y | Y | Y | Y | Y | Y | Y | Y | Y | Y | Y | Y | Y | Y | N |
| 3.2 | NA | NA | NA | NA | NA | NA | NA | NA | NA | NA | NA | NA | NA | NA | NA | N |
| 3.3 | NA | NA | NA | NA | NA | NA | NA | NA | NA | NA | NA | NA | NA | NA | NA | N |
| 3.4 | NA | NA | NA | NA | NA | NA | NA | NA | NA | NA | NA | NA | NA | NA | NA | NA |
| ROB | L | L | L | L | L | L | L | L | L | L | L | L | L | L | L | L |
| Domain 4: Measuremeng of the outcome | | | | | | | | | | | | | | | | |
| 4.1 | N | N | N | N | N | N | N | N | N | N | N | N | N | N | N | N |
| 4.2 | N | N | N | N | N | N | N | N | N | N | N | N | N | N | N | N |
| 4.3 | Y | Y | Y | Y | Y | Y | Y | Y | Y | Y | Y | Y | Y | Y | Y | Y |
| 4.4 | N | N | N | N | N | N | N | N | N | N | N | N | N | N | N | N |
| 4.5 | NA | NA | NA | NA | NA | NA | NA | NA | NA | NA | NA | NA | NA | NA | NA | NA |
| ROB | L | L | L | L | L | L | L | L | L | L | L | L | L | L | L | L |
| Domain 5: Selection of the reported result | | | | | | | | | | | | | | | | |
| 5.1 | NI | NI | NI | NI | Y | NI | NI | NI | NI | NI | NI | PY | NI | NI | Y | PY |
| 5.2 | N | N | N | N | N | N | N | N | N | N | N | N | N | N | N | N |
| 5.3 | N | N | N | N | N | N | N | N | N | N | N | N | N | N | N | N |
| 5.4 | N | N | N | N | N | N | N | N | N | N | N | N | N | N | N | N |
| ROB | SC | SC | SC | SC | L | SC | SC | SC | SC | SC | SC | L | SC | SC | L | L |
| Overall ROB | SC | SC | SC | SC | L | SC | SC | SC | SC | SC | SC | L | SC | SC | SC | SC |

### **eTable 6c.** *Detailed Risk of Bias Assessment - Cluster Studies*

| **Author, Year** | Aerts 2017b | vonNordheim 2022 |
| --- | --- | --- |
| Domain 1a: Randomization Process | | |
| 1a.1 | Y | Y |
| 1a.2 | Y | Y |
| 1a.3 | N | N |
| ROB | L | L |
| Domain 1b: Risk of bias arising from the timing of identification or recruitment of participants | | |
| 1b.1 | Y | N |
| 1b.2 | NA | N |
| 1b.3 | N | N |
| ROB | L | L |
| Domain 2: Deviations from intended interventions | | |
| 2.1a | Y | PY |
| 2.1b | N | N |
| 2.2 | Y | Y |
| 2.3 | N | N |
| 2.4 | NA | NA |
| 2.5 | NA | NA |
| 2.6 | Y | Y |
| 2.7 | NA | NA |
| ROB | L | L |
| Domain 3: Missing outcome data | | |
| 3.1a | Y | Y |
| 3.1b | Y | Y |
| 3.2 | NA | NA |
| 3.3 | NA | NA |
| 3.4 | NA | NA |
| ROB | L | L |
| Domain 4: Measurement of the outcome | | |
| 4.1 | N | N |
| 4.2 | N | N |
| 4.3a | Y | Y |
| 4.3b | Y | Y |
| 4.4 | N | N |
| 4.5 | NA | NA |
| ROB | L | L |
| Domain 5: Selection of the reported result | | |
| 5.1 | NI | NI |
| 5.2 | N | N |
| 5.3 | N | N |
| ROB | SC | SC |
| Overall ROB | SC | SC |

### **eTable 6d**. *Detailed Risk of Bias Assessment – ROBINS-I*

| Author, Year | Brown 2017 | Leonard 2019 | Keller 2012a | Kotler 2012 |
| --- | --- | --- | --- | --- |
| Domain 1. Bias due to confounding (Variant A) | | | | |
| 1.1 | SN | SN | SN | PY |
| 1.2 | NA | NA | NA | PY |
| 1.3 | NA | NA | NA | NA |
| 1.4 | N | N | N | N |
| ROB | Serious Risk of Bias | Serious Risk of Bias | Serious Risk of Bias | Low risk of bias except for concerns about uncontrolled confounding |
| Domain 2. Bias in classification of interventions | | | | |
| 2.1 | N | N | N | N |
| 2.2 | NA | NA | NA | NA |
| 2.3 | Y | Y | Y | Y |
| 2.4 | N | N | N | N |
| 2.5 | Y | Y | Y | Y |
| ROB | Low | Low | Low | Low |
| Domain 3. Bias in selection of participants into the study (or into the analysis) | | | | |
| 3.1 | N | N | N | N |
| 3.2 | NA | NA | NA | NA |
| 3.3 | Y | Y | Y | Y |
| 3.4 | NA | NA | NA | NA |
| 3.5 | N | N | N | N |
| 3.6 | NA | NA | NA | NA |
| 3.7 | NA | NA | NA | NA |
| 3.8 | NA | NA | NA | NA |
| 3.9 | NA | NA | NA | NA |
| 3.10. | NA | NA | NA | NA |
| ROB | Low | Low | Low | Low |
| Domain 4. Bias due to deviations from intended interventions (Variant A) | | | | |
| 4.1 | Y | Y | Y | Y |
| 4.2 | N | N | N | N |
| 4.3 | N | N | N | N |
| 4.4 | NA | NA | NA | NA |
| 4.5 | Y | Y | Y | Y |
| ROB | Low | Low | Low | Low |
| Domain 5: Selection of the reported result | | | | |
| 5.1 | Y | Y | Y | Y |
| 5.2 | Y | Y | Y | Y |
| 5.3 | Y | Y | Y | Y |
| 5.4 | NA | NA | NA | NA |
| 5.5 | NA | NA | NA | NA |
| 5.6 | NA | NA | NA | NA |
| 5.7 | NA | NA | NA | NA |
| 5.8 | NA | NA | NA | NA |
| 5.9 | NA | NA | NA | NA |
| 5.10. | NA | NA | NA | NA |
| 5.11. | NA | NA | NA | NA |
| ROB | Low | Low | Low | Low |
| Domain 6. Bias in measurement of the outcome | | | | |
| 6.1 | N | N | N | N |
| 6.2 | Y | Y | Y | Y |
| 6.3 | N | N | N | N |
| ROB | Low | Low | Low | Low |
| Domain 7. Bias in measurement of the outcome | | | | |
| 7.1 | NI | NI | NI | NI |
| 7.2 | N | N | N | N |
| 7.3 | N | N | N | N |
| 7.4 | N | N | N | N |
| ROB | Low | Low | Low | Low |
| Overall ROB | Low risk of bias except for concerns about uncontrolled confounding | Low risk of bias except for concerns about uncontrolled confounding | Low risk of bias except for concerns about uncontrolled confounding | Low risk of bias except for concerns about uncontrolled confounding |

### **eTable 7a.** *Instrument to assess the Credibility of Effect Modification Analyses (ICEMAN) in a Meta-analysis of Randomized Controlled Trials – Marketing Medium*

| **CREDIBILITY ASSESSMENT** | | | |
| --- | --- | --- | --- |
| **Essential preliminary considerations to define the possible effect modification of interest** | | | |
| State a single candidate effect modifier (e.g., age or comorbidity): Medium used for marketing of unhealthy food | | | |
| Was the effect modifier measured before or at randomization? [**X**] yes, continue [ ] no, stop here and refer to manual for further instructions | | | |
| State a single outcome and time-point (e.g., mortality at 1 year follow-up): Post-/during intervention dietary intakes | | | |
| State a single effect measure (e.g., relative risk or risk difference): Mean difference | | | |
| **1: Is the analysis of effect modification based on comparison within rather than between trials?** | | | |
| [ **x** ] Completely between | [ ] Mostly between or unclear | [ ] Mostly within | [ ] Completely within |
| *Subgroup analysis or meta-regression comparing overall effects of each individual trial. This is typical for aggregate data meta-analysis.* | *Subgroup analysis or meta-regression with most information coming from overall effects, but some trials providing within-trial subgroup information* | *Most trials providing within-trial subgroup information; or individual participant data analysis that combines within and between trial information* | *All trials providing within-trial subgroup information or individual participant data; and the analysis separates within from between trial information, e.g., meta-analysis of interactions* |
| Comment: None of the trials in the subgroup analysis provided within-trial subgroup information. | | | |
| **2: For within-trial comparisons, is the effect modification similar from trial to trial?** [ **x** ] Not applicable: no or one within-RCT comparison | | | |
| [ ] Definitely not similar | [ ] Probably not similar or unclear | [ ] Mostly similar | [ ] Definitely similar |
| *Effect modification reported for two or more trials and clearly different directions* | *Effect modification not reported for individual trials or too imprecise to tell* | *Effect modification reported for two or more trials, mostly similar in direction, but considerable differences in magnitude* | *Effect modification reported for two or more trials, similar in direction, only some differences in magnitude* |
| Comment: | | | |
| **3: For between-trial comparisons, is the number of trials large?** [ ] Not applicable: no between RCT comparison | | | |
| [ ] Very small | [ ] Rather small or unclear | [ ] Rather large | [ **x** ] Large |
| *1 or 2 or in smallest subgroup; 5 or less in continuous meta-regression* | *3-4 in smallest subgroup; 6-10 in continuous meta-regression* | *5-9 in smallest subgroup; 11 to 15 in continuous meta-regression* | *10 or more in smallest subgroup; more than 15 in continuous meta-regression* |
| Comment: The packaging subgroup has 11 studies. | | | |
| **4: Was the direction of effect modification correctly hypothesized a priori?** | | | |
| [ ] Definitely no | [ **x** ] Probably no or unclear | [ ] Probably yes | [ ] Definitely yes |
| *Clearly post-hoc or results inconsistent with hypothesized direction or biologically very implausible* | *Vague hypothesis or hypothesized direction unclear* | *No prior protocol available but unequivocal statement of a priori hypothesis with correct direction of effect modification* | *Prior protocol available and includes correct specification of direction of effect modification, e.g., based on a biologic rationale* |
| Comment: Prior protocol is available, but only hypothesized that there will be a difference, with no specification of the direction of the difference. | | | |
| **5: Does a test for interaction suggest that chance is an unlikely explanation of the apparent effect modification?** (consider irrespective of number of effect modifiers) | | | |
| [ ] Chance a very likely explanation | [ ] Chance a likely explanation or unclear | [ ] Chance may not explain | [ **x** ] Chance an unlikely explanation |
| *Interaction or meta-regression p-value >0.05* | *Interaction or meta-regression p-value ≤0.05 and >0.01, or no test of interaction reported and not computable* | *Interaction or meta-regression p-value ≤0.01 and >0.005* | *Interaction or meta-regression p-value ≤0.005* |
| Comment: The p-value is small. P < 0.001 | | | |
| **6: Did the authors test only a small number of effect modifiers or consider the number in their statistical analysis?** | | | |
| [ ] Definitely no | [ ] Probably no or unclear | [ **x** ] Probably yes | [ ] Definitely yes |
| *Explicitly exploratory analysis or large number of effect modifiers tested (e.g., greater than 10) and multiplicity not considered in analysis* | *No mention of number or 4-10 effect modifiers tested and number not considered in analysis* | *No protocol available but unequivocal statement of 3 or fewer effect modifiers tested* | *Protocol available and 3 or fewer effect modifiers tested or number considered in analysis* |
| Comment: The protocol lists marketing medium as one of the effect modifiers, and 5 effect modifiers are tested. | | | |
| **7: Did the authors use a random effects model?** | | | |
| [ ] Definitely no | [ ] Probably no or unclear | [ ] Probably yes | [ **x** ] Definitely yes |
| *Fixed (or common) effect or fixed effects model explicitly stated* | *Probably fixed effect(s) model* | *Probably random (or mixed) effects* | *Random (or mixed) effects explicitly stated* |
| Comment: The use of the random effects model is explicitly stated | | | |
| **8: If the effect modifier is a continuous variable, were arbitrary cut points avoided?** [ **x** ] not applicable: not continuous | | | |
| [ ] Definitely no | [ ] Probably no or unclear | [ ] Probably yes | [ ] Definitely yes |
| *Analysis based on exploratory cut point(s), e.g., picking cut point associated with highest interaction p-value* | *Analysis based on cut point(s) of unclear origin* | *Analysis based on pre-specified cut point(s), e.g., suggested by prior RCT* | *Analysis based on the full continuum, e.g., assuming a linear or logarithmic relationship* |
| Comment: | | | |
| **9 Optional: Are there any additional considerations that may increase or decrease credibility?** (manual section 3.9) [ **x** ] not applicable | | | |
|  | [ ] Yes, probably decrease | [ ] Yes, probably increase | |
| Comment: **10: How would you rate the overall credibility of the proposed effect modification?** The overall rating should be driven by the items that decrease credibility. The following provides a sensible strategy: - All responses definitely or probably decrease credibility or unclear 🡪 very low - Two or more responses definitely decrease credibility 🡪 maximum usually low even if all other responses satisfy credibility criteria - One response definitely decreases credibility 🡪 maximum usually moderate even if all other responses satisfy credibility criteria - Two responses probably decrease credibility 🡪 maximum usually moderate even if all other responses satisfy credibility criteria - No response options definitely or probably decrease credibility 🡪 high very likely Place a mark on the continuous line (or type “x” in editable version) \| \| \| \| \|; \| **x** \| \| \| \|; \| **Very low credibility** \| **Low credibility** \| **Moderate credibility** \| **High credibility** \|; \| Minimal to no support for effect modification; Use overall effect for each subgroup \| Some but insufficient support for effect modification; Use overall effect for each subgroup but note remaining uncertainty \| Likely effect modification; Use separate effects for each subgroup but note remaining uncertainty \| Very likely effect modification; Use separate effects for each subgroup \|; Comment: The analysis is completely based on between-trial subgroup information, but a random effect model was used, and the interaction p-value is small. \| \| \| \| \| | | | |

### **eTable 7b.** *ICEMAN in a Meta-analysis of Randomized Controlled Trials – Sex*

| **CREDIBILITY ASSESSMENT** | | | |
| --- | --- | --- | --- |
| **Essential preliminary considerations to define the possible effect modification of interest** | | | |
| State a single candidate effect modifier (e.g., age or comorbidity): Sex | | | |
| Was the effect modifier measured before or at randomization? [**X**] yes, continue [ ] no, stop here and refer to manual for further instructions | | | |
| State a single outcome and time-point (e.g., mortality at 1 year follow-up): Post-/during intervention dietary intakes | | | |
| State a single effect measure (e.g., relative risk or risk difference): Mean difference | | | |
| **1: Is the analysis of effect modification based on comparison within rather than between trials?** | | | |
| [ ] Completely between | [ ] Mostly between or unclear | [ ] Mostly within | [**x**] Completely within |
| *Subgroup analysis or meta-regression comparing overall effects of each individual trial. This is typical for aggregate data meta-analysis.* | *Subgroup analysis or meta-regression with most information coming from overall effects, but some trials providing within-trial subgroup information* | *Most trials providing within-trial subgroup information; or individual participant data analysis that combines within and between trial information* | *All trials providing within-trial subgroup information or individual participant data; and the analysis separates within from between trial information, e.g., meta-analysis of interactions* |
| Comment: The effect modification by sex is suggested by comparison within three studies. | | | |
| **2: For within-trial comparisons, is the effect modification similar from trial to trial?** [ ] Not applicable: no or one within-RCT comparison | | | |
| [ ] Definitely not similar | [ ] Probably not similar or unclear | [ **x** ] Mostly similar | [ ] Definitely similar |
| *Effect modification reported for two or more trials and clearly different directions* | *Effect modification not reported for individual trials or too imprecise to tell* | *Effect modification reported for two or more trials, mostly similar in direction, but considerable differences in magnitude* | *Effect modification reported for two or more trials, similar in direction, only some differences in magnitude* |
| Comment: For females, the 2 reported trials have a positive effect, and the other two have a negative effect, but only one trial has confidence intervals above 0. For males, all the reported trials have a positive effect, while only one study has a confidence interval above 0. | | | |
| **3: For between-trial comparisons, is the number of trials large?** [ **x** ] Not applicable: no between RCT comparison | | | |
| [ ] Very small | [ ] Rather small or unclear | [ ] Rather large | [ ] Large |
| *1 or 2 or in smallest subgroup; 5 or less in continuous meta-regression* | *3-4 in smallest subgroup; 6-10 in continuous meta-regression* | *5-9 in smallest subgroup; 11 to 15 in continuous meta-regression* | *10 or more in smallest subgroup; more than 15 in continuous meta-regression* |
| Comment: | | | |
| **4: Was the direction of effect modification correctly hypothesized a priori?** | | | |
| [ ] Definitely no | [ **x** ] Probably no or unclear | [ ] Probably yes | [ ] Definitely yes |
| *Clearly post-hoc or results inconsistent with hypothesized direction or biologically very implausible* | *Vague hypothesis or hypothesized direction unclear* | *No prior protocol available but unequivocal statement of a priori hypothesis with correct direction of effect modification* | *Prior protocol available and includes correct specification of direction of effect modification, e.g., based on a biologic rationale* |
| Comment: Prior protocol is available, but only hypothesized that there will be a difference, with no specification of the direction of the difference. | | | |
| **5: Does a test for interaction suggest that chance is an unlikely explanation of the apparent effect modification?** (consider irrespective of number of effect modifiers) | | | |
| [ **x** ] Chance a very likely explanation | [ ] Chance a likely explanation or unclear | [ ] Chance may not explain | [ ] Chance an unlikely explanation |
| *Interaction or meta-regression p-value >0.05* | *Interaction or meta-regression p-value ≤0.05 and >0.01, or no test of interaction reported and not computable* | *Interaction or meta-regression p-value ≤0.01 and >0.005* | *Interaction or meta-regression p-value ≤0.005* |
| Comment: The finding requires prospective confirmation as the p-value = 0.082 | | | |
| **6: Did the authors test only a small number of effect modifiers or consider the number in their statistical analysis?** | | | |
| [ ] Definitely no | [ ] Probably no or unclear | [ **x** ] Probably yes | [ ] Definitely yes |
| *Explicitly exploratory analysis or large number of effect modifiers tested (e.g., greater than 10) and multiplicity not considered in analysis* | *No mention of number or 4-10 effect modifiers tested and number not considered in analysis* | *No protocol available but unequivocal statement of 3 or fewer effect modifiers tested* | *Protocol available and 3 or fewer effect modifiers tested or number considered in analysis* |
| Comment: The protocol lists age as one of the effect modifiers, and 5 effect modifiers are tested. | | | |
| **7: Did the authors use a random effects model?** | | | |
| [ ] Definitely no | [ ] Probably no or unclear | [ ] Probably yes | [ **x** ] Definitely yes |
| *Fixed (or common) effect or fixed effects model explicitly stated* | *Probably fixed effect(s) model* | *Probably random (or mixed) effects* | *Random (or mixed) effects explicitly stated* |
| Comment: The use of the random effects model is explicitly stated | | | |
| **8: If the effect modifier is a continuous variable, were arbitrary cut points avoided?** [ **x** ] not applicable: not continuous | | | |
| [ ] Definitely no | [ ] Probably no or unclear | [ ] Probably yes | [ ] Definitely yes |
| *Analysis based on exploratory cut point(s), e.g., picking cut point associated with highest interaction p-value* | *Analysis based on cut point(s) of unclear origin* | *Analysis based on pre-specified cut point(s), e.g., suggested by prior RCT* | *Analysis based on the full continuum, e.g., assuming a linear or logarithmic relationship* |
| Comment: | | | |
| **9 Optional: Are there any additional considerations that may increase or decrease credibility?** (manual section 3.9) [ **x** ] not applicable | | | |
|  | [ ] Yes, probably decrease | [ ] Yes, probably increase | |
| Comment: **10: How would you rate the overall credibility of the proposed effect modification?** The overall rating should be driven by the items that decrease credibility. The following provides a sensible strategy: - All responses definitely or probably decrease credibility or unclear 🡪 very low - Two or more responses definitely decrease credibility 🡪 maximum usually low even if all other responses satisfy credibility criteria - One response definitely decreases credibility 🡪 maximum usually moderate even if all other responses satisfy credibility criteria - Two responses probably decrease credibility 🡪 maximum usually moderate even if all other responses satisfy credibility criteria - No response options definitely or probably decrease credibility 🡪 high very likely Place a mark on the continuous line (or type “x” in editable version) \| \| \| \| \|; \| **x** \| \| \| \|; \| **Very low credibility** \| **Low credibility** \| **Moderate credibility** \| **High credibility** \|; \| Minimal to no support for effect modification; Use overall effect for each subgroup \| Some but insufficient support for effect modification; Use overall effect for each subgroup but note remaining uncertainty \| Likely effect modification; Use separate effects for each subgroup but note remaining uncertainty \| Very likely effect modification; Use separate effects for each subgroup \|; Comment: There is clear consistency across studies and mention of prior knowledge of the subgroup differences. Random effect model was used, but the P-value is relatively large. \| \| \| \| \| | | | |

### **eTable 7c.** *ICEMAN in a Meta-analysis of Randomized Controlled Trials – Age Groups*

| **CREDIBILITY ASSESSMENT** | | | |
| --- | --- | --- | --- |
| **Essential preliminary considerations to define the possible effect modification of interest** | | | |
| State a single candidate effect modifier (e.g., age or comorbidity): Age groups | | | |
| Was the effect modifier measured before or at randomization? [**X**] yes, continue [ ] no, stop here and refer to manual for further instructions | | | |
| State a single outcome and time-point (e.g., mortality at 1 year follow-up): Post-/during intervention dietary intakes | | | |
| State a single effect measure (e.g., relative risk or risk difference): Mean difference | | | |
| **1: Is the analysis of effect modification based on comparison within rather than between trials?** | | | |
| [ **x** ] Completely between | [ ] Mostly between or unclear | [ ] Mostly within | [] Completely within |
| *Subgroup analysis or meta-regression comparing overall effects of each individual trial. This is typical for aggregate data meta-analysis.* | *Subgroup analysis or meta-regression with most information coming from overall effects, but some trials providing within-trial subgroup information* | *Most trials providing within-trial subgroup information; or individual participant data analysis that combines within and between trial information* | *All trials providing within-trial subgroup information or individual participant data; and the analysis separates within from between trial information, e.g., meta-analysis of interactions* |
| Comment: The subgroup analysis compared the overall effects across individual trials. No trial provided within-trial subgroup information. | | | |
| **2: For within-trial comparisons, is the effect modification similar from trial to trial?** [ **x** ] Not applicable: no or one within-RCT comparison | | | |
| [ ] Definitely not similar | [ ] Probably not similar or unclear | [ ] Mostly similar | [ ] Definitely similar |
| *Effect modification reported for two or more trials and clearly different directions* | *Effect modification not reported for individual trials or too imprecise to tell* | *Effect modification reported for two or more trials, mostly similar in direction, but considerable differences in magnitude* | *Effect modification reported for two or more trials, similar in direction, only some differences in magnitude* |
| Comment: | | | |
| **3: For between-trial comparisons, is the number of trials large?** [ ] Not applicable: no between RCT comparison | | | |
| [ ] Very small | [ ] Rather small or unclear | [ **x** ] Rather large | [ ] Large |
| *1 or 2 or in smallest subgroup; 5 or less in continuous meta-regression* | *3-4 in smallest subgroup; 6-10 in continuous meta-regression* | *5-9 in smallest subgroup; 11 to 15 in continuous meta-regression* | *10 or more in smallest subgroup; more than 15 in continuous meta-regression* |
| Comment: There are 6 trials in the smallest subgroup. | | | |
| **4: Was the direction of effect modification correctly hypothesized a priori?** | | | |
| [ ] Definitely no | [ **x** ] Probably no or unclear | [ ] Probably yes | [ ] Definitely yes |
| *Clearly post-hoc or results inconsistent with hypothesized direction or biologically very implausible* | *Vague hypothesis or hypothesized direction unclear* | *No prior protocol available but unequivocal statement of a priori hypothesis with correct direction of effect modification* | *Prior protocol available and includes correct specification of direction of effect modification, e.g., based on a biologic rationale* |
| Comment: Prior protocol is available, but only hypothesized that there will be a difference, with no specification of the direction of the difference. | | | |
| **5: Does a test for interaction suggest that chance is an unlikely explanation of the apparent effect modification?** (consider irrespective of number of effect modifiers) | | | |
| [ ] Chance a very likely explanation | [ ] Chance a likely explanation or unclear | [ ] Chance may not explain | [ **x** ] Chance an unlikely explanation |
| *Interaction or meta-regression p-value >0.05* | *Interaction or meta-regression p-value ≤0.05 and >0.01, or no test of interaction reported and not computable* | *Interaction or meta-regression p-value ≤0.01 and >0.005* | *Interaction or meta-regression p-value ≤0.005* |
| Comment: The p-value is small (= 0.003) | | | |
| **6: Did the authors test only a small number of effect modifiers or consider the number in their statistical analysis?** | | | |
| [ ] Definitely no | [ ] Probably no or unclear | [ **x** ] Probably yes | [ ] Definitely yes |
| *Explicitly exploratory analysis or large number of effect modifiers tested (e.g., greater than 10) and multiplicity not considered in analysis* | *No mention of number or 4-10 effect modifiers tested and number not considered in analysis* | *No protocol available but unequivocal statement of 3 or fewer effect modifiers tested* | *Protocol available and 3 or fewer effect modifiers tested or number considered in analysis* |
| Comment: The protocol lists age group as one of the effect modifiers, and 5 effect modifiers are tested. | | | |
| **7: Did the authors use a random effects model?** | | | |
| [ ] Definitely no | [ ] Probably no or unclear | [ ] Probably yes | [ **x** ] Definitely yes |
| *Fixed (or common) effect or fixed effects model explicitly stated* | *Probably fixed effect(s) model* | *Probably random (or mixed) effects* | *Random (or mixed) effects explicitly stated* |
| Comment: The use of the random effects model is explicitly stated | | | |
| **8: If the effect modifier is a continuous variable, were arbitrary cut points avoided?** [ **x** ] not applicable: not continuous | | | |
| [ ] Definitely no | [ ] Probably no or unclear | [ ] Probably yes | [ ] Definitely yes |
| *Analysis based on exploratory cut point(s), e.g., picking cut point associated with highest interaction p-value* | *Analysis based on cut point(s) of unclear origin* | *Analysis based on pre-specified cut point(s), e.g., suggested by prior RCT* | *Analysis based on the full continuum, e.g., assuming a linear or logarithmic relationship* |
| Comment: | | | |
| **9 Optional: Are there any additional considerations that may increase or decrease credibility?** (manual section 3.9) [ **x** ] not applicable | | | |
|  | [ ] Yes, probably decrease | [ ] Yes, probably increase | |
| Comment: **10: How would you rate the overall credibility of the proposed effect modification?** The overall rating should be driven by the items that decrease credibility. The following provides a sensible strategy: - All responses definitely or probably decrease credibility or unclear 🡪 very low - Two or more responses definitely decrease credibility 🡪 maximum usually low even if all other responses satisfy credibility criteria - One response definitely decreases credibility 🡪 maximum usually moderate even if all other responses satisfy credibility criteria - Two responses probably decrease credibility 🡪 maximum usually moderate even if all other responses satisfy credibility criteria - No response options definitely or probably decrease credibility 🡪 high very likely Place a mark on the continuous line (or type “x” in editable version) \| \| \| \| \|; \| **x** \| \| \| \|; \| **Very low credibility** \| **Low credibility** \| **Moderate credibility** \| **High credibility** \|; \| Minimal to no support for effect modification; Use overall effect for each subgroup \| Some but insufficient support for effect modification; Use overall effect for each subgroup but note remaining uncertainty \| Likely effect modification; Use separate effects for each subgroup but note remaining uncertainty \| Very likely effect modification; Use separate effects for each subgroup \|; Comment: A random effect model was used, with a relatively large number of trials and a small P-value. Prior knowledge of the subgroup differences is mentioned. However, the analysis is completely based on between-trial comparisons. \| \| \| \| \| | | | |

### **eTable 7d.** *ICEMAN in a Meta-analysis of Randomized Controlled Trials – Weight Status*

| **CREDIBILITY ASSESSMENT** | | | |
| --- | --- | --- | --- |
| **Essential preliminary considerations to define the possible effect modification of interest** | | | |
| State a single candidate effect modifier (e.g., age or comorbidity): Weight status | | | |
| Was the effect modifier measured before or at randomization? [**X**] yes, continue [ ] no, stop here and refer to manual for further instructions | | | |
| State a single outcome and time-point (e.g., mortality at 1 year follow-up): Post-/during intervention dietary intakes | | | |
| State a single effect measure (e.g., relative risk or risk difference): Mean difference | | | |
| **1: Is the analysis of effect modification based on comparison within rather than between trials?** | | | |
| [ ] Completely between | [ ] Mostly between or unclear | [ ] Mostly within | [**x**] Completely within |
| *Subgroup analysis or meta-regression comparing overall effects of each individual trial. This is typical for aggregate data meta-analysis.* | *Subgroup analysis or meta-regression with most information coming from overall effects, but some trials providing within-trial subgroup information* | *Most trials providing within-trial subgroup information; or individual participant data analysis that combines within and between trial information* | *All trials providing within-trial subgroup information or individual participant data; and the analysis separates within from between trial information, e.g., meta-analysis of interactions* |
| Comment: The effect modification by weight status is suggested by comparison within five studies. | | | |
| **2: For within-trial comparisons, is the effect modification similar from trial to trial?** [ ] Not applicable: no or one within-RCT comparison | | | |
| [ ] Definitely not similar | [ ] Probably not similar or unclear | [ **x** ] Mostly similar | [ ] Definitely similar |
| *Effect modification reported for two or more trials and clearly different directions* | *Effect modification not reported for individual trials or too imprecise to tell* | *Effect modification reported for two or more trials, mostly similar in direction, but considerable differences in magnitude* | *Effect modification reported for two or more trials, similar in direction, only some differences in magnitude* |
| Comment: For normal weight, the 4 reported trials have a positive effect, with one confidence interval including zero. The other two have a negative effect, but one of them has confidence intervals that include zero. For overweight or obese individuals, all reported trials show a positive effect, and half have confidence intervals that include 0. | | | |
| **3: For between-trial comparisons, is the number of trials large?** [ **x** ] Not applicable: no between RCT comparison | | | |
| [ ] Very small | [ ] Rather small or unclear | [ ] Rather large | [ ] Large |
| *1 or 2 or in smallest subgroup; 5 or less in continuous meta-regression* | *3-4 in smallest subgroup; 6-10 in continuous meta-regression* | *5-9 in smallest subgroup; 11 to 15 in continuous meta-regression* | *10 or more in smallest subgroup; more than 15 in continuous meta-regression* |
| Comment: | | | |
| **4: Was the direction of effect modification correctly hypothesized a priori?** | | | |
| [ ] Definitely no | [ **x** ] Probably no or unclear | [ ] Probably yes | [ ] Definitely yes |
| *Clearly post-hoc or results inconsistent with hypothesized direction or biologically very implausible* | *Vague hypothesis or hypothesized direction unclear* | *No prior protocol available but unequivocal statement of a priori hypothesis with correct direction of effect modification* | *Prior protocol available and includes correct specification of direction of effect modification, e.g., based on a biologic rationale* |
| Comment: The paper does not clarify whether the effect modification was hypothesized a priori. | | | |
| **5: Does a test for interaction suggest that chance is an unlikely explanation of the apparent effect modification?** (consider irrespective of number of effect modifiers) | | | |
| [ ] Chance a very likely explanation | [ **x** ] Chance a likely explanation or unclear | [ ] Chance may not explain | [ ] Chance an unlikely explanation |
| *Interaction or meta-regression p-value >0.05* | *Interaction or meta-regression p-value ≤0.05 and >0.01, or no test of interaction reported and not computable* | *Interaction or meta-regression p-value ≤0.01 and >0.005* | *Interaction or meta-regression p-value ≤0.005* |
| Comment: The p-value is not small. P = 0.012. | | | |
| **6: Did the authors test only a small number of effect modifiers or consider the number in their statistical analysis?** | | | |
| [ ] Definitely no | [ ] Probably no or unclear | [ **x** ] Probably yes | [ ] Definitely yes |
| *Explicitly exploratory analysis or large number of effect modifiers tested (e.g., greater than 10) and multiplicity not considered in analysis* | *No mention of number or 4-10 effect modifiers tested and number not considered in analysis* | *No protocol available but unequivocal statement of 3 or fewer effect modifiers tested* | *Protocol available and 3 or fewer effect modifiers tested or number considered in analysis* |
| Comment: The protocol did not include weight status, and 5 effect modifiers are tested. | | | |
| **7: Did the authors use a random effects model?** | | | |
| [ ] Definitely no | [ ] Probably no or unclear | [ ] Probably yes | [ **x** ] Definitely yes |
| *Fixed (or common) effect or fixed effects model explicitly stated* | *Probably fixed effect(s) model* | *Probably random (or mixed) effects* | *Random (or mixed) effects explicitly stated* |
| Comment: The use of the random effects model is explicitly stated | | | |
| **8: If the effect modifier is a continuous variable, were arbitrary cut points avoided?** [ **x** ] not applicable: not continuous | | | |
| [ ] Definitely no | [ ] Probably no or unclear | [ ] Probably yes | [ ] Definitely yes |
| *Analysis based on exploratory cut point(s), e.g., picking cut point associated with highest interaction p-value* | *Analysis based on cut point(s) of unclear origin* | *Analysis based on pre-specified cut point(s), e.g., suggested by prior RCT* | *Analysis based on the full continuum, e.g., assuming a linear or logarithmic relationship* |
| Comment: | | | |
| **9 Optional: Are there any additional considerations that may increase or decrease credibility?** (manual section 3.9) [ **x** ] not applicable | | | |
|  | [ ] Yes, probably decrease | [ ] Yes, probably increase | |
| Comment: **10: How would you rate the overall credibility of the proposed effect modification?** The overall rating should be driven by the items that decrease credibility. The following provides a sensible strategy: - All responses definitely or probably decrease credibility or unclear 🡪 very low - Two or more responses definitely decrease credibility 🡪 maximum usually low even if all other responses satisfy credibility criteria - One response definitely decreases credibility 🡪 maximum usually moderate even if all other responses satisfy credibility criteria - Two responses probably decrease credibility 🡪 maximum usually moderate even if all other responses satisfy credibility criteria - No response options definitely or probably decrease credibility 🡪 high very likely Place a mark on the continuous line (or type “x” in editable version) \| \| \| \| \|; \| **x** \| \| \| \|; \| **Very low credibility** \| **Low credibility** \| **Moderate credibility** \| **High credibility** \|; \| Minimal to no support for effect modification; Use overall effect for each subgroup \| Some but insufficient support for effect modification; Use overall effect for each subgroup but note remaining uncertainty \| Likely effect modification; Use separate effects for each subgroup but note remaining uncertainty \| Very likely effect modification; Use separate effects for each subgroup \|; Comment: There is some consistency across studies. A random effect model was used, but the P-value is not small, and the hypothesis is not included in the published protocol. \| \| \| \| \| | | | |

### **eTable 7e.** *ICEMAN in a Meta-analysis of Randomized Controlled Trials – Exposure Duration of Marketing*

| **CREDIBILITY ASSESSMENT** | | | |
| --- | --- | --- | --- |
| **Essential preliminary considerations to define the possible effect modification of interest** | | | |
| State a single candidate effect modifier (e.g., age or comorbidity): Exposure duration of marketing | | | |
| Was the effect modifier measured before or at randomization? [**X**] yes, continue [ ] no, stop here and refer to manual for further instructions | | | |
| State a single outcome and time-point (e.g., mortality at 1 year follow-up): Post-/during intervention dietary intakes | | | |
| State a single effect measure (e.g., relative risk or risk difference): Mean difference | | | |
| **1: Is the analysis of effect modification based on comparison within rather than between trials?** | | | |
| [ **x** ] Completely between | [ ] Mostly between or unclear | [ ] Mostly within | [ ] Completely within |
| *Subgroup analysis or meta-regression comparing overall effects of each individual trial. This is typical for aggregate data meta-analysis.* | *Subgroup analysis or meta-regression with most information coming from overall effects, but some trials providing within-trial subgroup information* | *Most trials providing within-trial subgroup information; or individual participant data analysis that combines within and between trial information* | *All trials providing within-trial subgroup information or individual participant data; and the analysis separates within from between trial information, e.g., meta-analysis of interactions* |
| Comment: None of the trials in the subgroup analysis provided within-trial subgroup information. | | | |
| **2: For within-trial comparisons, is the effect modification similar from trial to trial?** [ **x** ] Not applicable: no or one within-RCT comparison | | | |
| [ ] Definitely not similar | [ ] Probably not similar or unclear | [ ] Mostly similar | [ ] Definitely similar |
| *Effect modification reported for two or more trials and clearly different directions* | *Effect modification not reported for individual trials or too imprecise to tell* | *Effect modification reported for two or more trials, mostly similar in direction, but considerable differences in magnitude* | *Effect modification reported for two or more trials, similar in direction, only some differences in magnitude* |
| Comment: | | | |
| **3: For between-trial comparisons, is the number of trials large?** [ ] Not applicable: no between RCT comparison | | | |
| [ **x** ] Very small | [ ] Rather small or unclear | [ ] Rather large | [ ] Large |
| *1 or 2 or in smallest subgroup; 5 or less in continuous meta-regression* | *3-4 in smallest subgroup; 6-10 in continuous meta-regression* | *5-9 in smallest subgroup; 11 to 15 in continuous meta-regression* | *10 or more in smallest subgroup; more than 15 in continuous meta-regression* |
| Comment: The digital media subgroup, for more than 5 mins, has only 2 studies. | | | |
| **4: Was the direction of effect modification correctly hypothesized a priori?** | | | |
| [ ] Definitely no | [ ] Probably no or unclear | [ ] Probably yes | [ ] Definitely yes |
| *Clearly post-hoc or results inconsistent with hypothesized direction or biologically very implausible* | *Vague hypothesis or hypothesized direction unclear* | *No prior protocol available but unequivocal statement of a priori hypothesis with correct direction of effect modification* | *Prior protocol available and includes correct specification of direction of effect modification, e.g., based on a biologic rationale* |
| Comment: This effect modification was not hypothesized a priori in the published protocol. | | | |
| **5: Does a test for interaction suggest that chance is an unlikely explanation of the apparent effect modification?** (consider irrespective of number of effect modifiers) | | | |
| [ **x** ] Chance a very likely explanation | [ ] Chance a likely explanation or unclear | [ ] Chance may not explain | [ ] Chance an unlikely explanation |
| *Interaction or meta-regression p-value >0.05* | *Interaction or meta-regression p-value ≤0.05 and >0.01, or no test of interaction reported and not computable* | *Interaction or meta-regression p-value ≤0.01 and >0.005* | *Interaction or meta-regression p-value ≤0.005* |
| Comment: The p-value is large, especially for TV. P = 0.044 for digital media. P = 0.703 for TV. | | | |
| **6: Did the authors test only a small number of effect modifiers or consider the number in their statistical analysis?** | | | |
| [ ] Definitely no | [ ] Probably no or unclear | [ **x** ] Probably yes | [ ] Definitely yes |
| *Explicitly exploratory analysis or large number of effect modifiers tested (e.g., greater than 10) and multiplicity not considered in analysis* | *No mention of number or 4-10 effect modifiers tested and number not considered in analysis* | *No protocol available but unequivocal statement of 3 or fewer effect modifiers tested* | *Protocol available and 3 or fewer effect modifiers tested or number considered in analysis* |
| Comment: The protocol did not include the duration of marketing, and 5 effect modifiers are tested. | | | |
| **7: Did the authors use a random effects model?** | | | |
| [ ] Definitely no | [ ] Probably no or unclear | [ ] Probably yes | [ **x** ] Definitely yes |
| *Fixed (or common) effect or fixed effects model explicitly stated* | *Probably fixed effect(s) model* | *Probably random (or mixed) effects* | *Random (or mixed) effects explicitly stated* |
| Comment: The use of the random effects model is explicitly stated | | | |
| **8: If the effect modifier is a continuous variable, were arbitrary cut points avoided?** [ ] not applicable: not continuous | | | |
| [ ] Definitely no | [ ] Probably no or unclear | [ **x** ] Probably yes | [ ] Definitely yes |
| *Analysis based on exploratory cut point(s), e.g., picking cut point associated with highest interaction p-value* | *Analysis based on cut point(s) of unclear origin* | *Analysis based on pre-specified cut point(s), e.g., suggested by prior RCT* | *Analysis based on the full continuum, e.g., assuming a linear or logarithmic relationship* |
| Comment: Analysis based on pre-specified cut point suggested by previous review. | | | |
| **9 Optional: Are there any additional considerations that may increase or decrease credibility?** (manual section 3.9) [ **x** ] not applicable | | | |
|  | [ ] Yes, probably decrease | [ ] Yes, probably increase | |
| Comment: **10: How would you rate the overall credibility of the proposed effect modification?** The overall rating should be driven by the items that decrease credibility. The following provides a sensible strategy: - All responses definitely or probably decrease credibility or unclear 🡪 very low - Two or more responses definitely decrease credibility 🡪 maximum usually low even if all other responses satisfy credibility criteria - One response definitely decreases credibility 🡪 maximum usually moderate even if all other responses satisfy credibility criteria - Two responses probably decrease credibility 🡪 maximum usually moderate even if all other responses satisfy credibility criteria - No response options definitely or probably decrease credibility 🡪 high very likely Place a mark on the continuous line (or type “x” in editable version) \| \| \| \| \|; \| **x** \| \| \| \|; \| **Very low credibility** \| **Low credibility** \| **Moderate credibility** \| **High credibility** \|; \| Minimal to no support for effect modification; Use overall effect for each subgroup \| Some but insufficient support for effect modification; Use overall effect for each subgroup but note remaining uncertainty \| Likely effect modification; Use separate effects for each subgroup but note remaining uncertainty \| Very likely effect modification; Use separate effects for each subgroup \|; Comment: The analysis is completely based on between-trial subgroup information, and one of the subgroups only contains 2 studies. Also, the interaction p-value is large. \| \| \| \| \| | | | |

### **eFigure 1a.** *Risk of Bias Assessment – Parallel Studies*

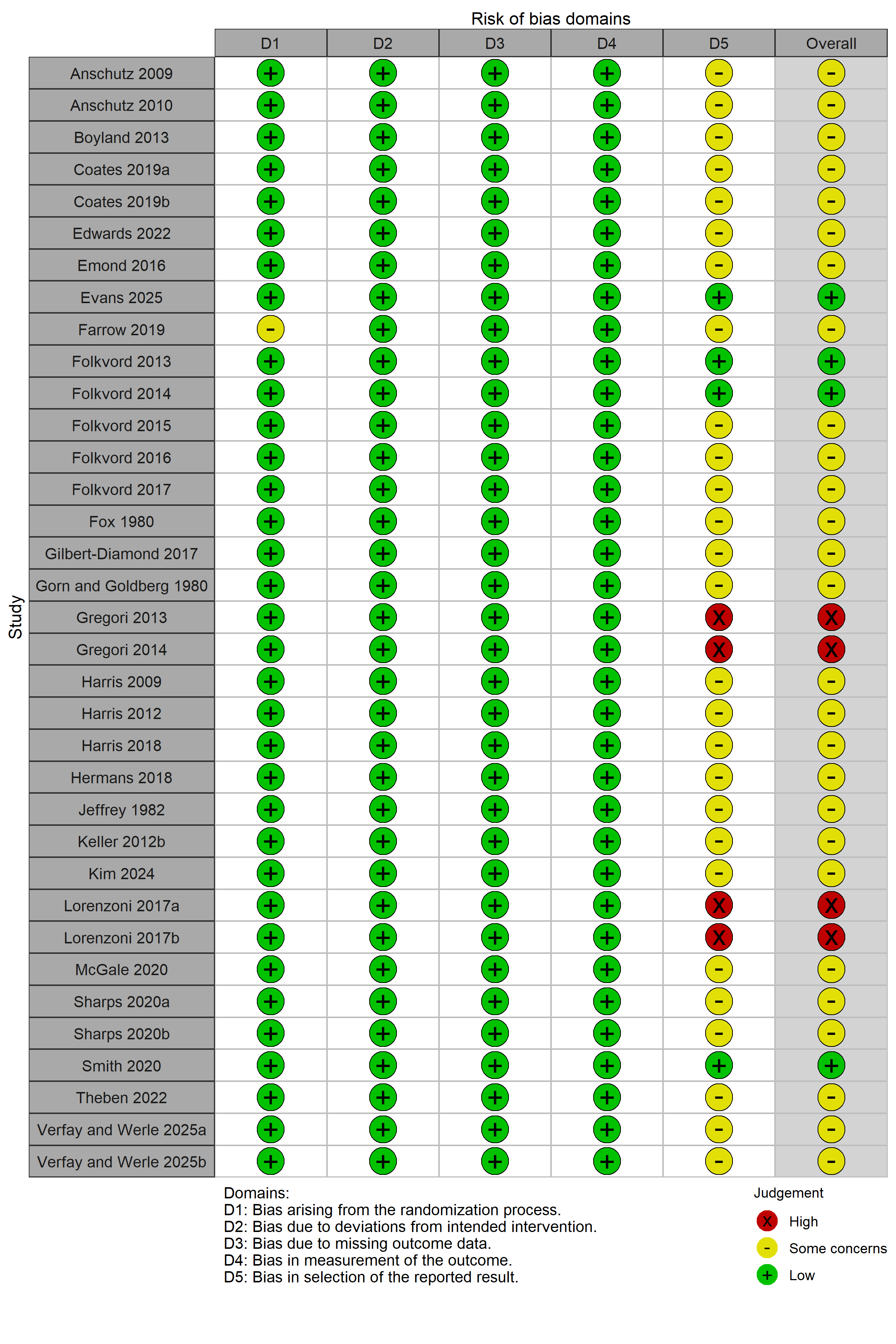

### **eFigure 1b.** *Risk of Bias Assessment – Crossover Studies*

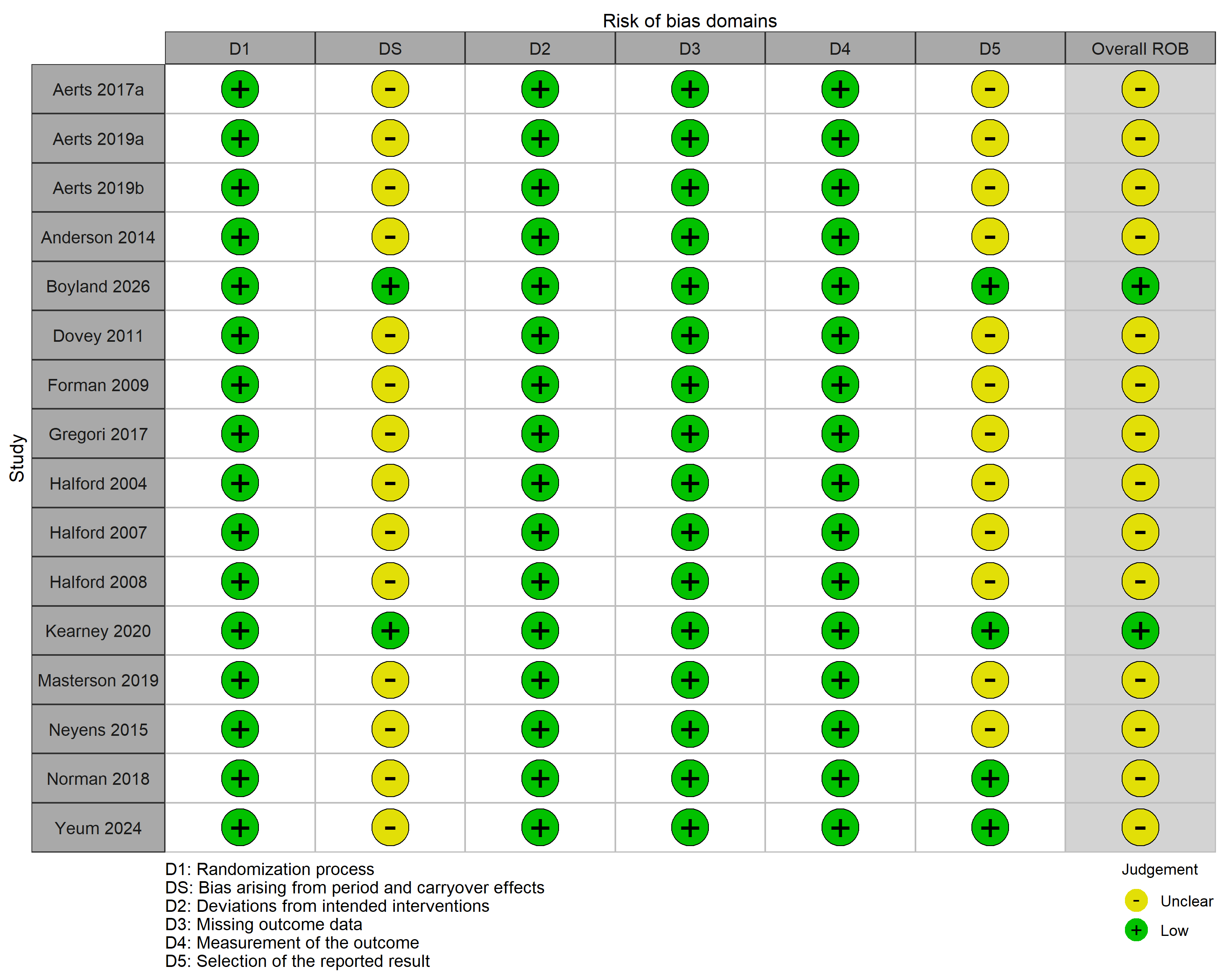

### **eFigure 1c.** *Risk of Bias Assessment – Cluster Studies*

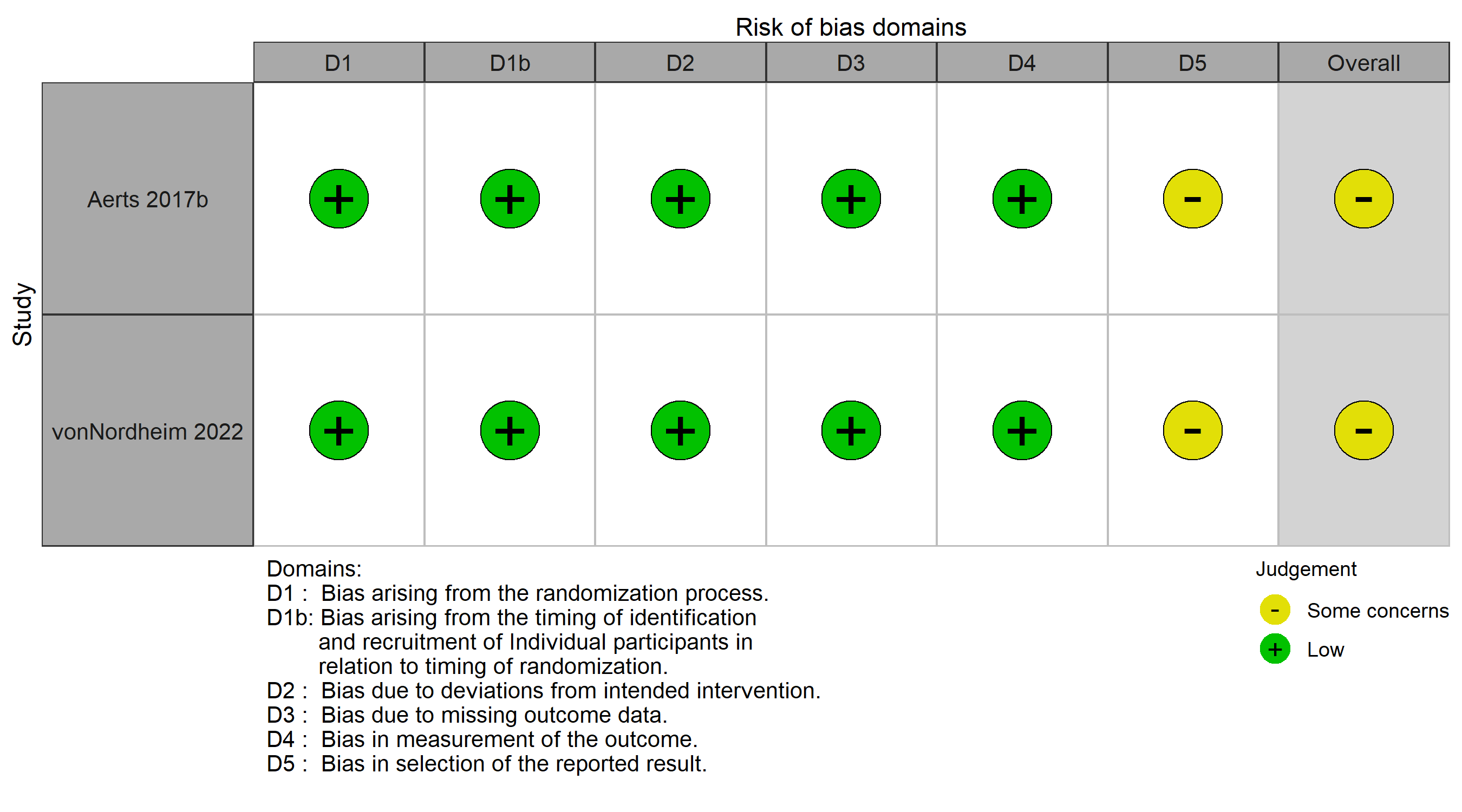

### **eFigure 1d**. *Risk of Bias Assessment – ROBINS-I*

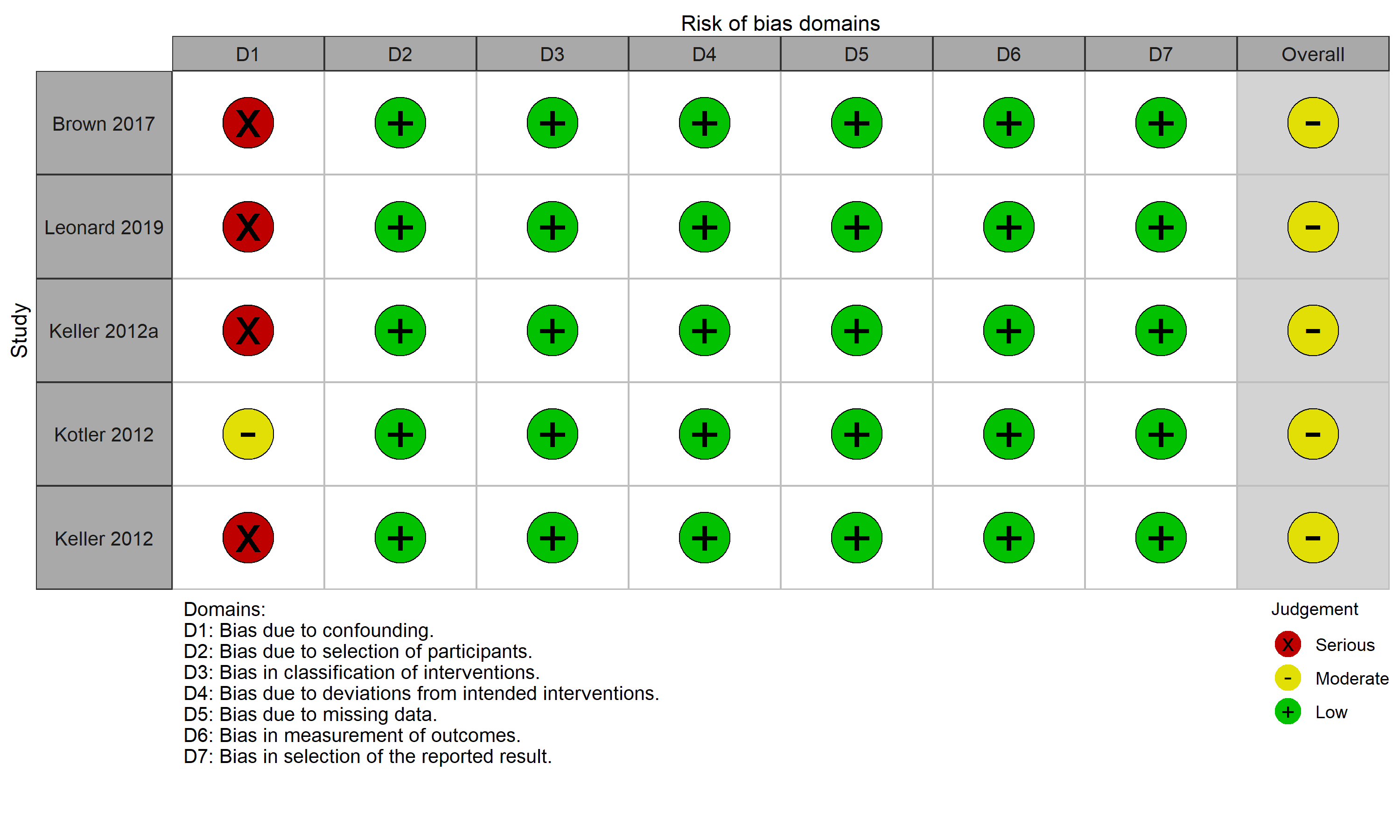

### **eFigure 2.** *Effect of M2K on Dietary Intakes by Marketing Medium in Randomized Clinical Trials (RCTs)*

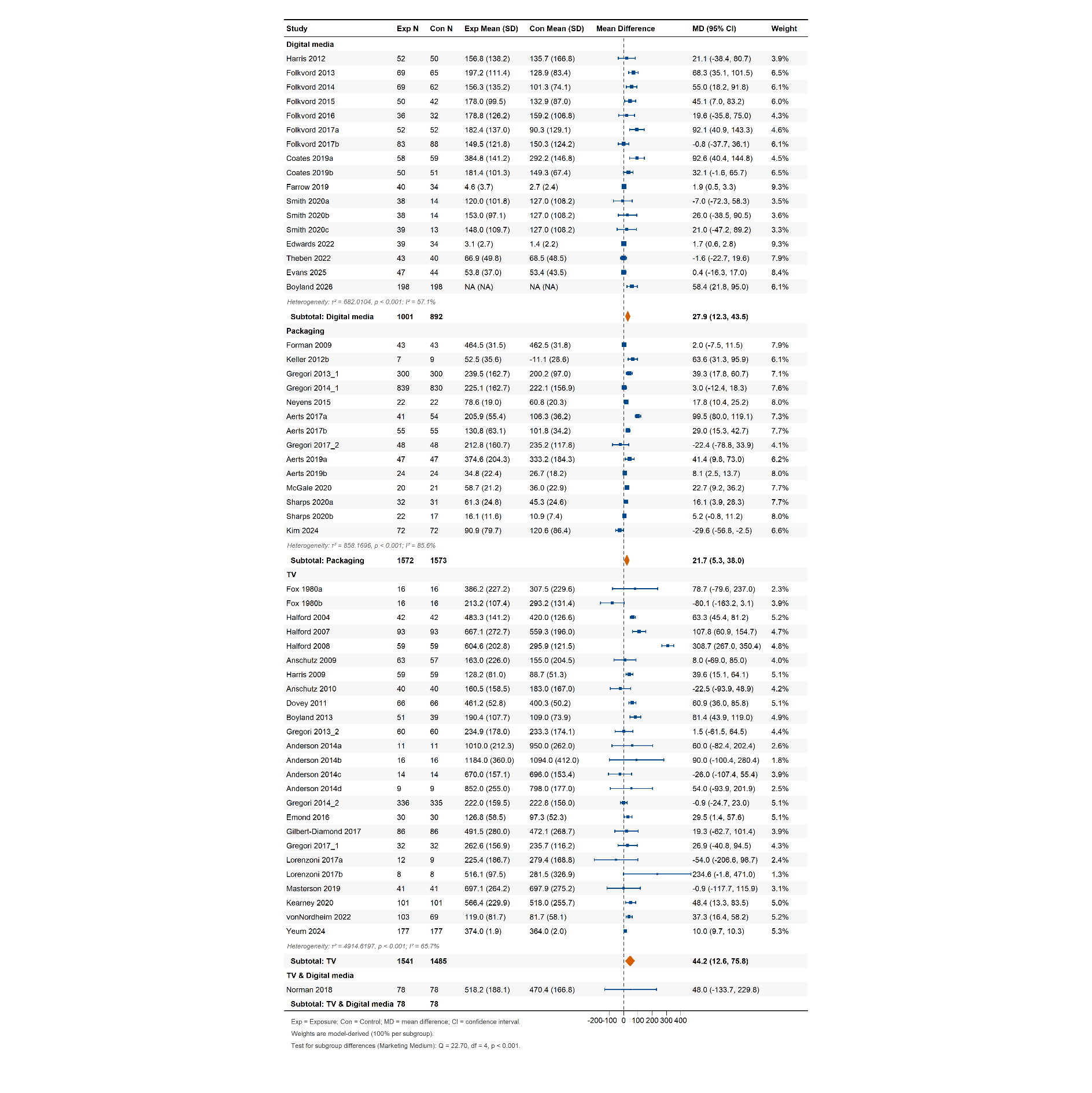

### **eFigure 3.** *Effect of Healthy M2K by Marketing Medium on Dietary Intakes in RCTs*

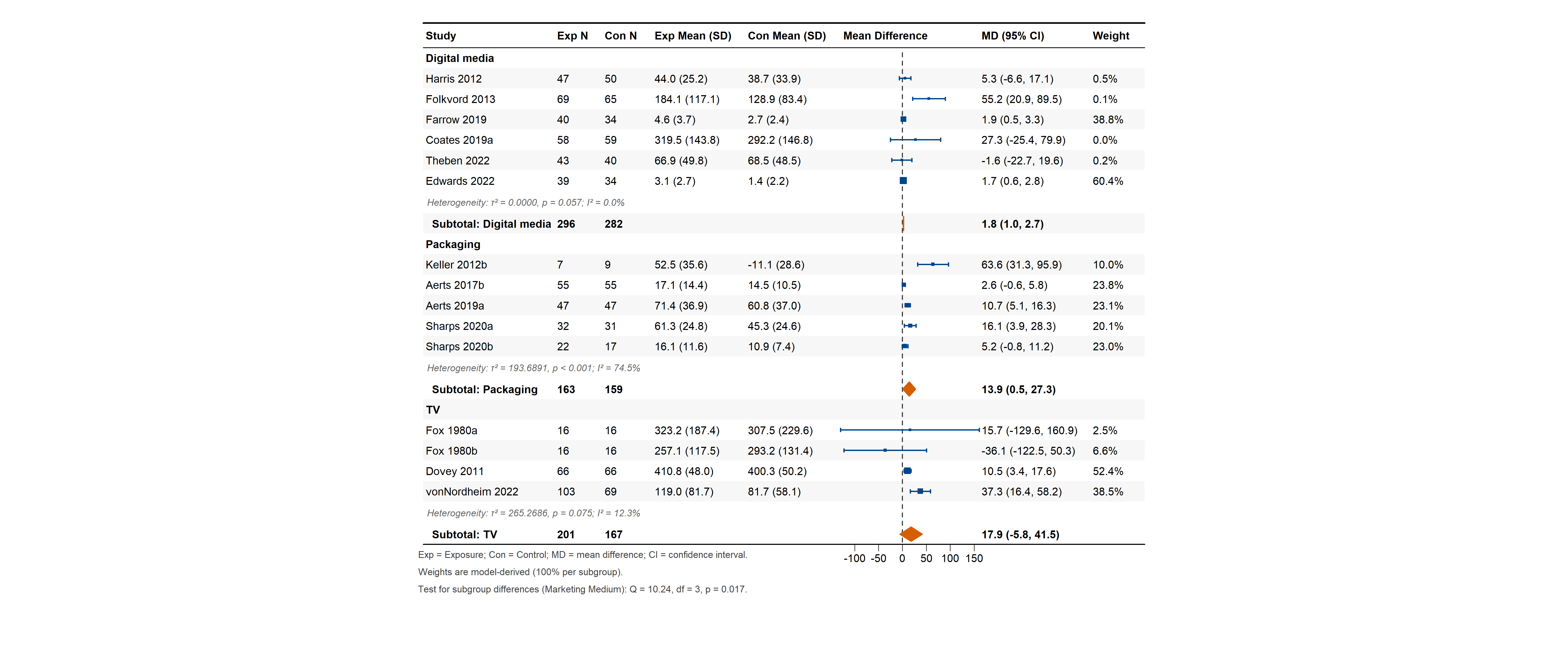

### **eFigure 4.** *The Effect of Unhealthy M2K on Dietary Intakes by Age Groups in RCTs*

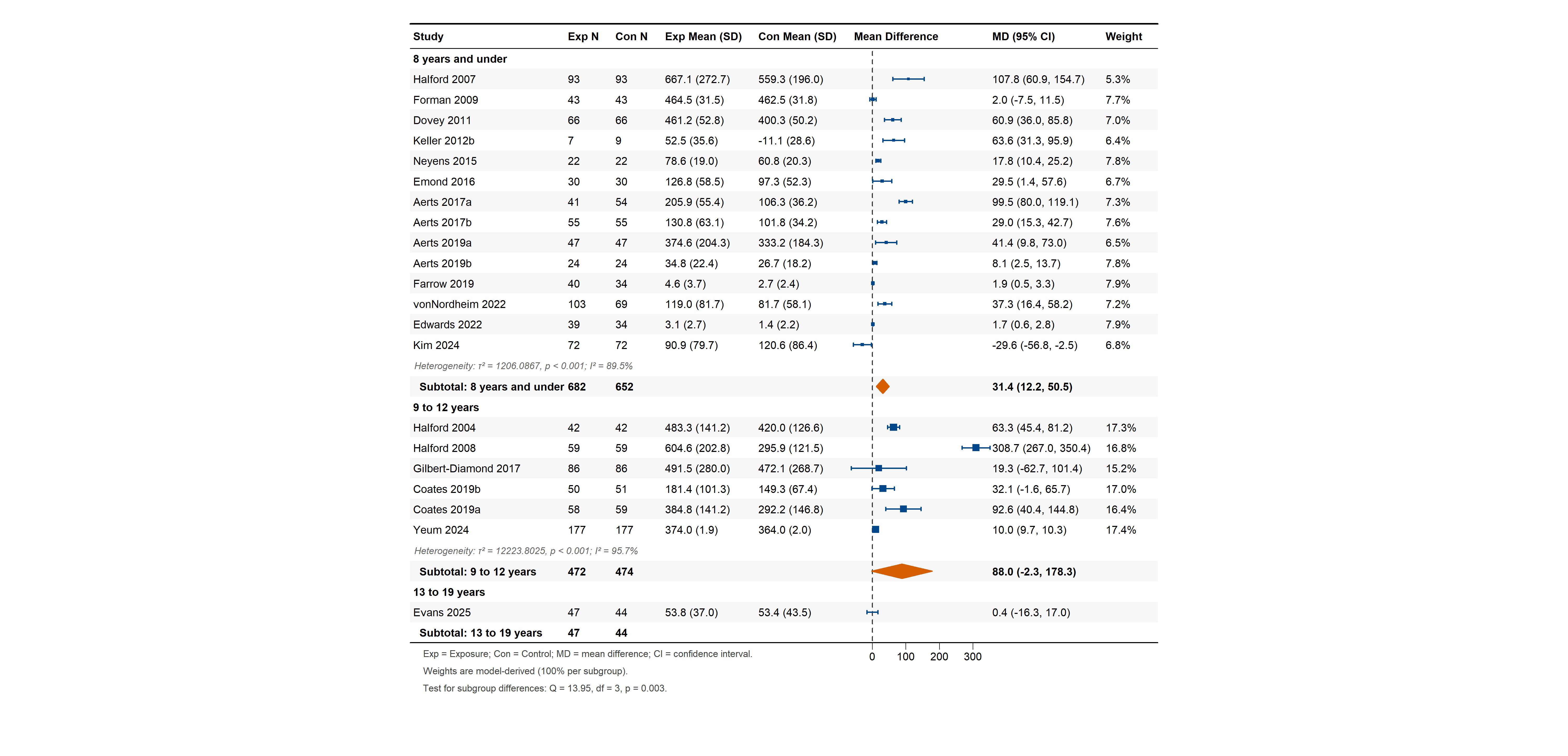

### **eFigure 5.** *The Effect of Unhealthy M2K on Dietary Intakes by Sex in RCTs*

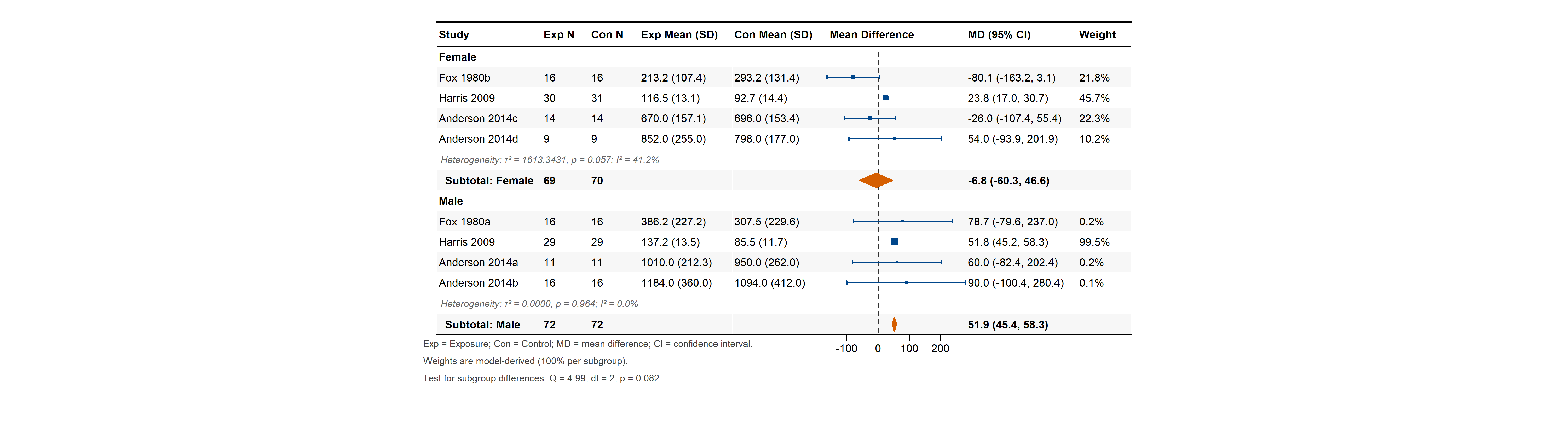

### **eFigure 6.** *The Effect of Unhealthy M2K on Dietary Intakes by Weight Status in RCTs*

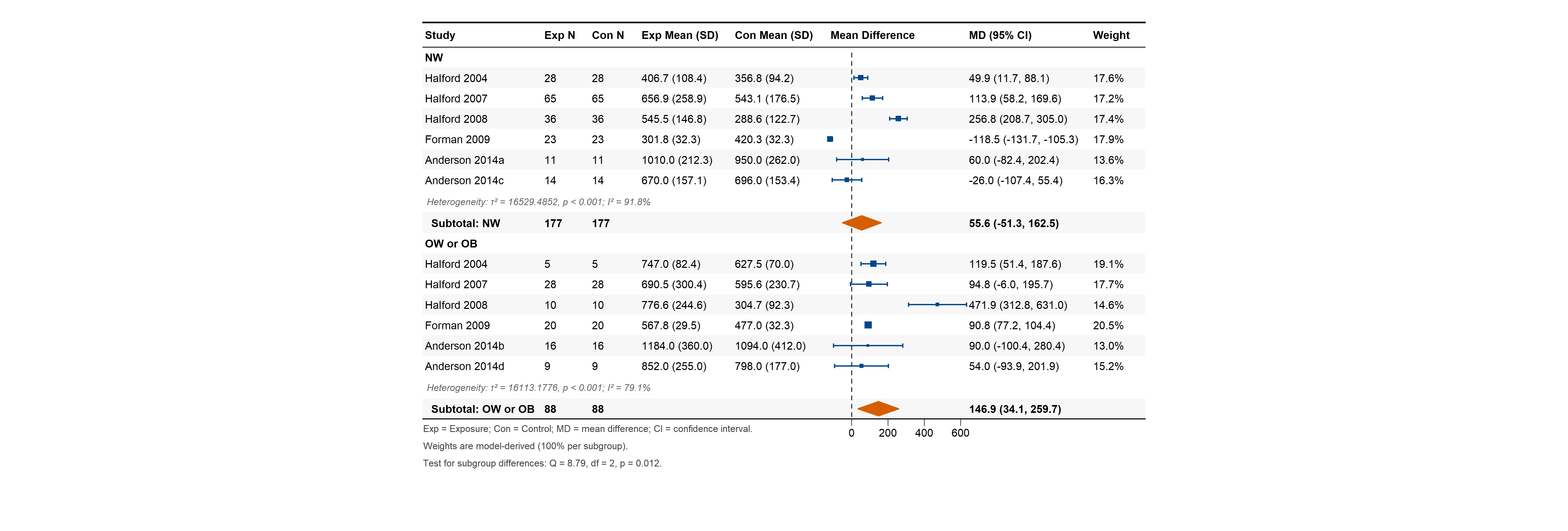

### **eFigure 7.** *The Effect of Unhealthy M2K on Dietary Intakes by Exposure Duration in RCTs*

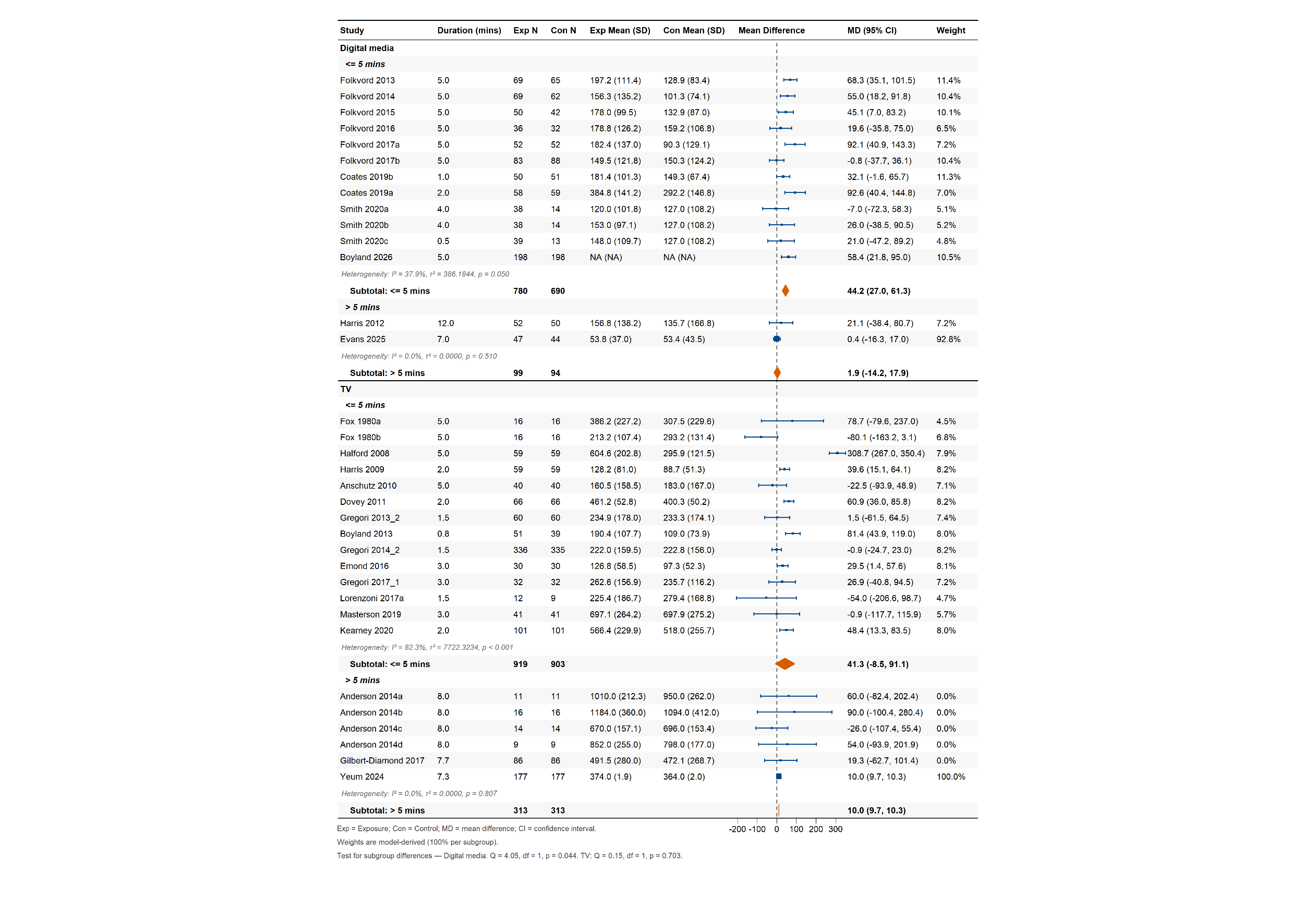

### **eFigure 8.** *Trim-and-fill Analysis: Adjusted Funnel Plot*

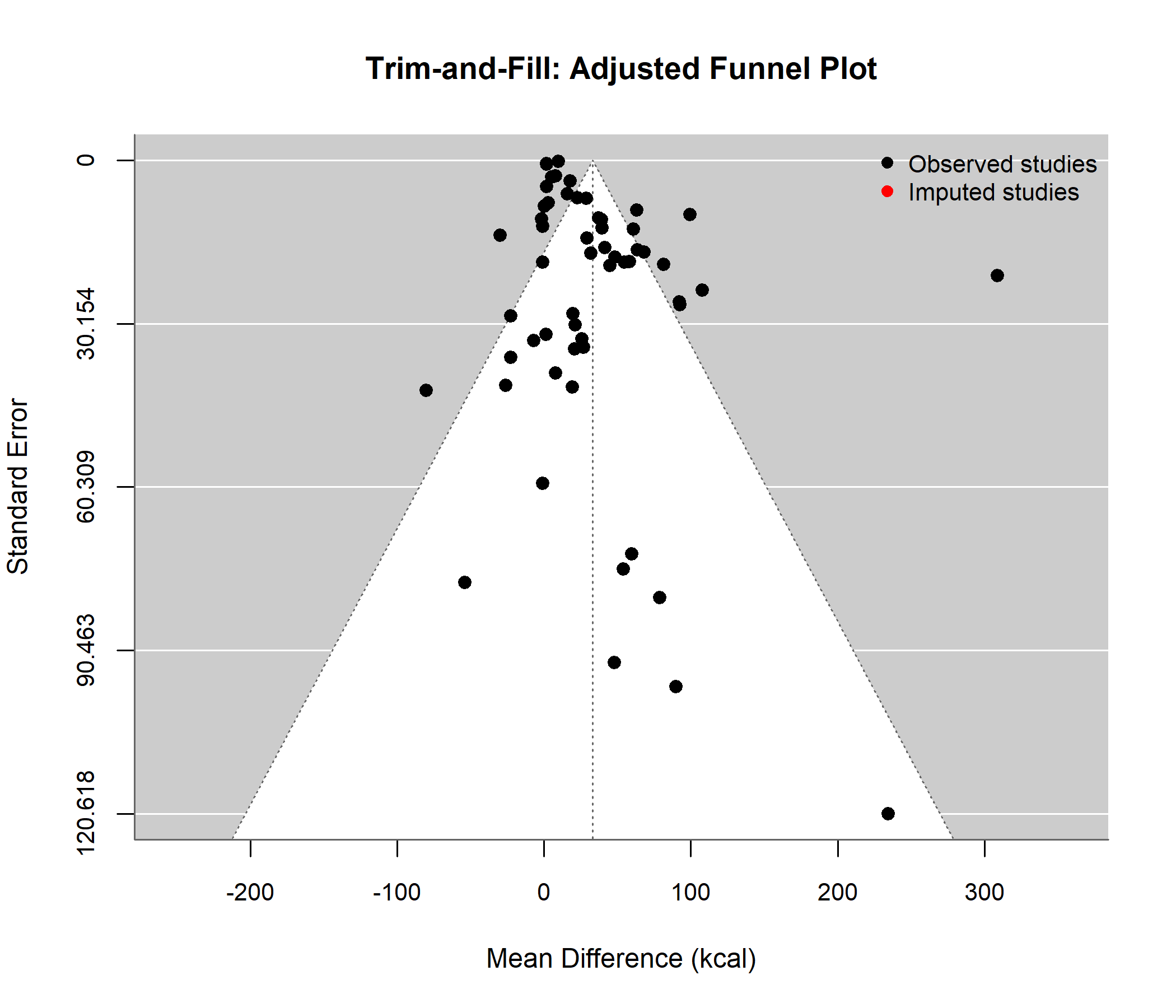

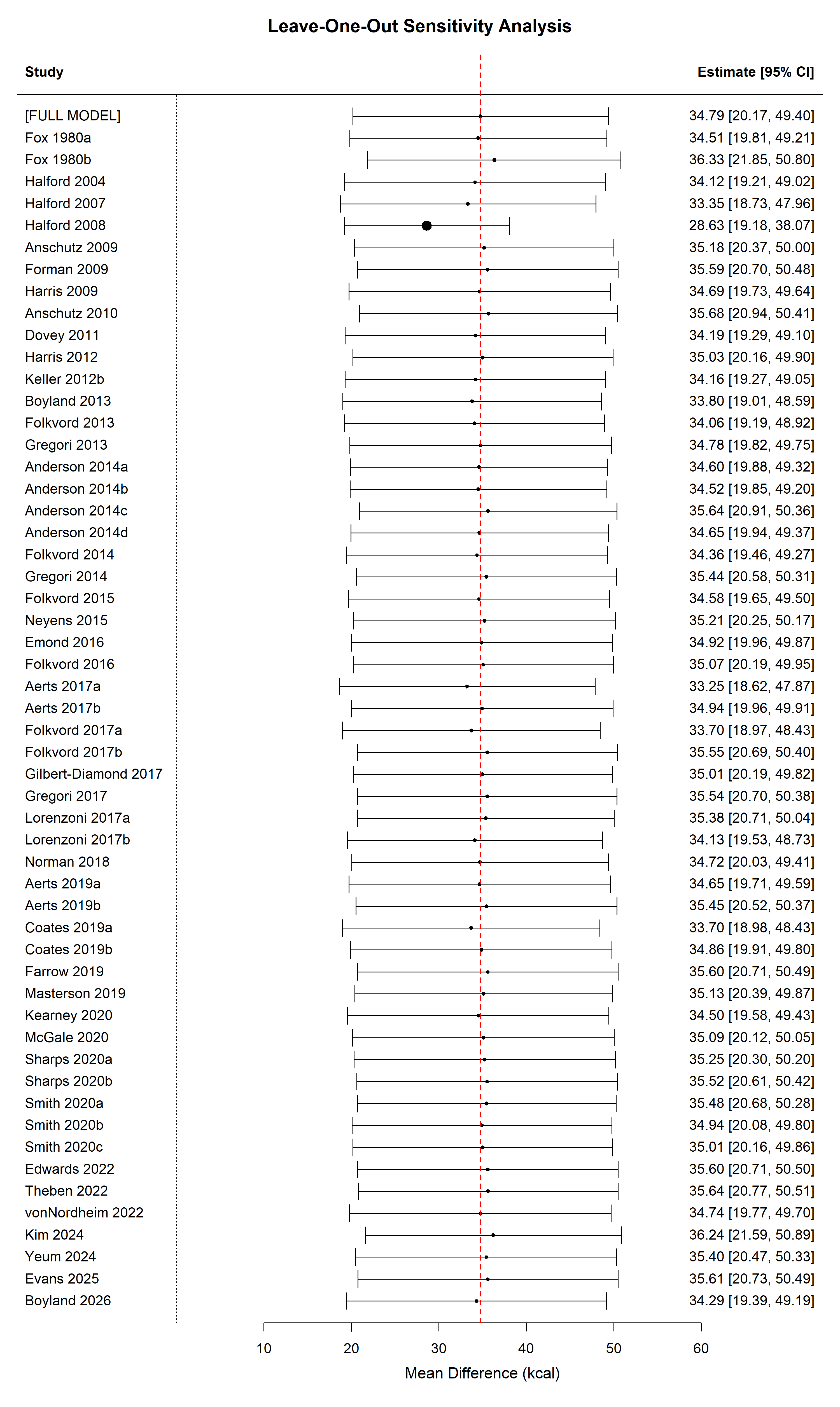

### **eFigure 9.** *Diagnostic Analysis: Funnel Plot, Cook’s Distance, and Leave-One-Out Analysis*

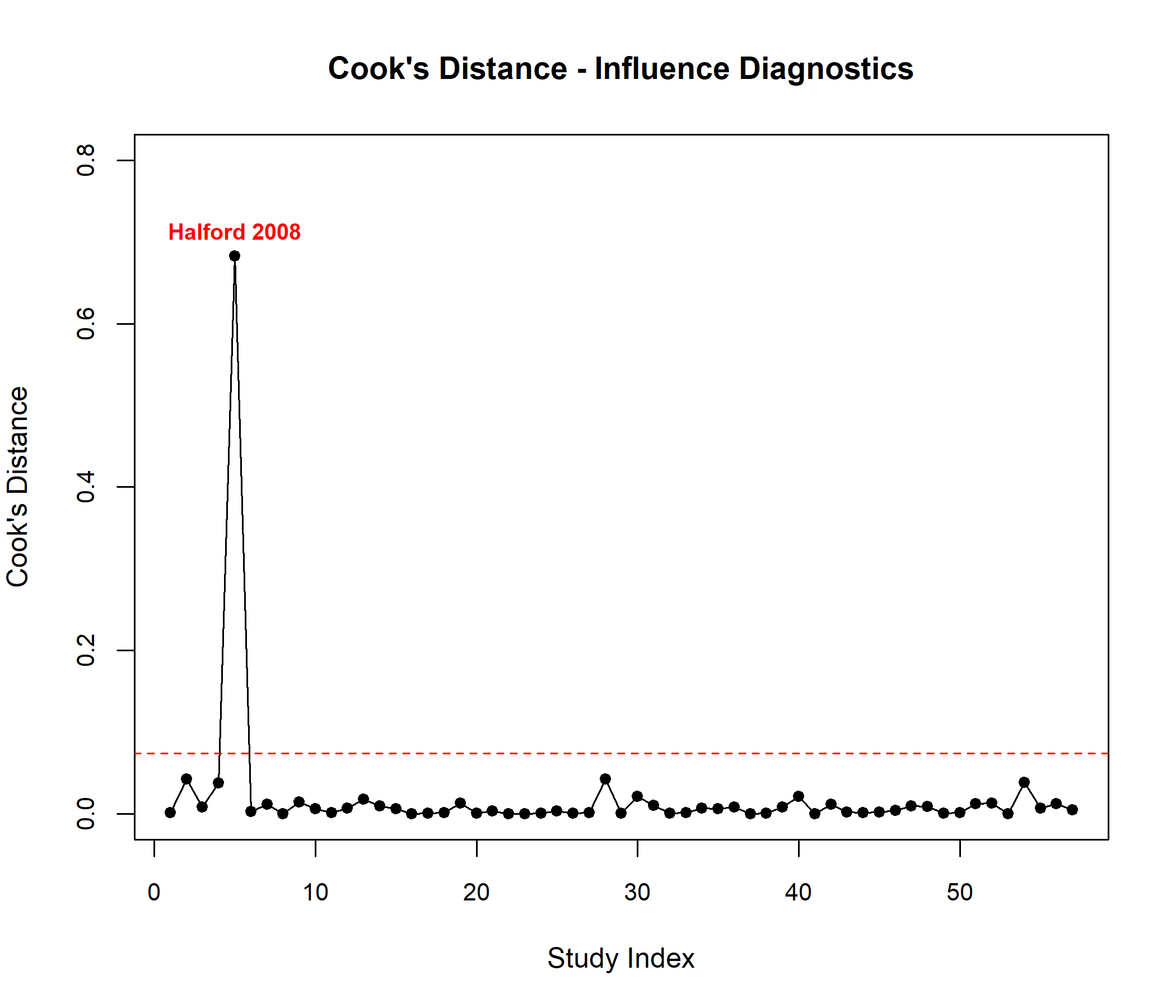

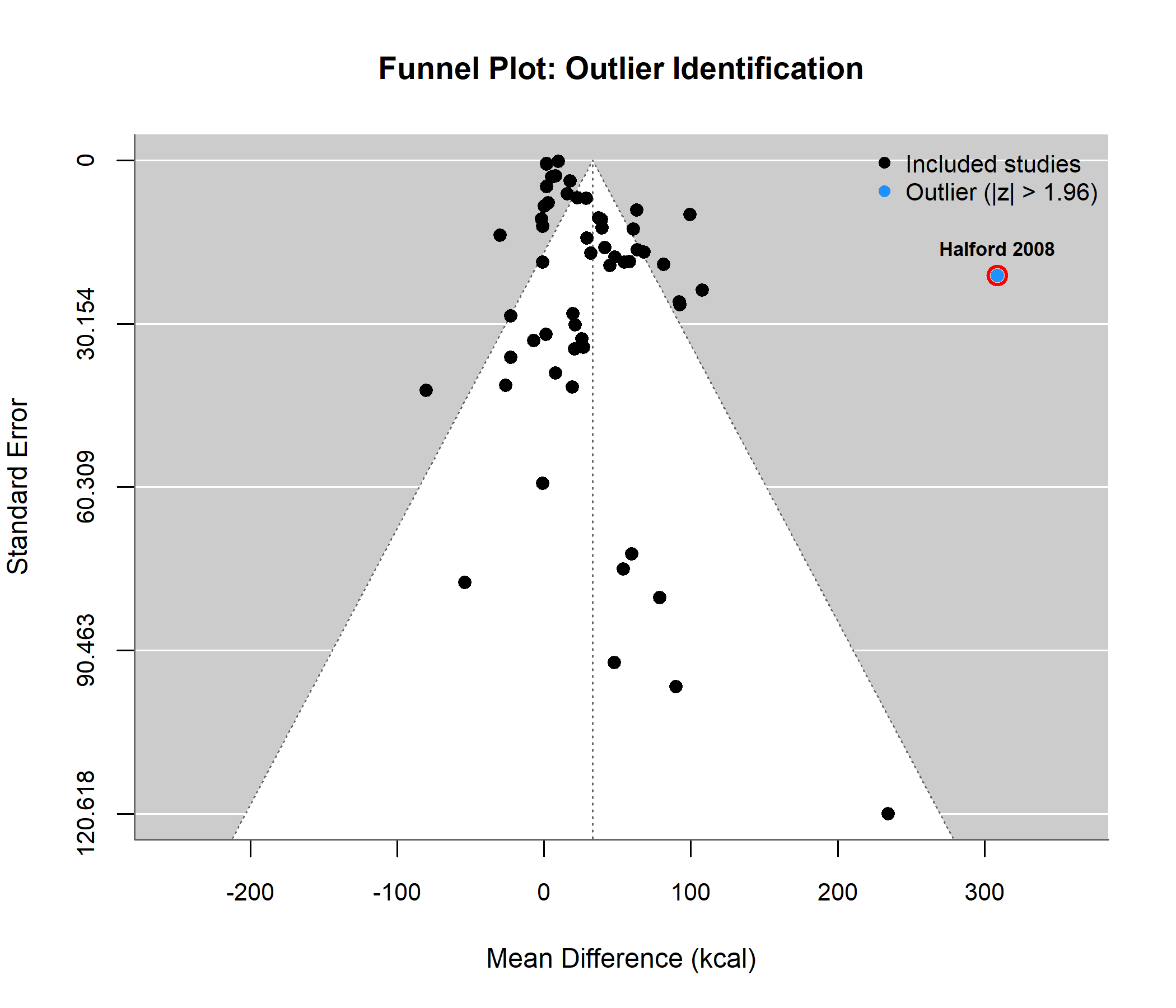

### **eMethod.** *Converting Dietary Intakes to Energy Intake in Kilocalories (Kcal)*

To address the variety of measurement units, all outcomes were converted to kcal of energy intake. For example, food weight reported in ounces or grams was converted to kcal by a Registered Dietitian using the nutrient content reported in the study. If such information was not available, the food company’s website, Open Food Facts (https://world.openfoodfacts.org/), or the Canadian Nutrient File (https://food-nutrition.canada.ca/cnf-fce/) was used to identify the nutrient content. Energy intake reported in kilojoules was converted to kilocalories (1 kcal = 4.184 kJ).
